# Safety and immunogenicity of heterologous boost immunization with an adenovirus type-5-vectored and protein-subunit-based COVID-19 vaccine (Convidecia/ZF2001): a randomized, observer-blinded, placebo-controlled trial

**DOI:** 10.1101/2022.02.24.22271445

**Authors:** Pengfei Jin, Xiling Guo, Wei Chen, Shihua Ma, Hongxing Pan, Lianpan Dai, Pan Du, Lili Wang, Lairun Jin, Yin Chen, Fengjuan Shi, Jingxian Liu, Xiaoyu Xu, Yanan Zhang, George F. Gao, Cancan Chen, Jialu Feng, Jingxin Li, Fengcai Zhu

## Abstract

**Background:** Heterologous boost vaccination has been proposed as an option to elicit stronger and broader, or longer-lasting immunity. We assessed the safety and immunogenicity of heterologous immunization with a recombinant adenovirus type-5-vectored COVID-19 vaccine (Convidecia) and a protein-subunit-based COVID-19 vaccine (ZF2001).

**Methods and Findings:** We did a randomized, observer-blinded, placebo-controlled trial in healthy adults previously received one dose of Convidecia. Participants were randomly assigned (2:1) to receive either ZF2001 (vaccine group) or a trivalent inactivated influenza vaccine (TIV) (placebo group) at either 28-day or 56-day intervals. For both regimens, all participants received the 2nd injection with ZF2001 at 4 months after a dose of ZF2001 or TIV, with three-dose schedules of Convidecia/Convidecia/ZF2001 at day 0, day 28 and month 5 (referred to as CV/ZF/ZF (D0-D28-M5)) and CV/ZF/ZF (D0-D56-M6), and two-dose schedules of CV/ZF (D0-M5) and CV/ZF (D0-M6). The primary outcome was the geometric mean titer (GMT) of the neutralizing antibodies against live SARS-CoV-2 virus 14 days after each boost vaccination. The safety outcome was 7-day reactogenicity, measured as solicited local or systemic adverse reactions after each vaccination. Between April 7, 2021, and May 6, 2021, 120 participants were enrolled, among whom 60 were randomly assigned to receive ZF2001 (n=40) or TIV (n=20) at a 28-day interval, and 60 were randomly assigned to receive ZF2001 (n=40) or TIV (n=20) at a 56-day interval. 113 (94.2%) participants received the 2nd injection with ZF2001 4 months after a dose of ZF2001 or TIV.

A total of 26 participants (21.7%) reported solicited adverse events within 7 days post boost vaccinations, and all the reported adverse reactions were mild. Among participants receiving ZF001 as second dose, the GMTs of neutralizing antibodies increased to 58.4 IU/ml (42.8-79.8) in 0-28 regimen, and to 80.8 IU/ml (53.1-122.9) in 0-56 regimen at 14 days post first boost dose. The GMTs of neutralizing antibodies increased to 334.9 IU/ml (95% CI 230.4, 486.9) in C/Z/Z (D0-D28-M5) regimen, and 441.2 IU/ml (260.8, 746.4) in C/Z/Z (D0-D56-M6) regimen at 14 days after the third dose. Two-dose schedules of CV/ZF (D0-M5) and CV/ZF (D0-M6) induced comparable antibody level comparable with that elicited by three-dose schedules, with the GMTs of 282.9 IU/ml (142.5, 561.8) and 293.9 IU/ml (137.6, 627.9), respectively. Study limitations include the absence of vaccine effectiveness in real-world, and current lack of immune persistence data and the neutralizing antibodies to Omicron.

**Conclusions:** Heterologous boosting with ZF001 following primary vaccination of Convidecia is safe and more immunogenic than a single dose of Convidecia. These results support flexibility in cooperating viral vectored vaccines and recombinant protein vaccine.

**Trial Registration:** ClinicalTrial.gov NCT04833101

## Introduction

Coronavirus disease 2019 (COVID-19), caused by severe acute respiratory syndrome coronavirus 2 (SARS-CoV-2), has severely impacted the world in terms of health, society, and economy(1). The mass vaccination campaigns are fundamental to reducing the burden of disease, and the subsequent economic recovery. As of 16 February 2022, 10.4 billion doses have been administered globally, and 61.9% of the world population has received at least one dose of a COVID-19 vaccine, but only 10.6% of people in low-income countries have received at least one dose(2). Currently, national regulatory authorities have granted authorizations for more than 15 COVID-19 vaccines, including four adenovirus-based vector vaccines: ChAdOx1 n CoV-19 (AstraZeneca), Ad26.COV 2-S (Janssen), rAd26+rAd5 (Gamaleya), and Ad5 nCoV (CanSino, Convidecia). Of total procured doses in the worldwide, 30.6 % is adenovirus vectored vaccines(3).

Compared with other COVID-19 vaccines approved as two-dose homologous schedules (e.g. BNT162b2, mRNA-1273, NVXCoV2373), a single dose of Ad26.COV2-S and Ad5 nCoV only showed a relatively lower immunogenicity and efficacy against symptomatic disease(4, 5). With the waning of antibodies against SARS-CoV-2 coincident with the emergence of the new variants, the vaccine effectiveness of COVID-19 vaccines declined over time(5–7), which raised the need to boost vaccination. For adenovirus-vectored vaccines, pre-existing adenovirus immunity is the biggest obstacle for homologous immunization to overcome. Hence, the Sputnik V vaccine programmer deployed a heterologous prime-boost schedule using Ad26 and Ad5 vectored COVID-19 vaccines, induced robust humoral and cellular responses and showed 91.5% efficacy against COVID-19(8, 9). Additionally, the occurrence of rare, but severe thrombotic events with thrombocytopenia is other challenge for adenovirus-based vaccines(10–12). Based on the both the concerns of long-term protective effect and safety, it has been recommended to heterologous immunization with ChAdOx1 n CoV-19 or Ad26.COV 2-S followed by an mRNA vaccine(13, 14).

Heterologous regimens have been proposed as an option to elicit stronger and broader, or longer-lasting immunity, which is particularly important for COVID-19 vaccines with moderate vaccine efficacy. The results from clinical trials and real-world studies suggested that heterologous prime-boost vaccination of adenovirus vectored vaccines (ChAdOx1-S or Ad26.COV 2-S) followed by mRNA vaccines (BNT162b2 or mRNA-1273) induced stronger immune responses, and provided higher effectiveness than homologous ChAdOx1-S vaccination(14, 15). Additionally, the results from Com-COV2 and COV-BOOST trials showed heterologous immunization with ChAdOx1 nCoV-19 and NVXCoV2373 induced both humoral and T-cell immune responses superior to that homologous ChAdOx1 nCoV-19 vaccination(16, 17). Robust data on the safety and immunogenicity of heterologous schedules with different COVID-19 vaccines will help enhance deployment flexibility and improve access to vaccines.

Here, we present safety and immunogenicity of a heterologous prime-boost vaccination of a recombinant adenovirus type-5-vectored COVID-19 vaccine (Convidecia) followed by a protein-subunit-based COVID-19 vaccine (ZF2001) in healthy adults.

## Methods

### Study design and participants

This study was designed as a randomized, observer-blinded, placebo-controlled trial to access the safety and immunogenicity of a heterologous prime-boost immunization with Convidecia and ZF2001 in Guanyun County, Jiangsu Province, China. In the original protocol, we planned to implement one dose of boosting at prime-boost intervals (28 days or 56 days) in individuals receiving Convidecia. For each heterologous prime-boost regimen, eligible participants were randomly (2:1) assigned to vaccine group receiving a dose of ZF2001 or placebo group receiving a dose of TIV. We made a protocol change to add an additional boost vaccination with ZF2001 at 4 months after the 1st boost dose to further boost the immune responses for all the participants. Four permutations of prime-boost schedule were investigated, including heterologous immunization of Convidecia/ZF2001/ZF2001 at an interval of day 0, day 28 and month 5 (referred to as CV/ZF/ZF (D0-D28-M5)), Convidecia/ZF2001 at an interval of day 0 and month 5 (CV/ZF (D0-M5)), Convidecia/ZF2001/ZF2001 at an interval of day 0, day 56 and month 6 (CV/ZF/ZF (D0-D56-M6)) and Convidecia/ZF2001 at an interval of day 0 and month 6 (CV/ZF (D0-M6)). The trial protocol was reviewed and approved by the institutional review board of the Jiangsu Provincial Center of Disease Control and Prevention (approval number: JSJK2021-A005-02). This study is registered with NCT04833101. The study was done in accordance with the Declaration of Helsinki and Good Clinical Practice.

Participants were healthy adults aged 18 years and above who have received a prime Convidecia vaccination within 28 days before the screening visit. Volunteers with a previous clinical or virologic COVID-19 diagnosis or SARS-CoV-2 infection, diagnosis of an immunocompromising or immunodeficiency disorder, or those who have received immunosuppressive therapy were excluded. Women with positive urine pregnancy test were also excluded from this study. Full details of the inclusion and exclusion criteria could be founded in the supplementary material (Table S5). All participants provided written informed consent before enrollment.

### Vaccines

Convidecia and ZF2001 have been authorized for conditional license or emergency use against COVID-19 in China and other countries, and manufactured by CanSino Biologics Inc. and Anhui Zhifei Longcom Biopharmaceutical Co., Ltd., respectively. ZF2001 is a dimeric form of SARS-CoV-2 spike receptor-binding domain (RBD) adjuvanted with aluminium hydroxide, containing 25μg antigen per 0·5mL in a vial(18). The control influenza vaccine is produced by Dalian Aleph Biomedical Co., Ltd. Administration is via 0.5mL intramuscular injection into the upper arm for both ZF2001 and TIV.

### Randomization and masking

The randomization lists were generated by an independent statistician using SAS (version 9·4). Participants were stratified by age (18-59 years and ≥ 60 years), and then randomly assigned (2:1) to vaccine group receiving ZF2001 and “placebo” group receiving a trivalent inactivated influenza vaccine (TIV) either 28 days or 56 days apart.

We masked participants, investigators, laboratory staff, and outcome assessors to the allocation of treatment groups, but not the prime-boost intervals. Personals who prepared and administered vaccination were aware of group allocation, but they were not otherwise involved in other trial procedures or data collection, and were instructed not to reveal the identity of the study vaccines to the participants and other investigators.

### Procedures

We recruited eligible participants who had primed with one dose of Convidecia, and assigned them to either “0-28 days” regimen group or “0-56 days” regimen group. Eligible participants received one shot of ZF2001 or TIV in a ratio of 2:1 at 28 days or 56 days after the prime vaccination. Four months after receiving the 1st boost dose, all the participants were administrated with an additional dose of ZF2001. Participants were monitored for 30 min post each vaccination for any immediate adverse reactions and instructed to record solicited adverse events up to day 7 and unsolicited adverse events up to day 28 after each dose on paper diary cards. Serious adverse events self-reported by participants were documented throughout the study. Adverse events were graded as mild (grad 1), moderate (grad 2), severe (grade 3), or life-threatening (grade 4) according to the scale issued by the China State Food and Drug Administration (version 2019).

Blood samples were taken from participants for serology tests at baseline (28 days post the prime vaccination), Day 14, Day 28 post the 1st boost dose, and Day14, Month 6 post the 2nd boost dose. Participants in “0-56 days” regimen group had an additional blood test at Day 0 before the 1st boost dose. Additionally, serum samples from 40 participants receiving homologous immunization of Convidecia following a 0-56 days regimen and 20 participants receiving homologous prime-boost vaccination of Convidecia with a 0-6 months regimen in previous trials(19, 20), were tested in live viral microneutralization assay side by side as external comparators.

### Outcomes

The primary outcomes were the occurrence of solicited local or systemic adverse reactions within 7 days post vaccination and the live virus neutralizing antibody titers against wild-type SARS-CoV-2 isolate at day 14 after each boost vaccination.

Safety secondary outcomes include unsolicited adverse events for 28 days after immunization and serious adverse events collected throughout the study. Immunological secondary outcomes include the binding IgG concentration against SARS-CoV-2 RBD and spike protein at day 14 after each boost vaccination, and live virus neutralization titers and binding IgG concentration at days 28 post prime dose, at days 28 post 1st boost dose and at months 6 post 2nd boost dose.

The exploratory outcomes were live virus neutralizing antibodies against delta variant B.1.617.2 at 28 days post prime dose and 14 days post 2nd boost dose, and cellular responses measured by IFN γ ELISpot in peripheral blood at 28 days post prime dose and 14 days post 1st boost dose.

### Immunogenicity assay

#### Microneutralization assay

SARS-CoV-2-specific neutralizing antibody titer in serum was determined using a cytopathic effect (CPE)-based microneutralization assay with the SARS-CoV-2 virus strain in Vero-E6 cells (wild-type SARS-CoV-2: BetaCoV/Jiangsu/JS02/2020 (EPI_ISL_411952); delta variant B.1.617.2: hCoV-19/China/JS07/2021(EPI_ISL_4515846)), as described previously(21). The titer of neutralizing antibodies was calculated as 50% tissue culture infectious dose of 100 in each well (100 TCID_50_), expressed as the reciprocal of two-fold serial dilution of heat-inactivated sera. The serum dilution for microneutralization assay started from 1:8 to 1:256 for wild-type, and 1:4 to 1:128 for delta variant B.1.617.2. If no neutralization reaction was observed at the initial serum dilution, half of the limit of quantification was calculated.

#### Binding-antibody assays

Binding antibodies against receptor-binding domain (RBD) and spike protein were detected by an indirect ELISA assay with a cutoff titer of 1:10. The commercial ELISA kits (Vazyme Biotech Co.,Ltd) were used for the detection. Briefly, serum samples were serially diluted (anti-RBD antibody detection, 1:10 to 1:1280; anti-Spike antibody detection, 1:10 to 1:21870) with sample diluent and tested in 96-well plates costed with a recombinant RBD or Spike antigen. IgG was detected using an anti-human IgG monoclonal antibody conjugated to horseradish peroxidase (HRP) diluted for each ELISA assay and TMB substrate. Data collection was performed using a Multiskan GO reader (Thermo Fisher) to detect optical density (OD) at 450 and 630 nm using SkanIt Software for Microplate Readers (version 4.1.0.43). A monoclonal antibody with neutralizing activity specific to the SARS-CoV-2 RBD or Spike protein was used as a calibrator, which was used to generate a standard curve to convert OD units into relative units per milliliter (RU/ml) in the ELISA.

#### Enzyme-linked immunospot (ELISpot) assay

The cellular immune responses of the expression of interferon (IFN) γ stimulated by the overlapping peptide pool of spike glycoprotein were detected by ELISpot assay (Mabtech, Stockholm, Sweden). PBMCs were isolated by Ficoll-Paque PLUS (Cytiva) density gradient centrifugation and cryopreserved before analysis. Per well, 100,000 isolated PBMCs were stimulated with peptide pools covering the full-length spike glycoprotein at a concentration of 1μg in the plates. Plates were scanned, and spots were counted on the Cellular Technology ImmunoSpot Analyzer (AID Diagnostika GmbH) for AID EliSpot 7.0 software. IFN γ-secreting spots forming cells was calculated as the number of spots forming cells in the presence of peptides minus the number of that without peptides, and were multiplied by ten to express frequencies pre 10^6^ PBMCs.

#### The calibration and harmonization of WHO international standard

The WHO international standard for anti-SARS-CoV-2 immunoglobulin (NIBSC code 20/136) was used side by side as reference with the serum samples measured in this study for calibration and harmonization of the serological assays(22). The WHO reference serum 1000 IU/mL equivalent to live viral neutralizing antibody titer of 1:320 against the wild-type and binding antibody concentrations of 1000 RU/ml. Seroconversion was defined as at least a fourfold increase in the antibody titers at different time points after boost immunization compared to baseline level (at 28 days post prime dose).

### Statistical analyses

We assumed that the GMT of neutralizing antibodies was about 1:20 at baseline (28 day after one dose of prime vaccination with Convidecia). After the second dose, GMT in the vaccine group was expected to reach 1:60 at day 14 post boost vaccination, while that in the control group remained unchanged. Assuming a standard deviation of 4, 40 and 20 participants receiving vaccine and placebo control in each regimen group, respectively, was estimated to provide 81·6% power for declaring the superiority.

We assessed the number and proportion of participants with adverse reactions post vaccination. The antibodies against SARS-CoV-2 were presented as GMTs, GMFIs and seroconversion with 95% CIs, and the cellular responses were shown as the average number of positive cells per PBMCs. We used the χ² test or Fisher’s exact test to analyze categorical data, T test to analyze the log transformed antibody titers, and Wilcoxon rank-sum test for non-normal distributed data. The correlation between concentrations of log-transformed neutralizing antibodies and binding antibody was analyzed using Spearman’s correlation with 95% CIs. Hypothesis testing was two-sided with an α value of 0·05. Statistical analyses were done by a statistician using SAS (version 9·4) or GraphPad Prism 8.0.1.

## Results

### Study participants

Between April 7, 2021, and May 6, 2021, a total of 120 adults over 18 years of age who had received a primary dose of Convidecia were enrolled, among whom 60 were randomly assigned (2:1) to receive a dose of ZF2001 (vaccine group, n=40) or a trivalent inactivated influenza vaccine (TIV) (placebo group, n=20) at an interval of 28 days, and 60 were randomly assigned (2:1) to receive a dose of ZF2001 (n=40) or TIV (n=20) at an interval of 56 days. 113 (94.2%) participants received the 2nd injection with ZF2001 4 months after a dose of ZF2001 or TIV, with 40 receiving heterologous immunization of Convidecia/ZF2001/ZF2001 at an interval of day 0, day 28 and month 5 (referred to as CV/ZF/ZF (D0-D28-M5) regimen), 19 receiving CV/ZF (D0-M5) regimen, 36 receiving CV/ZF/ZF (D0-D56-M5) regimen, and 18 receiving CV/ZF (D0-M6) regimen (Figure 1). The mean age was 54.0 years (SD 15.0) for the whole study cohort, with 57 (47.5%) female participants. Baseline characteristics of the participants were similar across the four regimens (Table 1). For extend comparator cohorts, the mean age of participants receiving homologous vaccination of Convidecia with a 0-56 days regimen (n=40) and 0-6 months regimen (n=20) was 59.0 years and 40.2 years, respectively (Table S1).

**Fig. 1.**
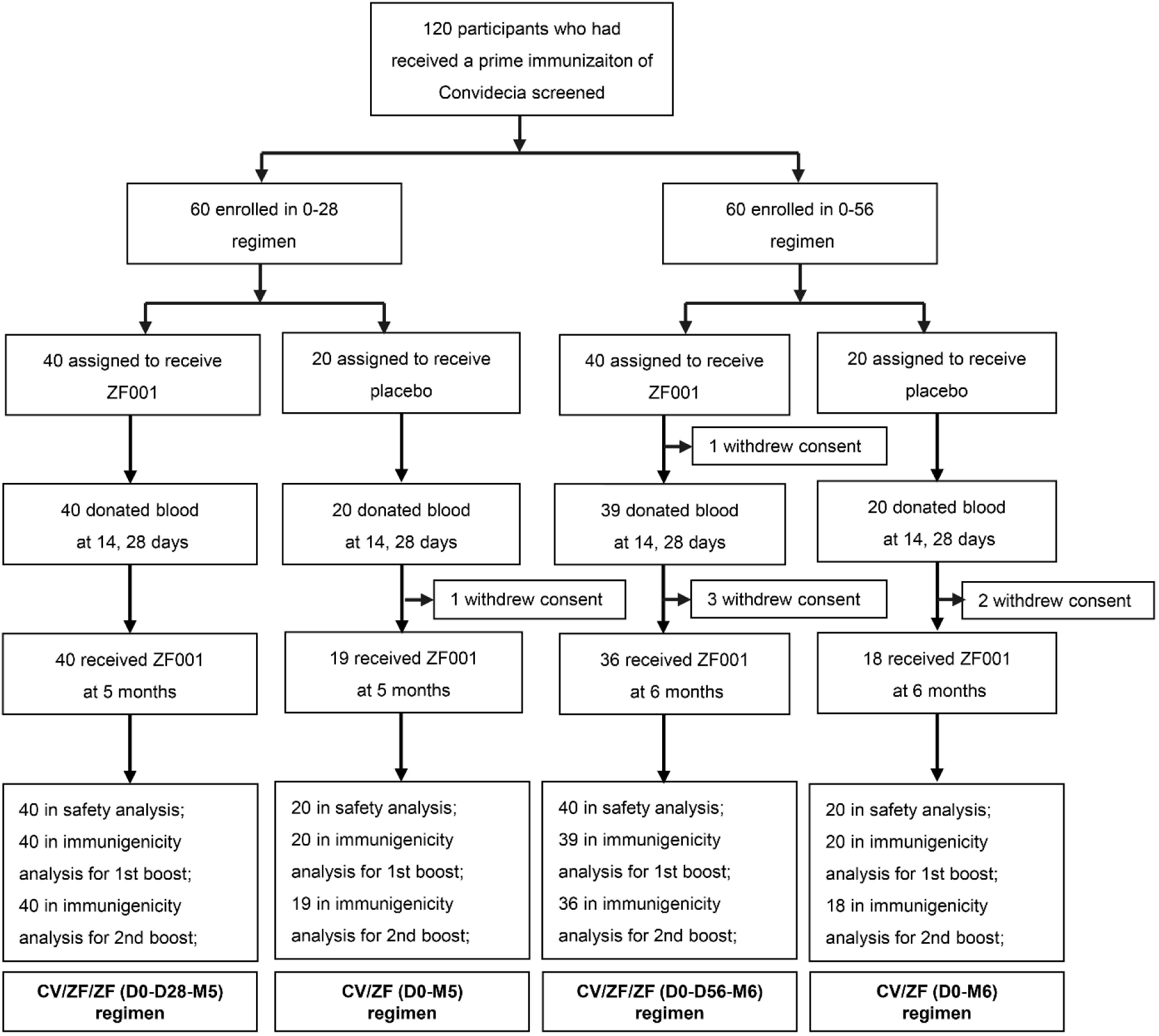
Trial profile.

**Table 1.**
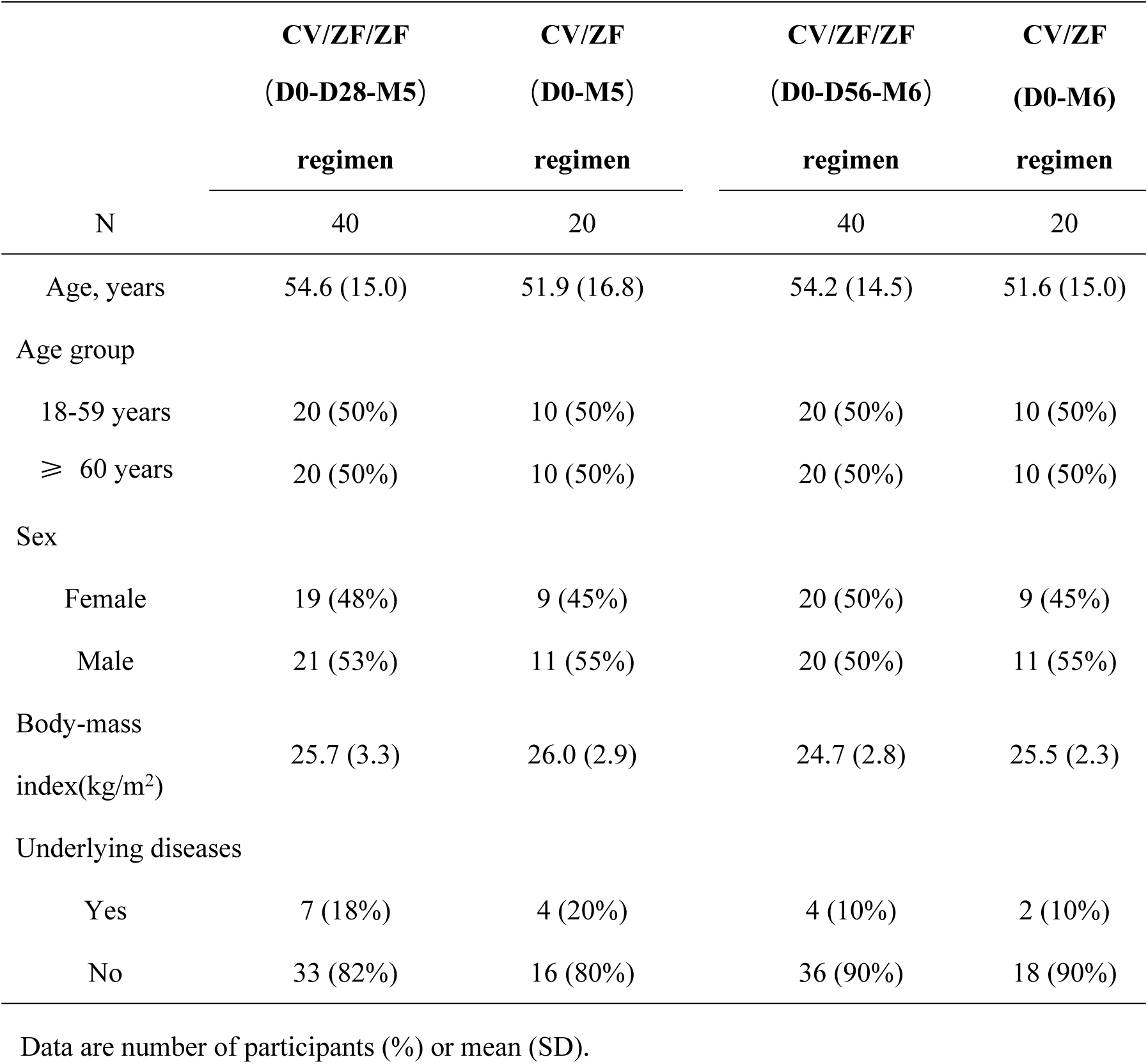
Baseline characteristics of the participants by vaccination schedules.

### Safety

A total of 26 participants (21.7%) reported solicited adverse events within 7 days post boost vaccination, with 13 (32.5%) in CV/ZF/ZF (D0-D28-M5) regimen, 7 (35.0%) in CV/ZF (D0-M5) regimen, 4 (10.0%) in CV/ZF/ZF (D0-D56-M6) regimen, and 2 (10.0%) in CV/ZF (D0-M6) regimen, respectively (Table 2). All the reported adverse reactions post boost dose were mild, and the most common adverse reaction was injection-site pain (20.0%, 24/120).

**Table 2.** Adverse reactions occurred within 7 days and unsolicited adverse events within 28 days post boost vaccination.

|  |  | Boost dose (N=120) |  |  |  |
| --- | --- | --- | --- | --- | --- |
|  | Prime dose | CV/ZF/ZF | CV/ZF | CV/ZF/ZF | CV/ZF |
|  | (N=120) | (D0-D28-M5) | (D0-M5) | (D0-D56-M6) | (D0-M6) |
|  |  | (N=40) | (N=20) | (N=40) | (N=20) |
| Adverse reaction within 7 days post vaccination |  |  |  |  |  |
| Total | 21 (17.5) | 13 (32.5) | 7 (35.0) | 4 (10.0) | 2 (10.0) |
| Injection-site adverse reaction within 7 days post vaccination |  |  |  |  |  |
| Total | 9 (7.5) | 13 (32.5) | 7 (35.0) | 3 (7.5) | 2 (10.0) |
| Pain | 9 (7.5) | 13 (32.5)* | 6 (30.0) | 3 (7.5)* | 2 (10.0) |
| Redness | 0 (0.0) | 0 (0.0) | 1 (5.0) | 0 (0.0) | 0 (0.0) |
| Swelling | 1 (0.8) | 1 (2.5) | 0 (0.0) | 0 (0.0) | 0 (0.0) |
| Induration | 0 (0.0) | 1 (2.5) | 0 (0.0) | 0 (0.0) | 0 (0.0) |
| Systemic adverse reaction within 7 days post vaccination |  |  |  |  |  |
| Total | 15 (12.5) | 1 (2.5) | 0 (0.0) | 1 (2.5) | 0 (0.0) |
| Fever | 10 (8.3) | 0 (0.0) | 0 (0.0) | 1 (2.5) | 0 (0.0) |
| Grade 3 | 2 (1.7) | 0 (0.0) | 0 (0.0) | 0 (0.0) | 0 (0.0) |
| Fatigue | 4 (3.3) | 0 (0.0) | 0 (0.0) | 0 (0.0) | 0 (0.0) |
| Headache | 2 (1.7) | 0 (0.0) | 0 (0.0) | 0 (0.0) | 0 (0.0) |
| Muscle pain | 1 (0.8) | 0 (0.0) | 0 (0.0) | 0 (0.0) | 0 (0.0) |
| Cough | 2 (1.7) | 1 (2.5) | 0 (0.0) | 0 (0.0) | 0 (0.0) |
| Unsolicited adverse event within 28 days post vaccination |  |  |  |  |  |
| Total | 7 (5.8) | 2 (5.0) | 1 (5.0) | 1 (2.5) | 2 (10.0) |

Data are n (%): n = the number of participants, % = proportion of participants; N= the number of participants included in the safety analysis. \*There was significant difference for the incidence of injection-site pain between CV/ZF/ZF (D0-D28-M5) regimen and CV/ZF/ZF (D0-D56-M6) regimen ( $p=0.0052$ ).

Adverse reactions occurring within 7 days after prime immunization with Convidecia were reported by 17.5% (21/120) of the total participants, with injection-site pain (7.5%), fever (8.3%), and fatigue (3.3%) as the most commonly reported symptoms (Table 2). Two participants had grade 3 fever (axilla temperature ≥ 38.5℃) after prime vaccination. As of February 8, 2022, no serious adverse events were observed, and no prespecified trial-halting rules per protocol were met during the study.

### Immunogenicity

#### Neutralizing antibody responses against wild-type virus

The neutralization responses were detectable in 58.3% (35/60) of the participants aged 18-59 years, and in 43.3% (26/60) of the participants aged ≥60 years at 28 days post prime vaccination with Convidecia. A heterologous boost dose with ZF001 induced significantly higher neutralizing antibody against SARS-CoV-2 wild-type virus compared with the baseline (day 28 after prime vaccination with Convidecia) (Figure 2A and 2B). In the vaccine group, geometric mean titers (GMTs) of the neutralizing antibodies increased from 23.7 IU/ml (95% CI 18.0-31.3) at baseline to 58.4 IU/ml (42.8-79.8) at 14 days post 1st boost dose with an interval of 28 days, and from 25.4 IU/ml (19.1-33.9) to 80.8 IU/ml (53·1-122·9) at an interval of 56 days (Figure 2A and 2B, Table S3). While, the neutralizing antibodies of participants receiving a dose of TIV showed no increase, which were significantly lower than that of those receiving a boost vaccination with ZF001 (p =0.0125; p =0.0005).

**Figure 2.**
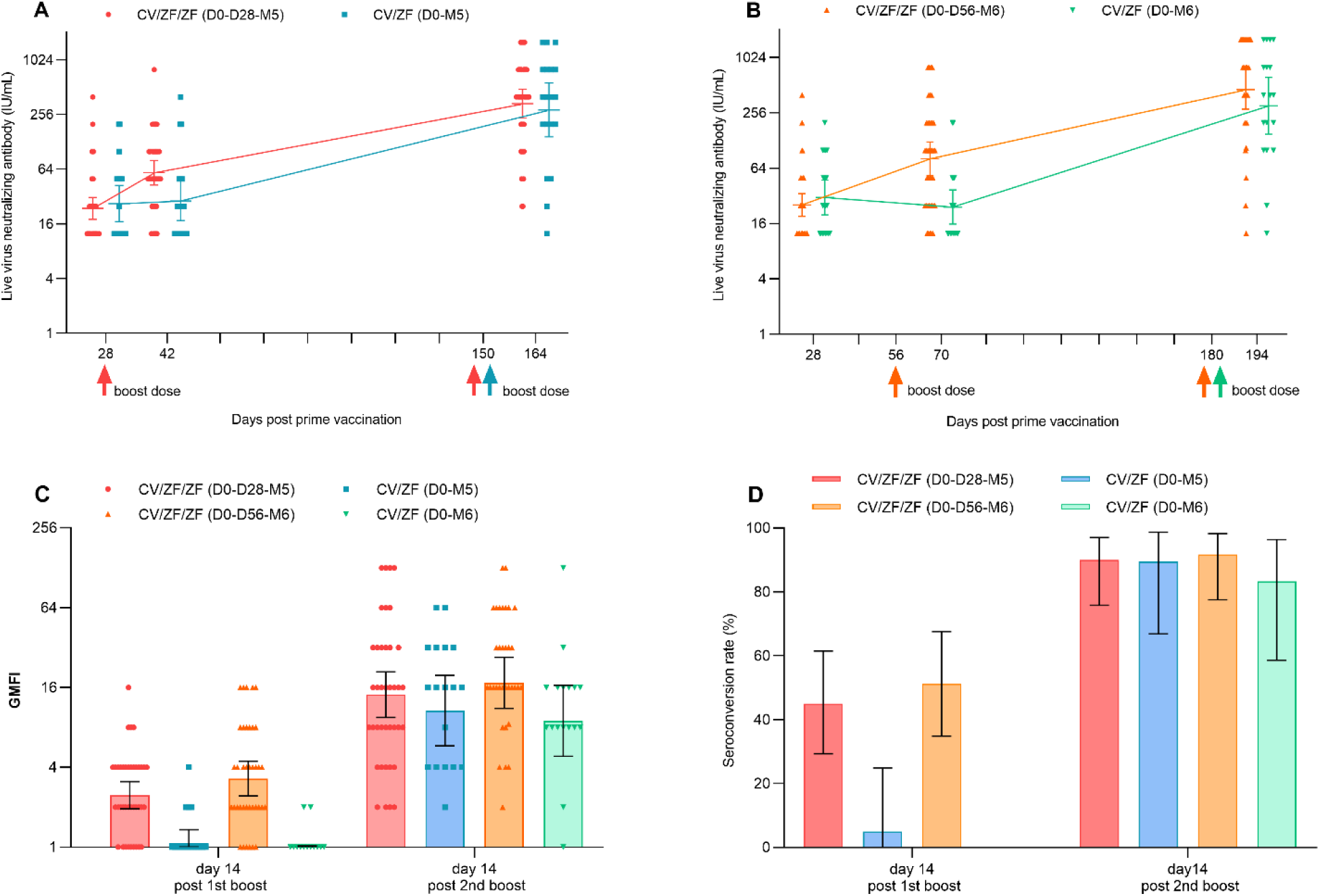
Live virus neutralizing antibodies against wild-type SARS-CoV-2 after prime and boost immunization. (**A** and **B**) GMTs of neutralizing antibodies to wild-type SARS-CoV-2 day 28 after prime vaccination, day 14 after 1st and 2nd boost dose in CV/ZF/ZF (D0-D28-M5) and CV/ZF (D0-M5) regimen (**A**) and CV/ZF/ZF (D0-D56-M6) and CV/ZF (D0-M6) regimen (**B**). (**C**) GMFI of neutralizing antibodies to wild-type SARS-CoV-2 day 14 after 1st and 2nd boost dose. (**D**) Seroconversion rate (%) of neutralizing antibodies to wild-type SARS-CoV-2 day 14 after 1st and 2nd boost dose. Neutralizing antibody (IU/ml) was converted to the WHO international standard (NIBSC code 20/136) using the following conversion factors: IU/ml=100 TCID_50_ ×3.125. IU/ml=International units per milliliter. TCID_50_=50% tissue culture infectious dose. Seroconversion was defined as at least a fourfold increase in the antibody titers at different time points after boost immunization compared to baseline level (at 28 days post prime dose). Horizontal bars show geometric mean or mean and error bas show 95% confidence interval. Up arrows represent the times of boost vaccination. GMT=geometric mean titer; GMFI=geometric mean fold increase.

Among participants receiving a dose of ZF001, the GMTs of neutralizing antibodies at 14 days post 2nd vaccination with ZF001 4 months apart increased to 334.9 IU/ml (95% CI 230.4, 486.9) in CV/ZF/ZF (D0-D28-M5), and 441.2 IU/ml (260.8, 746.4) in CV/ZF/ZF (D0-D56-M6) regimen group, with the geometric mean fold increases (GMFIs) of 14.1 and 17.3 compared with baseline, respectively (Figure 2A, 2B and 2C, Table S3). Among participants receiving a dose of TIV, the GMTs of neutralizing antibodies at 14 days post boost vaccination was 282.9 IU/ml (142.5, 561.8) in CV/ZF (D0-M5), and 293.9 IU/ml (137.6, 627.9) in CV/ZF (D0-M6) regimen group, with the GMFIs of 10.7 and 8.9, respectively (Figure 2A, 2B and 2C, Table S3). At 14 days post 2nd boost vaccination, the seroconversion of neutralizing antibody titer were observed in 90.0% of the participants in CV/ZF/ZF (D0-D28-M5), and 89.5% in CV/ZF (D0-M5), and 91.7% in CV/ZF/ZF (D0-D56-M6) and 83.3% in CV/ZF (D0-M6) regimen, respectively (Figure 2D, Table S3). Homologous immunization with Convidecia at “0-56 days” regimen and “0-6 months” regimen induced neutralizing antibodies with the GMTs of 100.0 IU/ml (95% CI 74.3-134.6) and 386.4 IU/ml (258.9, 576.7) at 28 days post boost dose, respectively, which were equivalent to those induced by heterologous boost immunisation with ZF001 (Figure S1). Neutralizing antibodies were numerically higher in participants aged 18-59 years than in those over 60 years (Figure 3).

**Figure 3.**
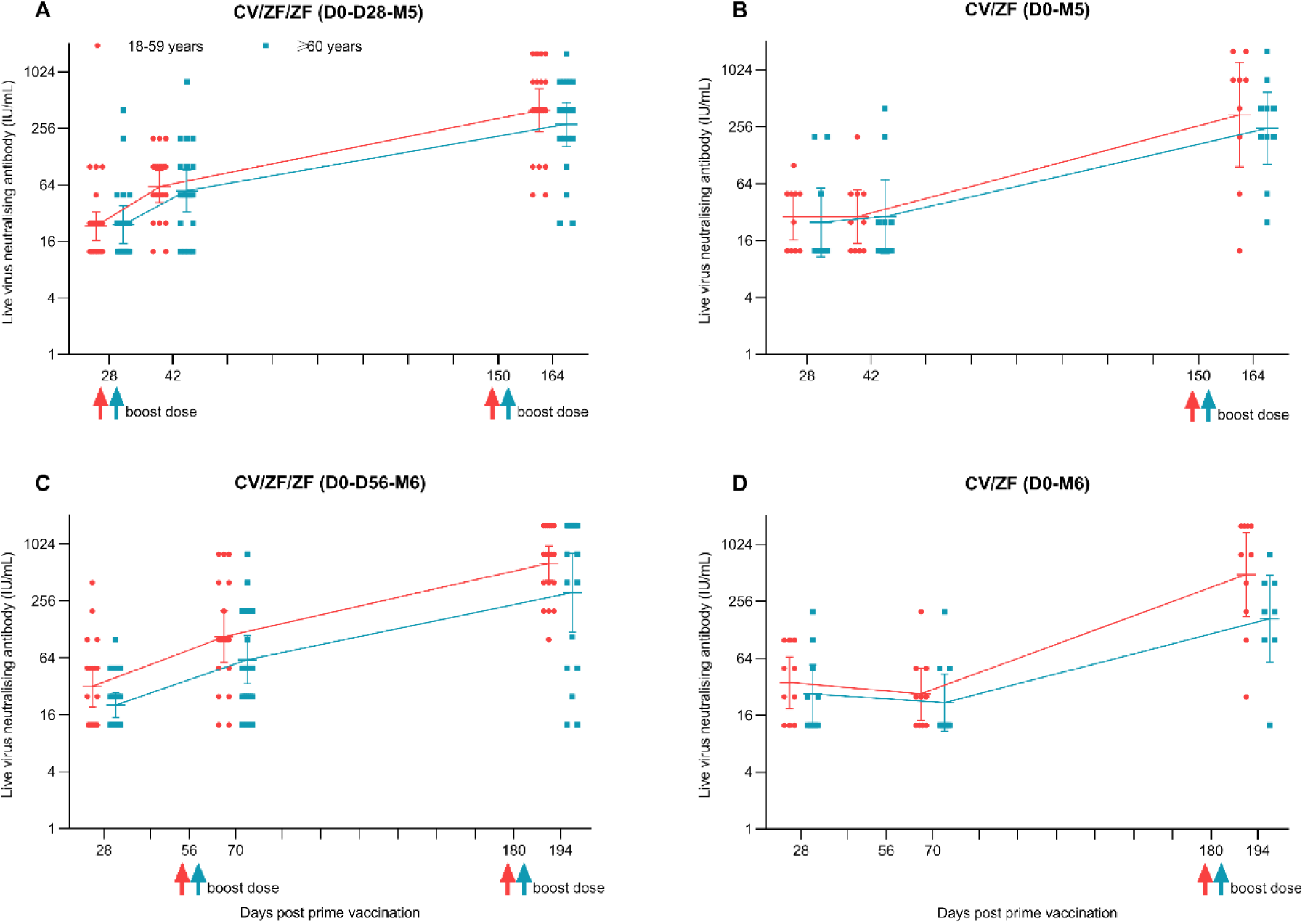
Live virus neutralizing antibodies to wild-type SARS-CoV-2 wild-type after prime and boost immunization according vaccination schedule, and age. Data presented are live virus neutralizing antibodies across four heterologous schedules at 28 days post prime vaccination with Convidecia, and at 14 days post 1st and 2nd boost vaccination with ZF001, according to age. Neutralizing antibody (IU/ml) was converted to the WHO international standard (NIBSC code 20/136) using the following conversion factors: IU/ml for Wuhan isolate=100 TCID_50_ ×3.125. IU/ml=International units per milliliter. TCID_50_=50% tissue culture infectious dose. Horizontal bars show geometric mean titre and error bas show 95% confidence interval. Up arrows represent the times of vaccination.

#### Anti-RBD and anti-spike IgG antibodies

In line with live virus neutralizing antibodies, for participants receiving a dose of either ZF001 or TIV, the 2nd boost immunization with ZF001 increased anti-RBD IgG to comparable level, with GMTs at 14 days post boost vaccination of 695.6 IU/ml (95% CI 465.9, 1038.5) in CV/ZF/ZF (D0-D28-M5) regimen, 514.7 IU/ml (255.9, 1035·2) in CF/ZF (D0 -M5) regimen, 951.4 IU/ml (594.0, 1523.9) in CF/ZF/ZF (D0-D56-M6) regimen and 534.5 IU/ml (256.7, 1112.9) in CF/ZF (D0-M6) regimen, respectively (Figure 4A and 4B, Table S4). Compared with anti-RBD IgG, the GMTs of anti-spike IgG were reduced according to point estimates, with the GMTs 14 days post 2nd boost of 571.9 IU/ml (95% CI 396.9, 823.9) in CV/ZF/Z F(D0-D28-M5) regimen, 412.9 IU/ml (202.1, 843.9) in CF/ZF (D0 -M5) regimen, 686.1 IU/ml (435.8, 1080.4) in CF/ZF/ZF IU/ml (D0-D56-M6) regimen and 407.3 IU/ml (211.4, 784.9) in CF/ZF (D0-M6) regimen, respectively (Figure 4C and 4D, Table S4).

**Figure 4.**
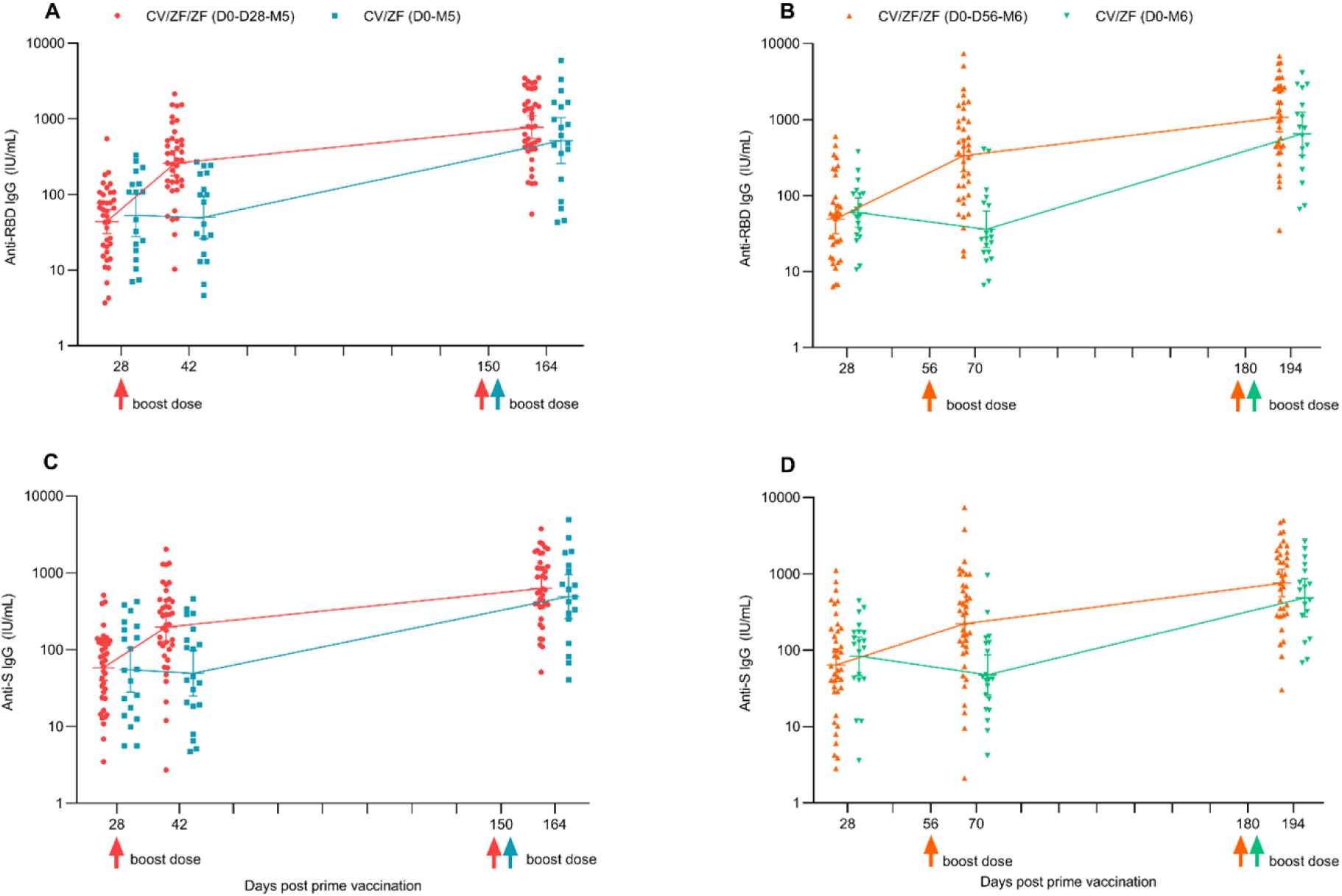
IgG binding antibodies after prime and boost immunization. (**A** and **B**) GMTs of anti-RBD IgG antibodies 28 after prime vaccination, day 14 after 1st and 2nd boost dose. (**C** and **D)** GMTs of anti-Spike IgG antibodies 28 after prime vaccination, day 14 after 1st and 2nd boost dose. IgG binding antibody (IU/ml) was converted to the WHO international standard using the following conversion formula: (IU/ml)=x*Dilution ratio. Before conversion, standard curves were constructed using the calibrator sample: y=0.0044*x+0.0841 for anti-RBD IgG antibodies, and y=0.0035*x+0.141 anti-S IgG antibodies (y=optical density (OD) value, x= the titer of calibrator sample). IU/ml=International units per milliliter. Horizontal bars show geometric mean titer (concentration) and error bas show 95% confidence interval. Up arrows represent the times of boost vaccination.

Similar patterns of humoral responses were found in all subgroup according to age, with binding antibodies to SARS-CoV-2 consistently higher in Convidecia/ZF001 regimen with an interval of 5 months or 6 months compared with an interval of 28 days or 56 days. Additionally, the younger adults had numerically higher humoral responses than did the older adults (Figure S2, Figure S3). Strong correlations were found between neutralizing antibodies and SARS-CoV-2 anti-RBD IgG, and neutralizing antibodies and SARS-CoV-2 anti-spike IgG at 14 days post 2nd boost dose (Pearson correlation coefficients of 0.87-0.92) (Figure S4).

#### Neutralizing antibody responses against the Delta variant

28 days after prime immunization with Convidecia, the GMTs of neutralizing antibody titers to B.1.617.2 variant was 2.8 (95%CI 2.5, 3.2). 14 days post 2nd boost vaccination, the GMTs of neutralizing antibody titers to B.1.617.2 variant was 38.0 (95% CI 26.7, 54.2) in CV/ZF/ZF (D0-D28-M5), 29·7 (15·3, 57·9) in CV/ZF (D0-M5), 41·9 (27·0, 65·0) in CV/ZF/ZF (D0-D56-M6) and 34.6 (18.1, 65.9) CV/ZF (D0-M6) regimen group, respectively (Figure 5C, Table S3). Similar with that after prime immunization with Convidecia, the GMTs ratio of neutralizing antibodies against Delta variant to wild-type elicited by boost vaccination ranged 0.29 and 0.35 across four heterologous regimens (Figure 5A and 5B). Compared with the prime immunization of Convidecia, however, the boost immunization with ZF001 at an interval of 5 months or 6 months induced higher neutralizing antibodies against B.1.617 variant, with the GMFIs of 10·6 to 14·4 (Table S3).

**Figure 5.**
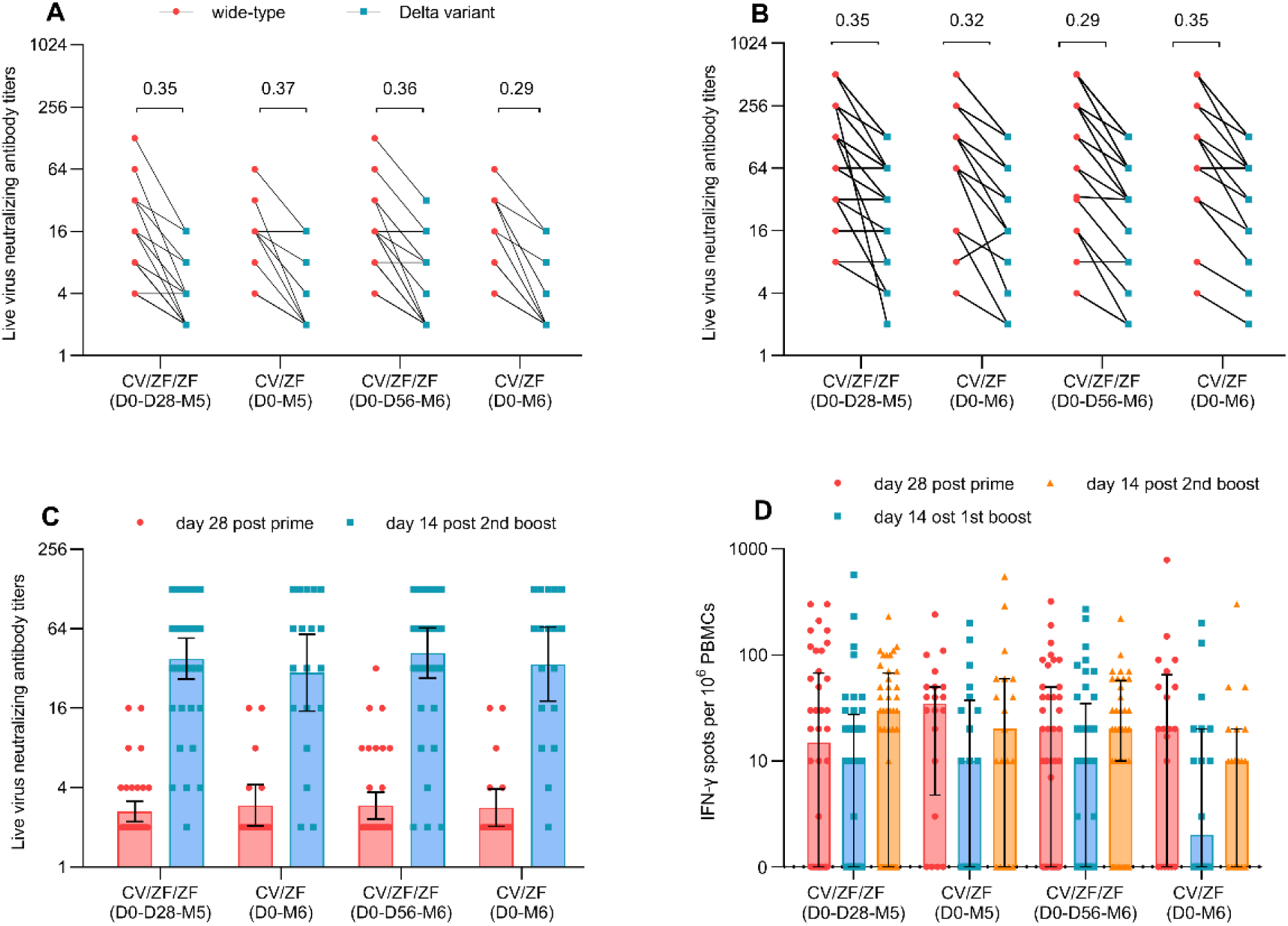
Live virus neutralizing antibody titers against Delta and specific T-cell response measured by ELISpot after prime and boost immunization. (**A** and **B**) Data above a short horizontal line indicates the geometric mean of neutralizing antibody titers against Delta to wild-type ratio 28 days post prime dose (A), and 14 days post 2nd boost dose (**B**). (**C**) GMTs of neutralizing antibody titers to Delta 28 days post prime dose and 14 days post 2nd boost dose. (D) Spot-forming cells with secretion of IFN-γ cytokines per 1×106 PBMCs 28 days post prime dose and 14 days post and 1st and 2nd boost dose. Horizontal bars show geometric mean titre and error bas show 95% confidence interval in panel (**C**). Horizontal bars show the median and error bars show the interquartile range in panel (**D**). ELISpot= enzyme-linked immunospot; IFN=interferon; PBMCs= peripheral blood mononuclear cells.

#### Vaccine-induced T cell responses

Ad5-vectored COVID-19 vaccine induced significant specific T -cell responses measured by enzyme-linked immunospot (ELISpot) assay. A median of 20·0 (interquartile range (IQR): 00·0, 57·5) spot-forming cells secreting IFN-γ per 1×10^6^ peripheral blood mononuclear cells (PBMCs) was observed at 28 days after prime vaccination, and no further increase was seen post boost immunization with ZF001 (Figure 5D). The median of IFN-γ spot counts per 10^6^ PBMCs 14 days after 1st boost was 10.0 (IQR: 0.0, 27.5) in CV/ZF/ZF (D0-D28-M5), 10.0 (IQR: 0.0, 37.5) in CV/ZF (D0-M5), 10.0 (IQR: 0.0, 25.0) in CV/ZF/ZF (D0-D56-M6) and 1.5 (IQR: 0.0, 20.0) in CV/ZF (D0-M6), respectively. The median of IFN-γ spot counts per 10^6^ PBMCs 14 days after 2nd boost was 10.0 (IQR: 0.0, 27.5) in CV/ZF/ZF (D0-D28-M5), 10.0 (IQR: 0.0, 37.5) in CV/ZF (D0-M5), 10.0 (IQR: 0.0, 25.0) in CV/ZF/ZF (D0-D56-M6) and 1.5 (IQR: 0.0, 20.0) in CV/ZF (D0-M6), respectively.

## Discussion

Our findings show that heterologous immunization of ZF001 in individuals whom were vaccinated with Convidecia is safe, showing the lower frequency of systemic adverse reactions was reported after boost dose with ZF001, compared with that after prime dose with Convidecia. Heterologous immunization of ZF001 with prime-boost intervals of 28 days or 56 days induced 2.5 and 3.3 folds higher humoral responses against SARS-CoV-2 than those induced by a single dose of Convidecia, respectively. In addition, we also found that heterologous vaccination with Convidecia and ZF001 at an interval of 5 months or 6 months following the one dose priming of Convidecia are more efficient in eliciting neutralizing antibodies, with GMFIs of 8.9 and 10.7 compared with a single dose of Convidecia, respectively.

In the present study, we founded that the impact of dose interval on the immune responses was greater than the number of doses, which may relate to memory B-cell maturation undergoing during 4-6 months (23). Notably, heterologous vaccination with Convidecia and ZF001 at an interval of 5 months or 6 months induced higher antibody titers than that elicited by immunization 28 days or 56 days apart. Additionally, two-dose schedule with D0-M5 and D0-M6 induced comparable antibody level comparable with that elicited by three doses of heterologous immunization with D0-D28-M5 and D0-D56-M6 schedules. Zhao et al (24) also showed that prolonged-interval ZF2001 (receiving three doses of ZF2001 at interval of month 0, 1 and 4, M0-M1-M4) induced higher binding and neutralizing antibodies than the short-interval ZF2001 (M0-M1-M2), including against SARS-CoV-2 prototype strain and variants of concern such as Delta and Omicron. These findings support the use of a prolonged booster interval to elicit stronger immune responses in persons who had previously received prime immunization of COVID-19 vaccine.

As we known, pre-existing anti-adenovirus immunity is the biggest obstacle for the adenovirus-vectored vaccines to overcome, especially for Ad5 eliciting widespread pre-existing immunity in the human population. In the previous phase IIb trial of Convidecia, the boosting effect of homologous prime-boost regime apart 56 days on immune responses was limited due to high anti-Ad5 antibodies(19). In order to minimize the negative effect of pre-existing anti-Ad5 antibody, heterologous prime-boost regimens and a wider prime-boost interval are necessary to provide enhance of immune responses. Our study indicated that homologous Convidecia vaccination apart 6 months induced comparable antibodies with that elicited by heterologous Convidecia-ZF001 immunization at an interval of 5 months or 6 months. The impact of dose interval was also be observed in other vectored COVID-19 vaccines. There is better immunogenicity when a second dose of Ad26 is given at 6 months after the first dose of Ad26 compared with 2 months(25). ChAdOx1 nCoV-19 has also shown that a longer prime-boost interval (≥12 weeks) provided higher protective efficacy than a short interval (>6 weeks)(26).

The neutralizing antibodies against Delta variant elicited by heterologous immunization with Convidecia and ZF001 decreased about 3-4 folds relative to wild-type across the four different regimens, and which was similar with that after the prime immunization. Nevertheless, heterologous schedules maintained higher neutralizing antibodies against Delta variant than prime vaccination. However, the use of ZF001 as a boost dose not increases the cellular immunity responses obtained after the initial dose of Convidecia, which was in line with that reported in a previous trial with ZF2001 booster at interval of 4-8 months following two-dose inactivated vaccines(27). Compared with protein-subunit-based vaccines containing aluminium adjuvants, those with novel adjuvants could induce stronger immune responses. The results of Com-COV2 study showed that heterologous immunization with ChAdOx1 nCoV-19 vaccine and NVXCoV2373 (a Matrix-M adjuvanted recombinant spike protein vaccine) in a interval of 8-12 weeks induced both humoral and T-cell immune responses superior to that homologous ChAdOx1 nCoV-19 vaccine(16).

Data from the phase 3 efficacy trial showed a single dose of Convidecia could provide 57·5% protective efficacy against symptomatic COVID-19 at 28 days or more post-vaccination and 91·7% vaccine efficacy against severe disease(28). The preliminary efficacy of ZF001 indicated that the three-dose schedule (30 days apart) could provide 81·7% efficacy against symptomatic COVID-19 and 100% efficacy against severe disease. In addition, vaccine efficacy against Alpha and Delta variants was 92·9% and 77·5%, respectively(29). Our findings indicate that the heterologous schedule of Convidecia followed by a boost dose of ZF001 with 5-6 months interval increased neutralizing antibodies by 9-17 folds, compared with that after an initial dose of Convidecia. Given the established associations between neutralizing antibody titers and vaccine efficacy(4, 30), heterologous immunization with Convidecia and ZF001 5-6 months apart are also likely to be highly effective, and could be considered in some circumstances for national vaccine programmers.

This study has several limitations. First, it is the absence of a randomized control group completing the homologous Convidecia scheme. Although we select two extend controls receiving homologous immunization of Convidecia following a 0-56 days regimen and 0-6 months regimen, which are both comparable with the cohorts receiving heterologous immunization between Convidecia and ZF001 in baseline characteristics, there may be some potential bias. As an immunogenicity and reactogenicity study, we do not know whether the immune responses observed in our study will result in better effectiveness, and it is needed to be confirmed in real-world studies. Additionally, we are unable, at this point, to determine whether higher antibody titers measured at 14 days post boost immunization will result in a more sustained elevation of antibodies, and this will be assessed at 6 moths post 2nd boost vaccination. Lastly, the recently emerged SARS-CoV-2 Omicron variants of concern is quickly rising in worldwide and raised concerns about the effectiveness of available vaccines due to multiple amino acid mutations in the spike protein(31). Preliminary studies indicated that the neutralizing activity of plasma from individuals receiving prime COVID-19 vaccination from different platforms is severely reduced against Omicron variant(32, 33). In this study, the neutralizing activity of heterologous immunization with Convidecia and ZF001 against Omicron is not tested.

In conclusion, our study shows that heterologous schedules of ZF001 following the primary vaccination of Convidecia are safe and can induce significant humoral immunity, particularly with a 5-6 months prime-boost interval. These results support flexibility in cooperating viral vectored vaccines and recombinant protein vaccine, subject to supply and logistical considerations, especially for vaccines being deployed in low-income and middle-income countries.

## Data Availability

All relevant data are within the manuscript and its Supporting Information files. Individual participant data will be available for request 1 month after the completion of the study (anticipated in April 2022), upon requests directed to the corresponding author

## Author Contributions

JXL is the principal investigator of this trial. FCZ, JXL, PFJ and WC contributed to the protocol and design of the study. XLG led the laboratory analyses. FCZ, LPD and GFG contributed to critical review revising of the report. JXL and PFJ contributed to the data interpretation and revising of this manuscript. PD, YC, FJS, JXL and XYX contributed to the laboratory tests. SHM and LLW led and participated in the site work, including the recruitment, follow-up and data collection. YNZ and CCC contributed to study supervision. LRJ and JLF was responsible for statistical analysis and have verified the underlying data. PFJ and JXL drafted of the manuscript. All authors reviewed and approved the final report. All authors had full access to all the data in the study and had final responsibility for the decision to submit for publication.

## Conflict-of-interest statement

JXL reports grants from National Natural Science Foundation of China (grant 82173584). FCZ reports grants from Jiangsu Provincial Key Research and Development Program grant (BE2021738). WC, YNZ and CCC are the employees of Anhui Zhifei Longcom Biopharmaceutical. All other authors declare no competing interest.

## Acknowledgments

We thank all study participants enrolled in this trial. We gratefully acknowledge the participation and support of many staff in this study. The work was funded by National Natural Science Foundation of China (grant 82173584), Jiangsu Provincial Key Research and Development Program grant (BE2021738), and Anhui Zhifei Longcom Biopharmaceutical Co., Ltd.

## Data availability

The study protocol is provided in the Supplementary Materials. Researchers who provide a scientifically sound proposal will be allowed to access to the de-identified individual participant data. Individual participant data will be available for request 1 month after the completion of the study (anticipated in April 2022), upon requests directed to the corresponding author; after approval of a proposal, data can be shared through a secure online platform.

## Supplementary materials

**Figure 1.**
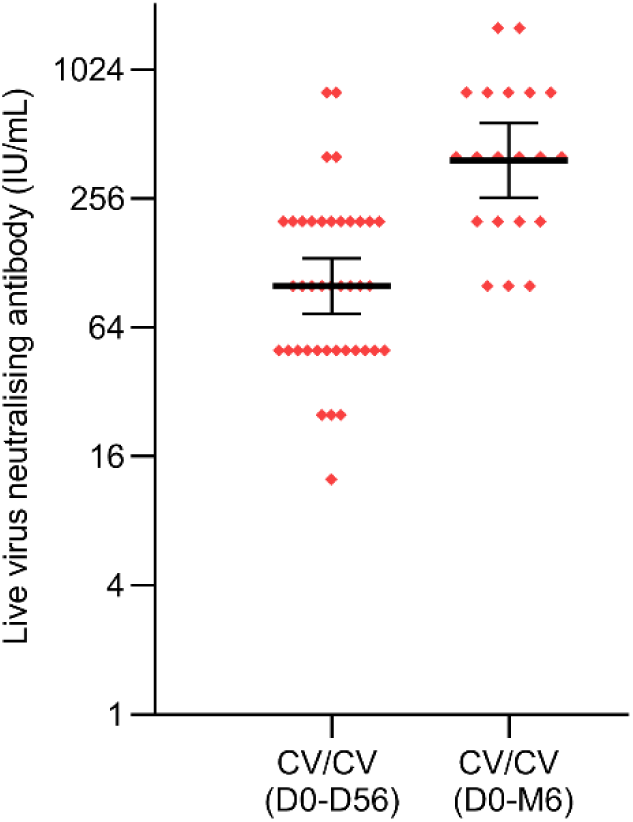
Live virus neutralizing antibodies to wild-type SARS-CoV-2 day 28 after homologous boost immunization from external cohort. Neutralizing antibody (IU/ml) was converted to the WHO international standard (NIBSC code 20/136) using the following conversion factors: IU/ml=100 TCID_50_ ×3.125. IU/ml=International units per milliliter. TCID_50_=50% tissue culture infectious dose. Horizontal bars show geometric mean and error bas show 95% confidence interval. Up arrows represent the times of boost vaccination. CV/CV (D0-D56)=receiving Convidecia/Convidecia at day 0 and day 56. CV/CV (D0-M6)=receiving Convidecia/Convidecia at day 0 and month 6.

**Figure S2.**
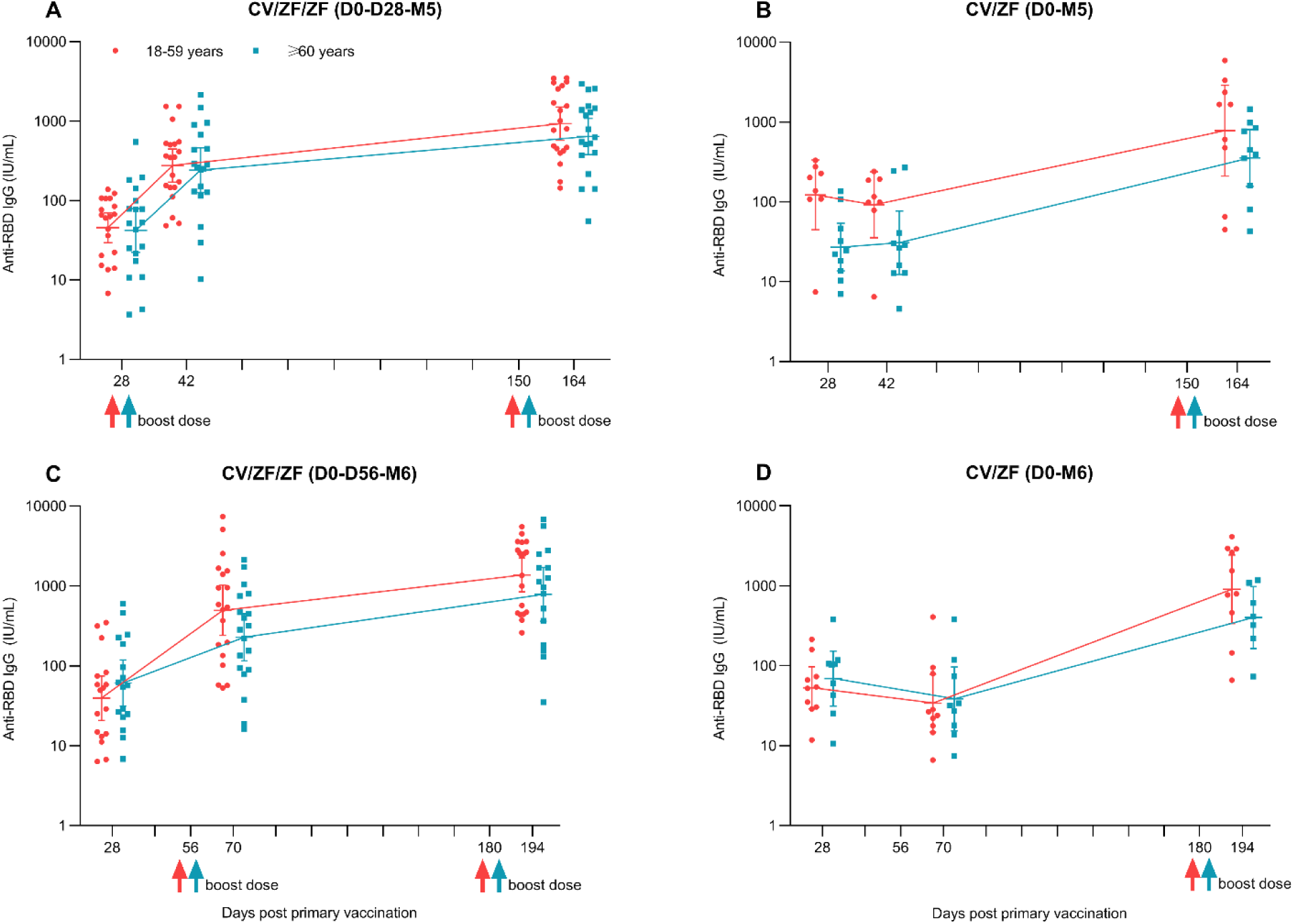
SARS-CoV-2 anti-RBD IgG antibodies after prime and boost immunization according vaccination schedule, and age. ELISA IgG binding antibody (IU/ml) was converted to the WHO international standard using the following conversion formula: (IU/ml)=x* Dilution ratio. Before conversion, standard curves were constructed using the calibrator sample: y=0.0044*x+0.0841 (y=optical density (OD) value, x= the titre of calibrator sample). Horizontal bars show geometric mean concentration and error bas show 95% confidence interval. Up arrows represent the boost vaccination with ZF001. IU/ml=International units per milliliter. CV/ZF/ZF (D0-D28-M5)=receiving Convidecia/ZF001/ZF001 at day 0, day 28 and month 5; CV/ZF (D0-M5)=receiving Convidecia/ZF001 at day 0 and month 5; CV/ZF/ZF (D0-D56-M6)=receiving Convidecia/ZF001/ZF001 at day 0, day 56 and month 6; CV/ZF (D0-M6)=receiving Convidecia/ZF001 at day 0 and month 6.

**Figure S3.**
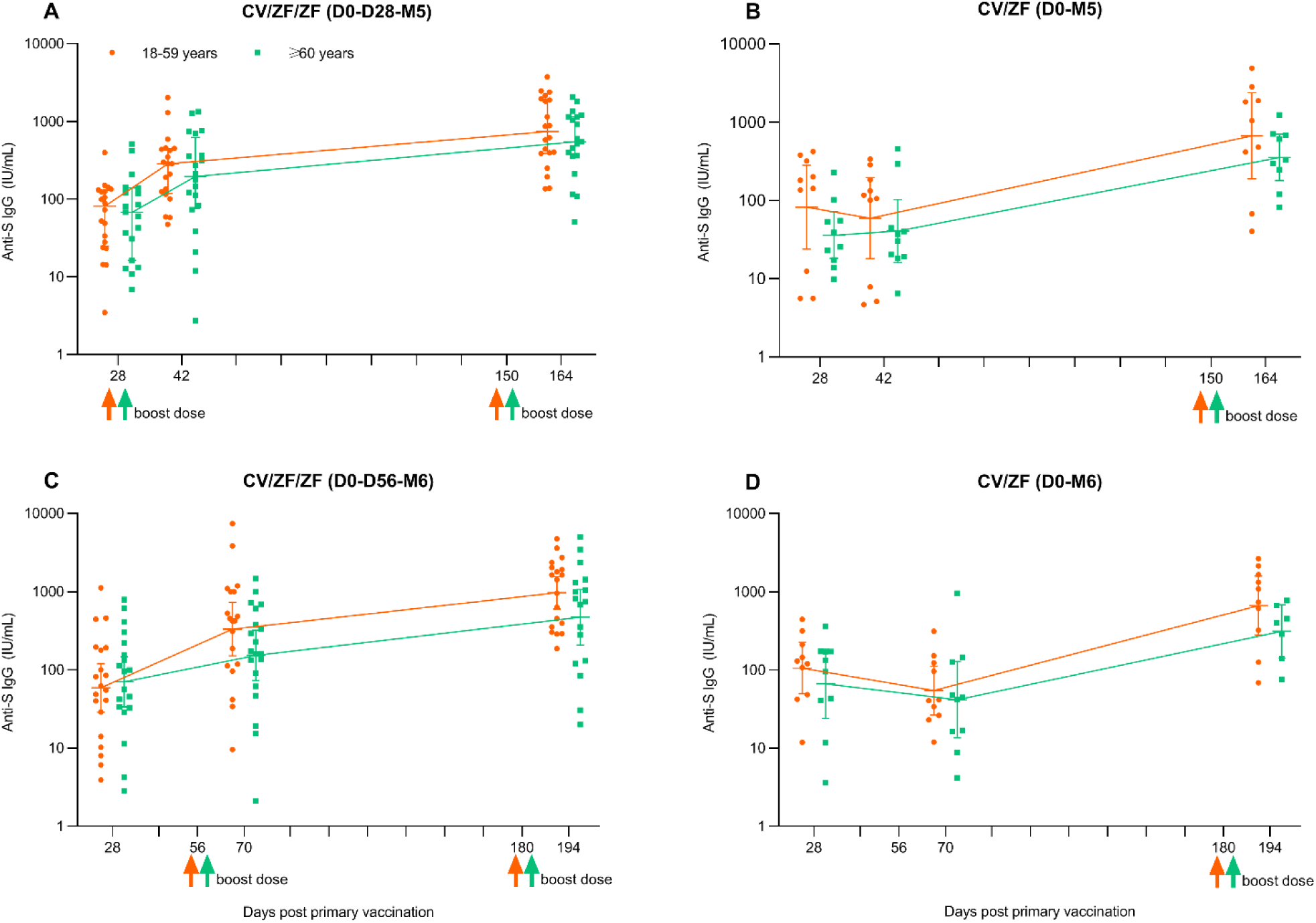
SARS-CoV-2 anti-S IgG antibodies after prime and boost immunization according vaccination schedule, and age. ELISA IgG binding antibody (IU/ml) was converted to the WHO international standard using the following conversion formula: (IU/ml)=x* Dilution ratio. Before conversion, standard curves were constructed using the calibrator sample: y=0.0035*x+0.141 (y=optical density (OD) value, x= the titre of calibrator sample). Horizontal bars show geometric mean concentration and error bas show 95% confidence interval. Up arrows represent the boost vaccination with ZF001. IU/ml=International units per milliliter. CV/ZF/ZF (D0-D28-M5)=receiving Convidecia/ZF001/ZF001 at day 0, day 28 and month 5; CV/ZF (D0-M5)=receiving Convidecia/ZF001 at day 0 and month 5; CV/ZF/ZF (D0-D56-M6)=receiving Convidecia/ZF001/ZF001 at day 0, day 56 and month 6; CV/ZF (D0-M6)=receiving Convidecia/ZF001 at day 0 and month 6.

**Figure S4.**
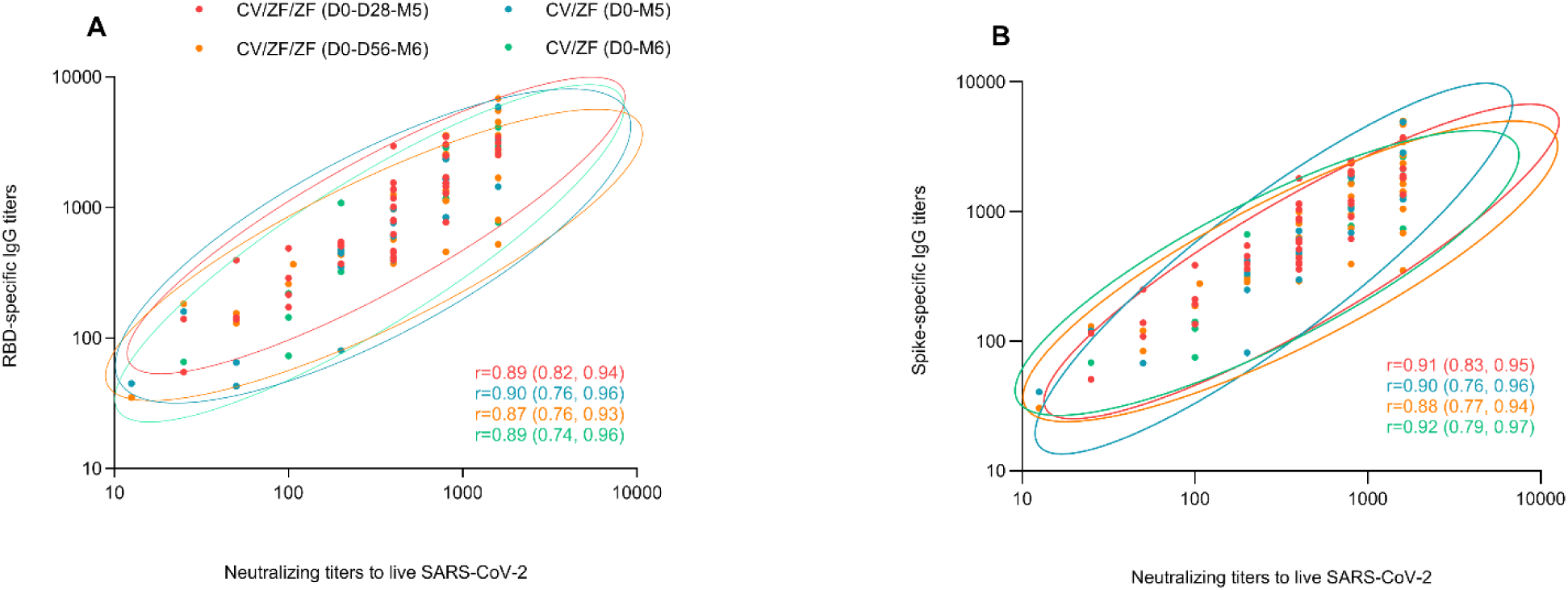
Correlations between immune response by vaccination schedules. Correlations at 14 days post 2nd boost vaccination were analysed between neutralizing titers to live SARS-CoV-2 wide-type virus and RBD-specific IgG titers (A), between neutralizing titers to live SARS-CoV-2 wide-type virus and spike-specific IgG titers (B). Ellipses show the 95% CIs for different vaccine schedules, assuming multivariate normal distributions. Pearson correlation coefficients (95% CIs) are presented for each vaccine schedule. CV/ZF/ZF (D0-D28-M5)=receiving Convidecia/ZF001/ZF001 at day 0, day 28 and month 5; CV/ZF (D0-M5)=receiving Convidecia/ZF001 at day 0 and month 5; CV/ZF/ZF (D0-D56-M6)=receiving Convidecia/ZF001/ZF001 at day 0, day 56 and month 6; CV/ZF (D0-M6)=receiving Convidecia/ZF001 at day 0 and month 6.

**Table S1.**
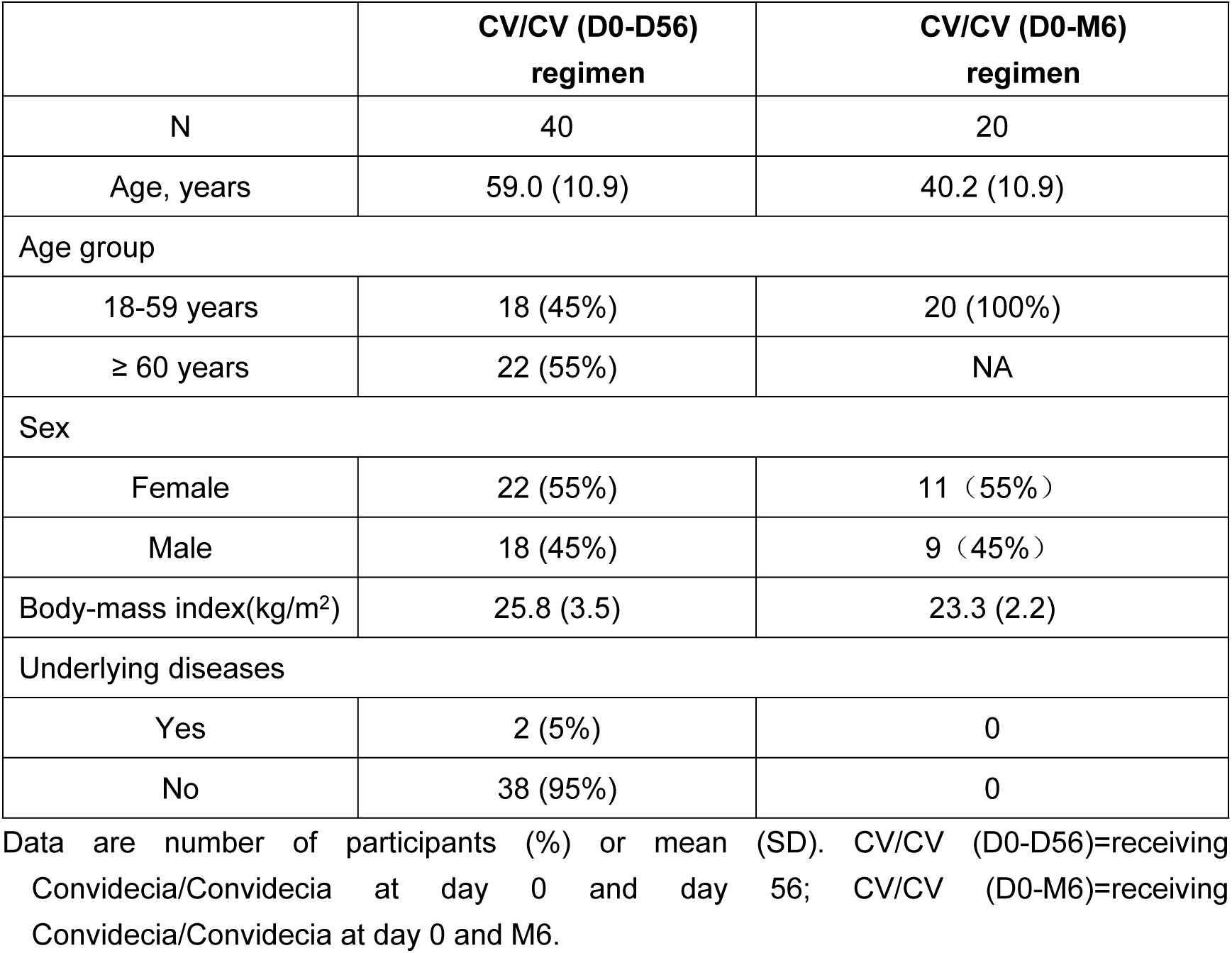
Baseline characteristics of the participants from external comparators

**Table S2.**
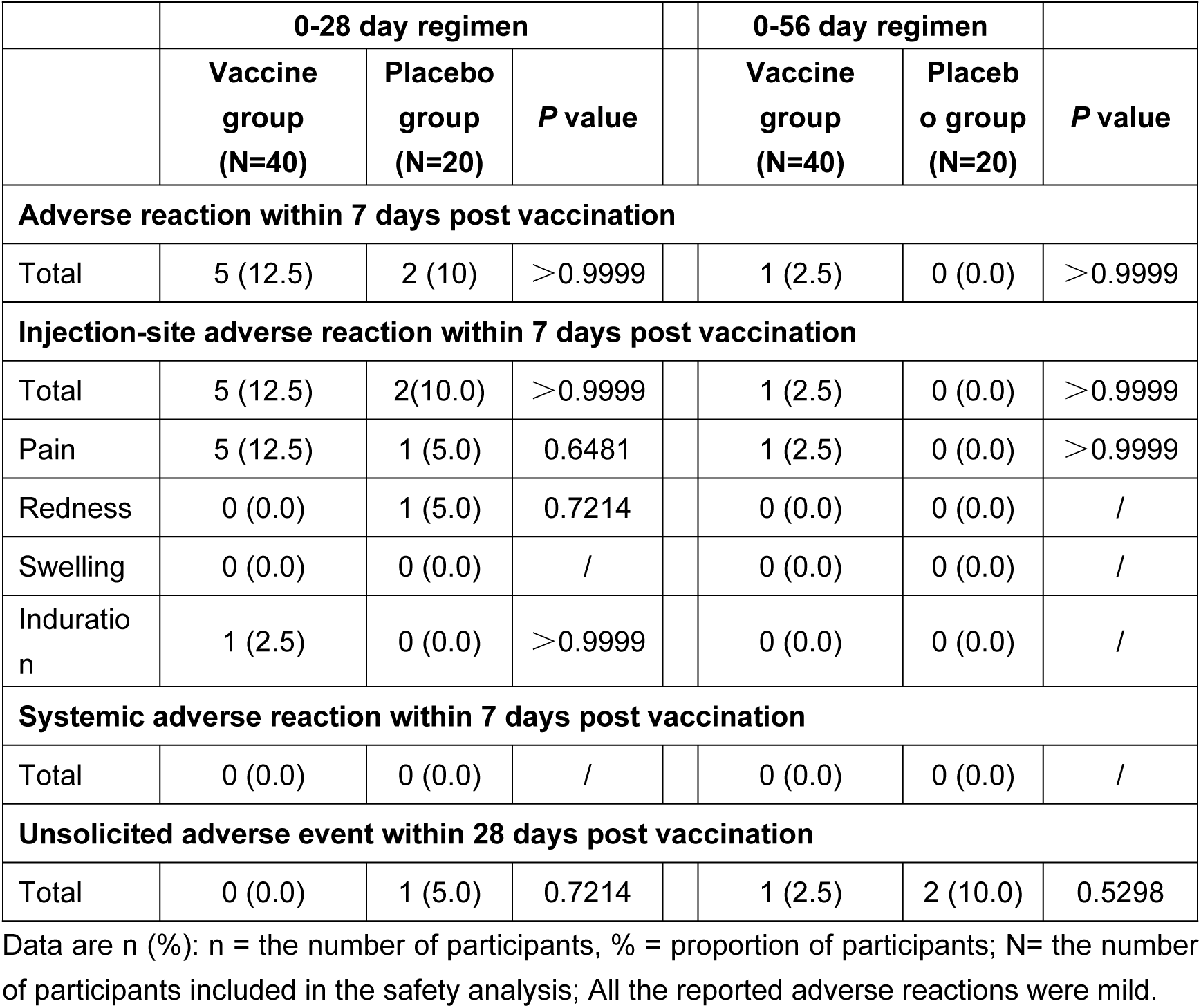
Adverse reactions occurred within 7 days and unsolicited adverse events within 28 days post 1st booster vaccination.

**Table S3.** Live virus neutralizing antibodies after post prime and boost dose.

|  | **CV/ZF/ZF**<br>**(D0-D28-M5)**<br>**regimen** | **CV/ZF**<br>**(D0-M5)**<br>**regimen** | **P value** |  | **CV/ZF/ZF**<br>**(D0-D56-M6)**<br>**regimen** | **CV/ZF**<br>**(D0-M6)**<br>**regimen** | **P value** |
| --- | --- | --- | --- | --- | --- | --- | --- |
| **Live virus neutralizing antibodies against wild-type isolate** |  |  |  |  |  |  |  |
| **Day 28 after priming** |  |  |  |  |  |  |  |
| N | 40 | 20 |  |  | 40 | 20 |  |
| GMT | 7.6<br>(5.8, 10.028) | 8.6<br>(5.4, 13.6) | 0.6259 |  | 8.1<br>(6.1, 10.8) | 9.9<br>(6.4, 15.2) | 0.4457 |
| GMT(IU/mL) | 23.7<br>(18.0, 31.3) | 26.8<br>(17.0, 42.3) | 0.6259 |  | 25.4<br>(19.1, 33.9) | 30.8<br>(19.9, 47.6) | 0.4457 |
| **Day 56 after priming** |  |  |  |  |  |  |  |
| N | NA | NA | NA |  | 40 | 20 |  |
| GMT | NA | NA | NA |  | 6.5<br>(4.9, 8.5) | 8.3<br>(4.9, 13.9) | 0.3469 |
| GMT (IU/mL) | NA | NA | NA |  | 20.3<br>(15.6, 26.5) | 25.9<br>(15.4, 43.6) | 0.3469 |
| GMFI | NA | NA | NA |  | 0.8<br>(0.6, 1.0) | 0.8<br>(0.6, 1.2) | 0.7929 |
| Seroconversion<br>rate(%) | NA | NA | NA |  | 2.5<br>(0.1, 13.2) | 5.0<br>(0.1, 24.9) | 1.0000 |
| **Day 14 post-1st boosting** |  |  |  |  |  |  |  |
| N | 40 | 20 |  |  | 39 | 20 |  |
| GMT | 18.7<br>(13.7, 25.5) | 9.2<br>(5.6, 15.2) | 0.0125 |  | 25.9<br>(17.0, 39.3) | 7.7<br>(5.0, 11.9) | 0.0005 |
| GMT (IU/mL) | 58.4<br>(42.8, 79.8) | 28.7<br>(17.4, 47.4) | 0.0125 |  | 80.8<br>(53.1, 122.9) | 24.2<br>(15.8, 37.0) | 0.0005 |
| GMFI | 2.5<br>(1.9, 3.1) | 1.1<br>(0.9, 1.4) | <0.0001 |  | 3.3<br>(2.4, 4.4) | 0.8<br>(0.6, 1.0) | <0.0001 |
| Seroconversion<br>rate(%) | 45.0<br>(29.3, 61.5) | 5.0<br>(0.1, 24.9) | 0.0017 |  | 51.3<br>(34.8, 67.6) | 0.00(-) | <0.0001 |
| **Day 28 post-1st boosting** |  |  |  |  |  |  |  |
| N | 40 | 20 |  |  | 39 | 20 |  |
| GMT | 11.9<br>(8.9, 15.9) | 6.1<br>(4.4, 8.4) | 0.0046 |  | 22.0<br>(14.7, 33.1) | 6.9<br>(4.2, 11.5) | 0.0009 |
| GMT (IU/mL) | 37.3<br>(27.9, 49.7) | 18.9<br>(13.7, 26.2) | 0.0046 |  | 68.8<br>(45.9, 103.3) | 21.8<br>(13.2, 35.9) | 0.0009 |
| GMFI | 1.6<br>(1.3, 1.9) | 0.7<br>(0.5, 0.9) | <0.0001 |  | 2.8<br>(2.1, 3.7) | 0.7<br>(0.5, 1.0) | <0.0001 |
| Seroconversion<br>rate(%) | 12.5<br>(4.2, 26.8) | 0.00(-) | 0.1588 |  | 56.4<br>(39.6, 72.2) | 5.0<br>(0.1, 24.9) | 0.0001 |

| Day 14 post-2nd boosting |  |  |  |  |  |  |  |
| --- | --- | --- | --- | --- | --- | --- | --- |
| N | 40 | 19 |  |  | 36 | 18 |  |
| GMT | 107.2<br>(73.7, 155.8) | 90.5<br>(45.6, 179.8) | 0.6298 |  | 141.2<br>(83.4, 238.8) | 94.1<br>(44.0, 200.9) | 0.3669 |
| GMT (IU/mL) | 334.9<br>(230.3, 486.9) | 282.8<br>(142.4, 561.8) | 0.6298 |  | 441.2<br>(260.8, 746.4) | 293.9<br>(137.6, 627.9) | 0.3669 |
| GMFI | 14.1<br>(9.5, 20.9) | 10.7<br>(5.8, 19.8) | 0.4271 |  | 17.3<br>(11.1, 26.9) | 8.9<br>(4.8, 16.7) | 0.0837 |
| Seroconversion rate (%) | 90.0<br>(75.8, 97.1) | 89.5<br>(66.9, 98.7) | 1.0000 |  | 91.7<br>(77.5, 98.3) | 83.3<br>(58.6, 96.4) | 0.3883 |
| Live virus neutralizing antibodies against Delta variant B.1.617.2 |  |  |  |  |  |  |  |
| Day 28 after priming |  |  |  |  |  |  |  |
| N | 40 | 20 |  |  | 40 | 20 |  |
| GMT | 2.6 (2.2, 3.2) | 2.9 (2.1, 4.2) | 0.5391 |  | 2.9 (2.3, 3.7) | 2.8 (2.0, 3.9) | 0.8614 |
| Delta to wild-type ratio | 0.35<br>(0.29, 0.42) | 0.37<br>(0.27, 0.50) | 0.7456 |  | 0.36<br>(0.30, 0.43) | 0.29<br>(0.21, 0.39) | 0.1743 |
| Day 14 post-2nd boosting |  |  |  |  |  |  |  |
| N | 40 | 19 |  |  | 36 | 18 |  |
| GMT | 38.0<br>(26.7, 54.2) | 29.7<br>(15.3, 57.9) | 0.4650 |  | 41.9<br>(27.0, 65.0) | 34.6<br>(18.1, 65.9) | 0.6095 |
| GMFI | 14.4<br>(10.1, 20.6) | 10.6<br>(5.6, 20.3) | 0.3684 |  | 14.3<br>(9.5, 21.3) | 11.8<br>(6.3, 21.8) | 0.5826 |
| Delta to wild-type ratio | 0.35<br>(0.27, 0.47) | 0.32<br>(0.23, 0.46) | 0.6994 |  | 0.29<br>(0.24, 0.35) | 0.35<br>(0.26, 0.48) | 0.2044 |

Data shown are geometric mean (95% CI) for continuous variables, and n (%; 95%CI) for binary variables. \* Neutralizing antibody (IU/ml) was converted to the WHO international standard (NIBSC code 20/136) using the following conversion factors: IU/ml for wild-type isolate=100 TCID<sub>50</sub> ×3.125. IU/ml=International units per milliliter, TCID<sub>50</sub>=50% tissue culture infectious dose. Seroconversion was defined as at least a fourfold increase in the antibody titers at different time points after boost immunisation compared to baseline level (at 28 days post prime dose). GMT=geometric mean titer; GMFI=geometric mean fold increase. NA=Not Applicable. CV/ZF/ZF (D0-D28-M5)=receiving Convidecia/ZF001/ZF001 at day 0, day 28 and month 5; CV/ZF (D0-M5)=receiving Convidecia/ZF001 at day 0 and month 5; CV/ZF/ZF (D0-D56-M6)=receiving Convidecia/ZF001/ZF001 at day 0, day 56 and month 6; CV/ZF (D0-M6)=receiving Convidecia/ZF001 at day 0 and month 6.

**Table S4.**
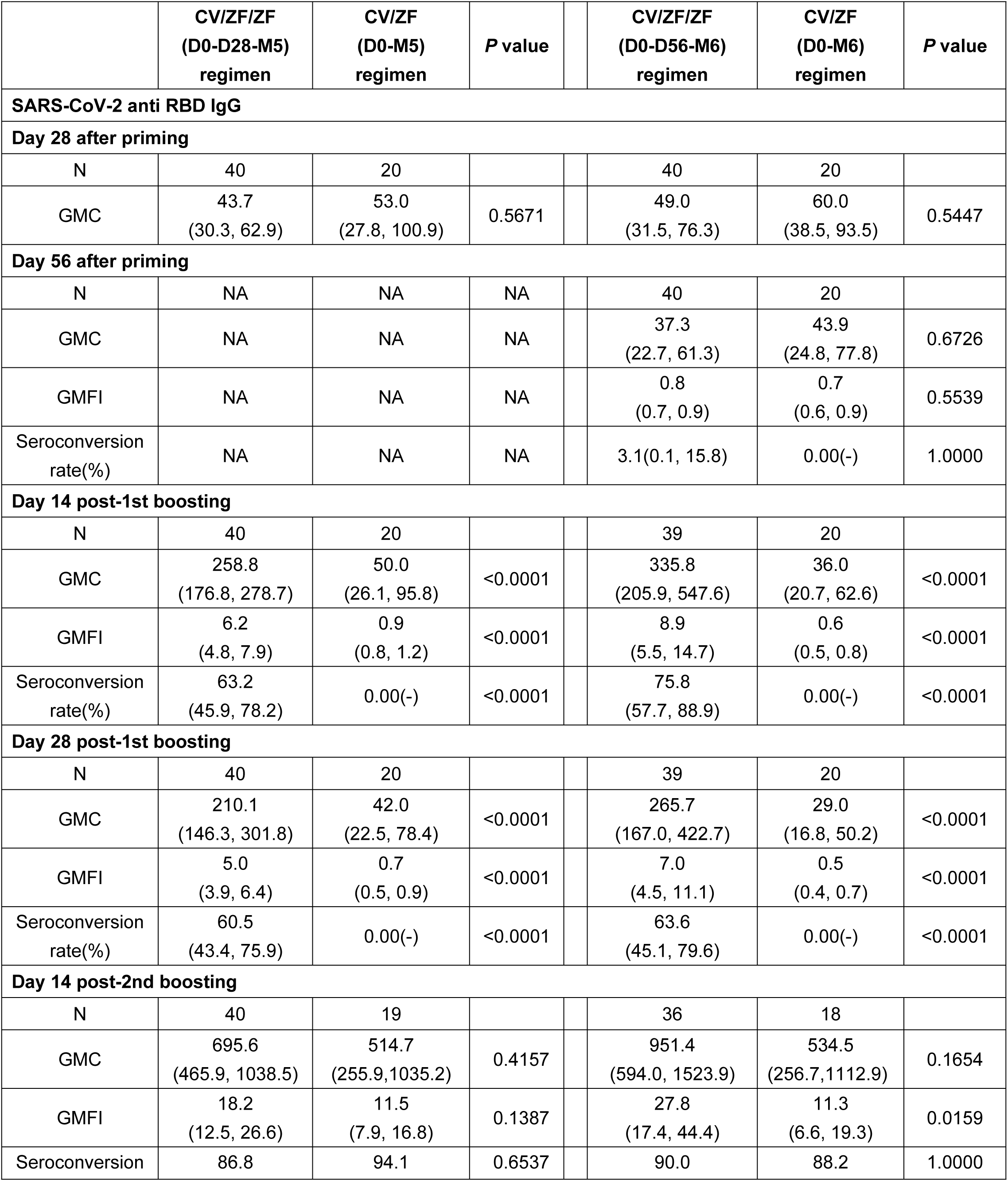
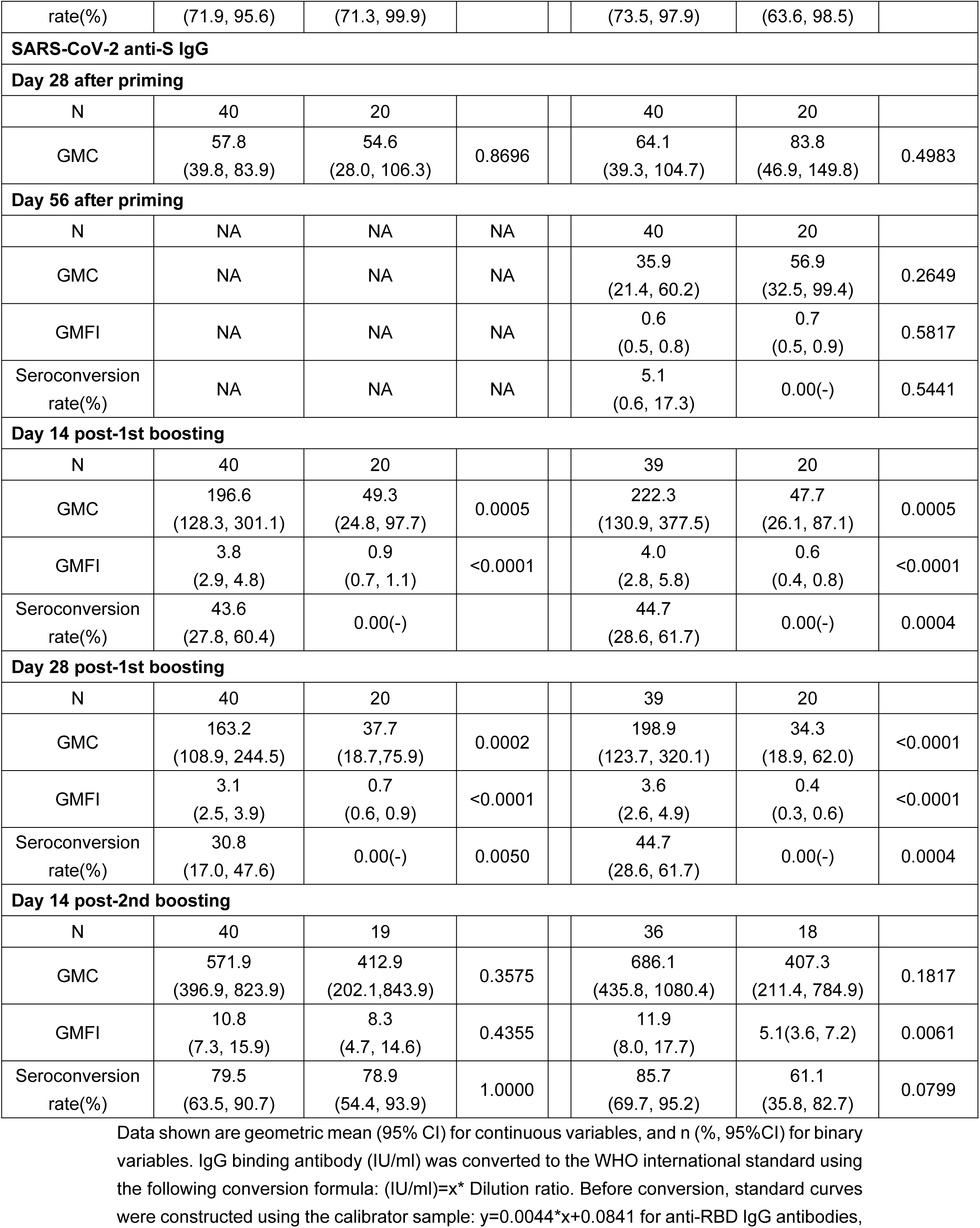
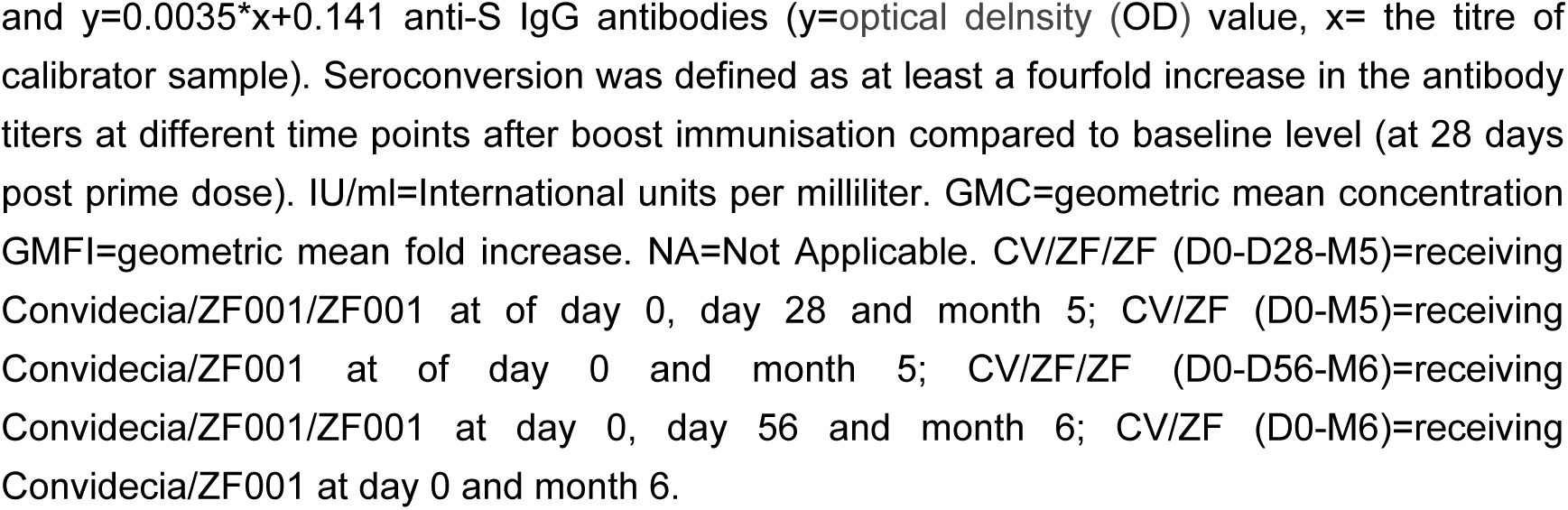
SARS-CoV-2 anti-RBD IgG and anti-S IgG antibodies after prime and boost dose.

**Table S5.**
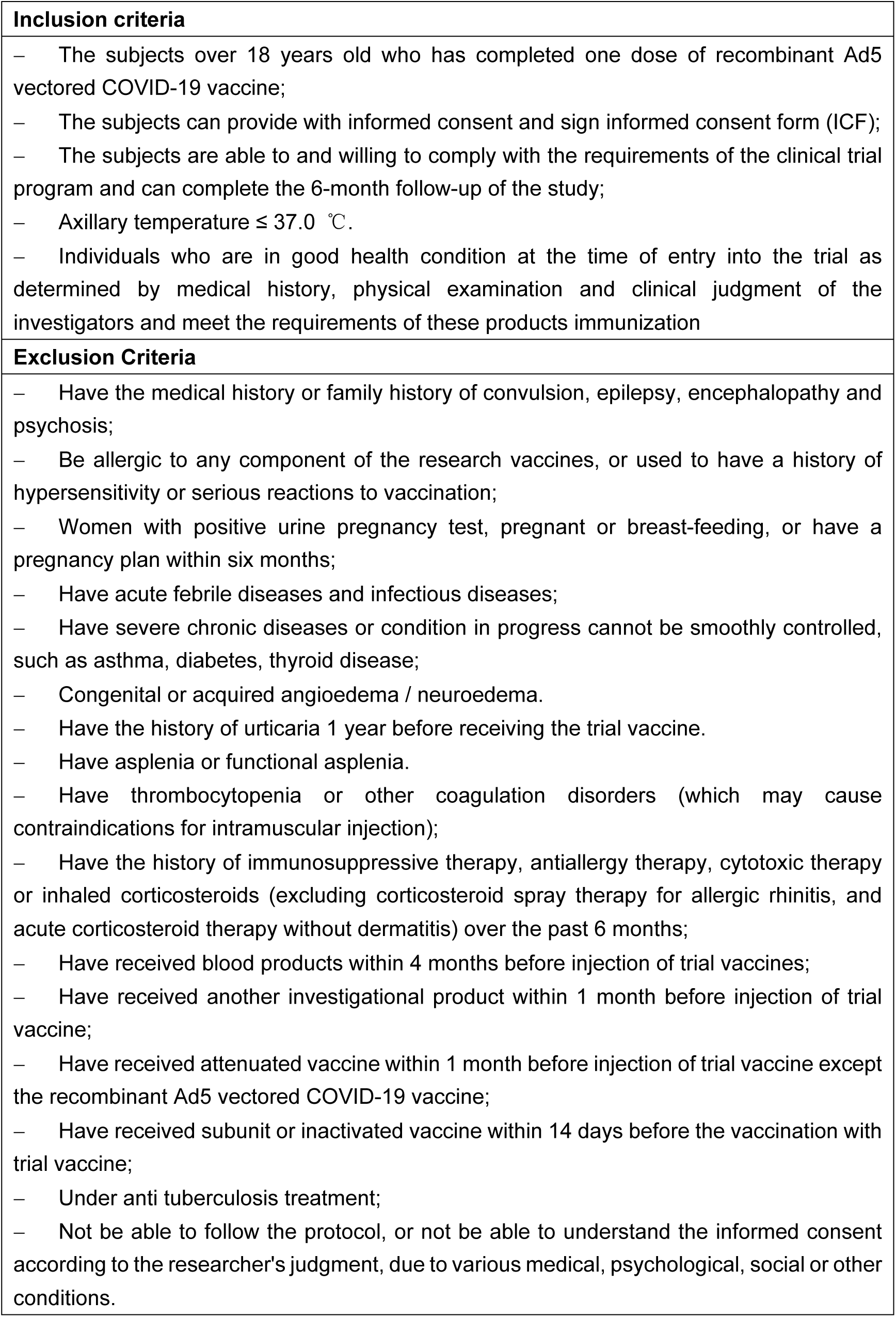
The inclusion and exclusion criteria Inclusion criteria

## CONSORT 2010 checklist of information to include when reporting a randomised trial*

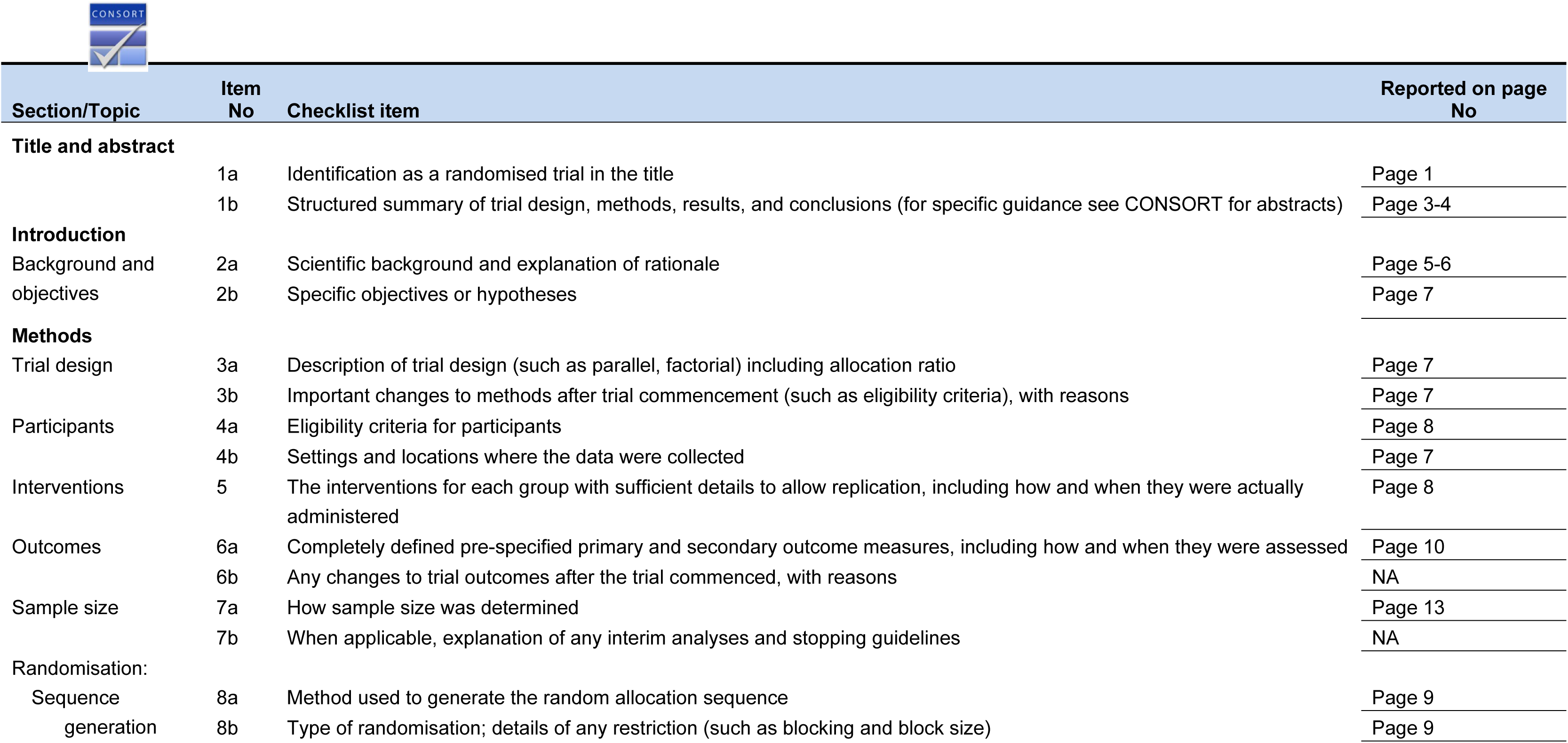
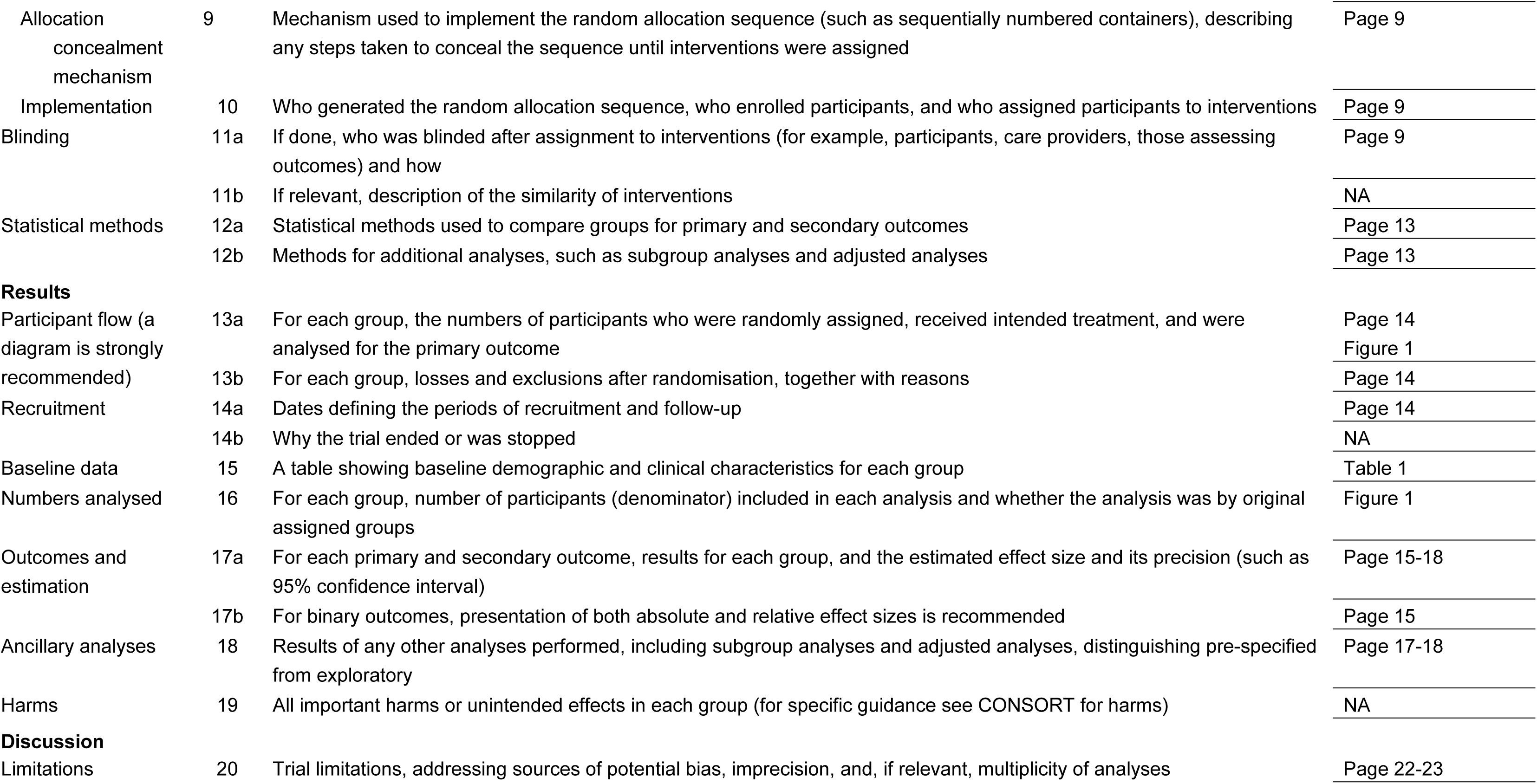
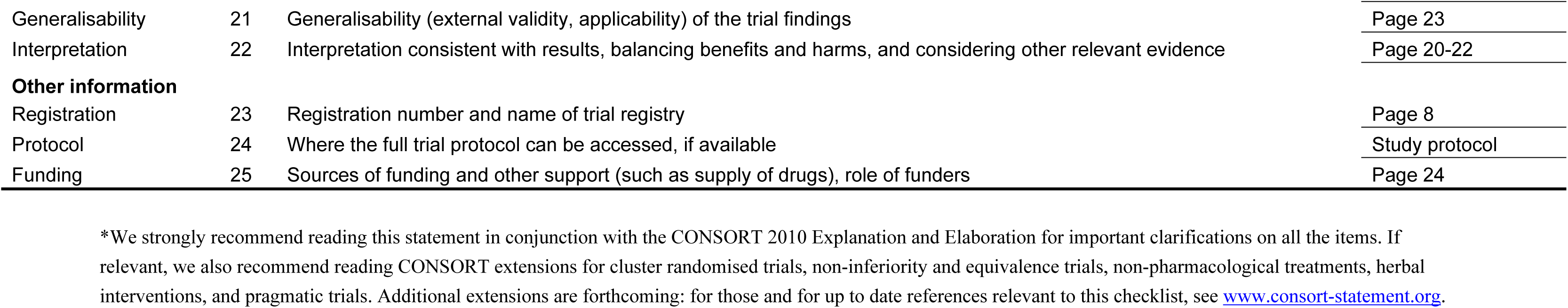

## Study Protocol

**Study on heterologous prime-boost immunization of recombinant COVID-19 vaccine (Ad5 vector) and RBD-based protein subunit vaccine (CHO)**

Protocol Number: JSVCT115

Principle Investigator: Jing-Xin Li

Sponsor: Jiangsu Provincial Center for Disease Control and Prevention

Version: Version 1.3

Protocol Date: July 11, 2021

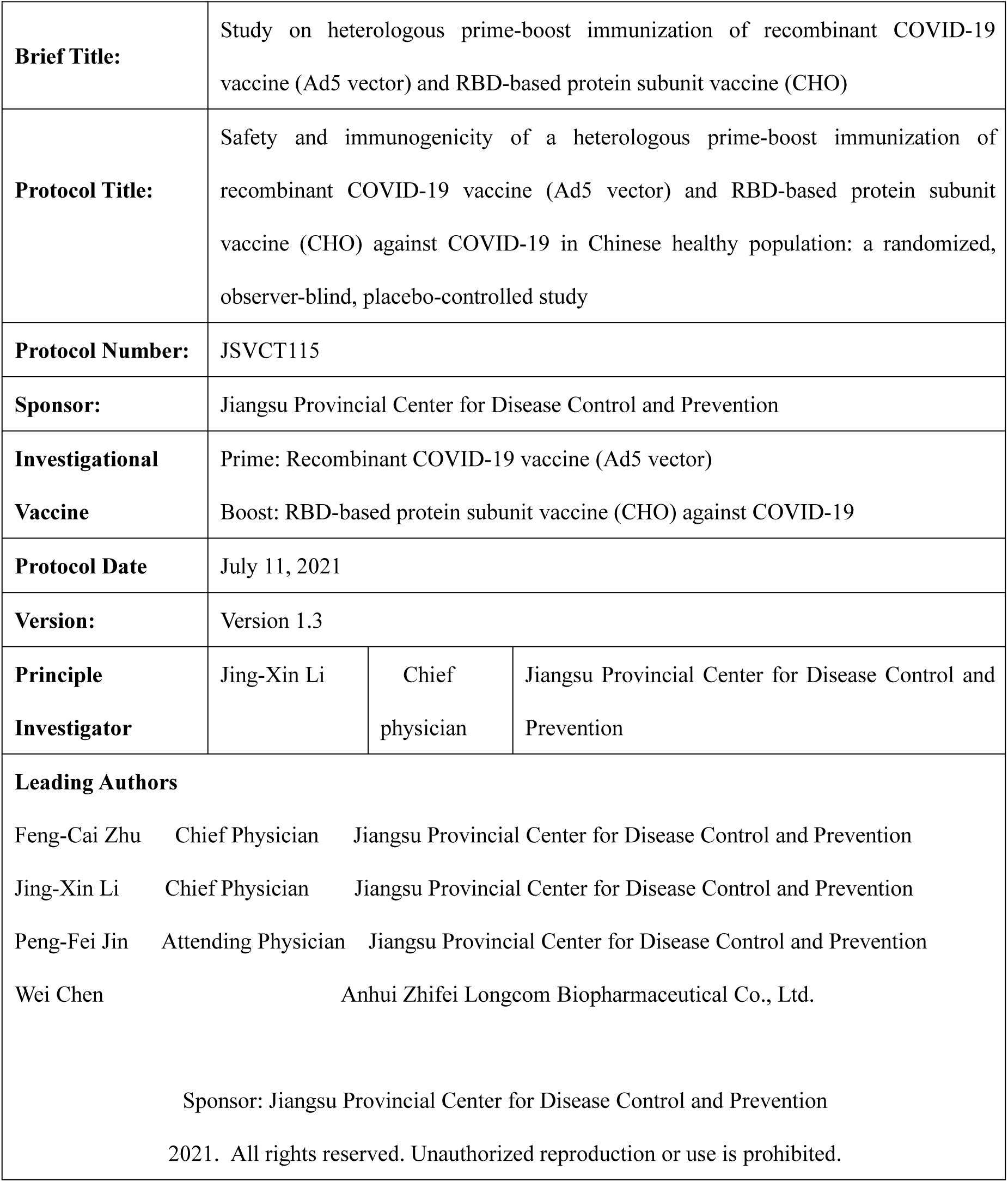

|  |  |  |  |
| --- | --- | --- | --- |
| **Brief Title:** | Study on heterologous prime-boost immunization of recombinant COVID-19 vaccine (Ad5 vector) and RBD-based protein subunit vaccine (CHO) |  |  |
| **Protocol Title:** | Safety and immunogenicity of a heterologous prime-boost immunization of recombinant COVID-19 vaccine (Ad5 vector) and RBD-based protein subunit vaccine (CHO) against COVID-19 in Chinese healthy population: a randomized, observer-blind, placebo-controlled study |  |  |
| **Protocol Number:** | JSVCT115 |  |  |
| **Sponsor:** | Jiangsu Provincial Center for Disease Control and Prevention |  |  |
| **Investigational Vaccine** | Prime: Recombinant COVID-19 vaccine (Ad5 vector)<br>Boost: RBD-based protein subunit vaccine (CHO) against COVID-19 |  |  |
| **Protocol Date** | July 11, 2021 |  |  |
| **Version:** | Version 1.3 |  |  |
| **Principle Investigator** | Jing-Xin Li | Chief physician | Jiangsu Provincial Center for Disease Control and Prevention |
| **Leading Authors**<br>Feng-Cai Zhu Chief Physician Jiangsu Provincial Center for Disease Control and Prevention<br>Jing-Xin Li Chief Physician Jiangsu Provincial Center for Disease Control and Prevention<br>Peng-Fei Jin Attending Physician Jiangsu Provincial Center for Disease Control and Prevention<br>Wei Chen Anhui Zhifei Longcom Biopharmaceutical Co., Ltd.<br>Sponsor: Jiangsu Provincial Center for Disease Control and Prevention<br>2021. All rights reserved. Unauthorized reproduction or use is prohibited. |  |  |  |

## Statement by Principal Investigator

I agree:

″ Assume the primary investigator responsibility for this clinical study.
″ Ensure that the study is carried out in accordance with the protocol and standard operating procedure (SOP) in site.
″ Ensure that no changes to the protocol are made without the review and written approval of the IEC, unless necessary to eliminate immediate harm to subjects or to comply with regulatory requirements (e.g., administrative aspects).
″ I am fully in control of the proper use of the investigational vaccines as described in the protocol.
″ I am familiar with and will comply with the Good Practice for Quality Management of Drug Clinical Trials (GCP) and all relevant regulatory requirements.

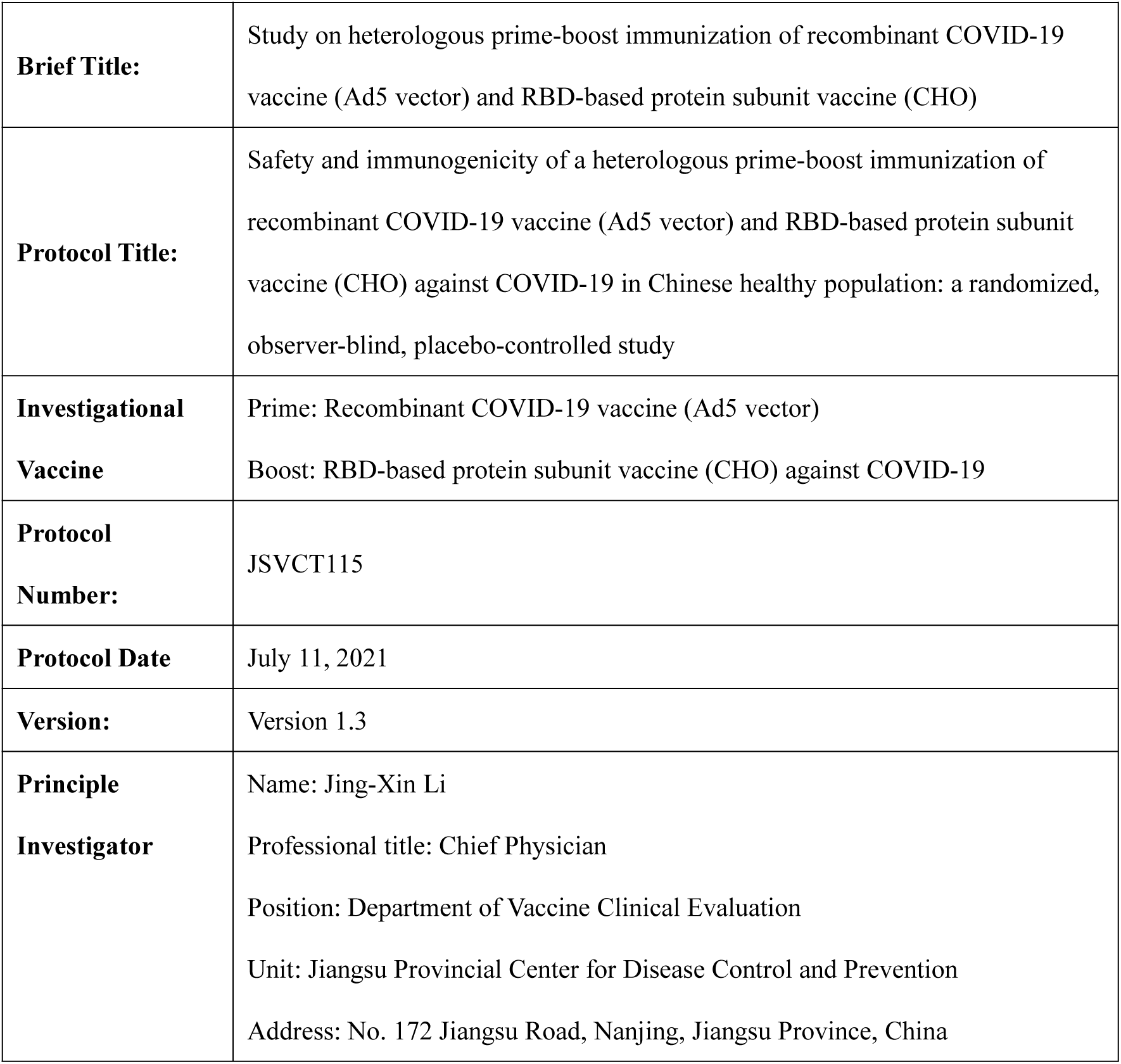
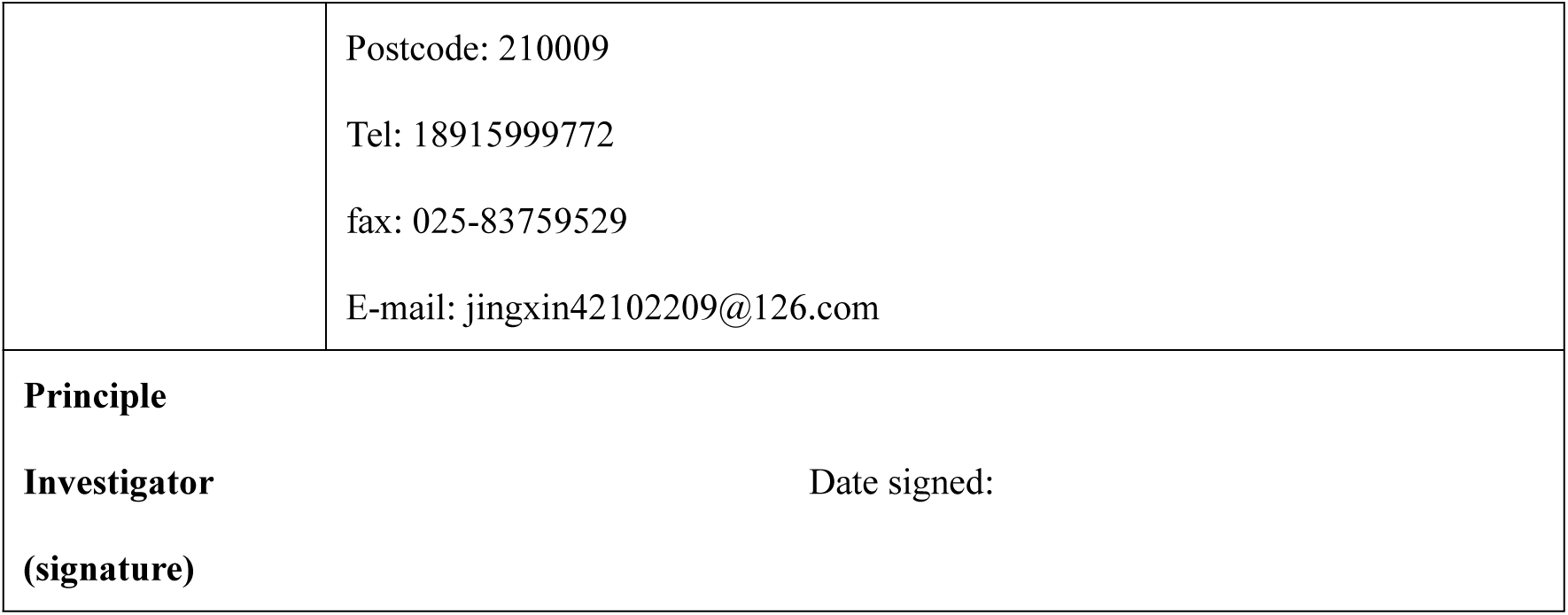

## DOCUMENT HISTORY

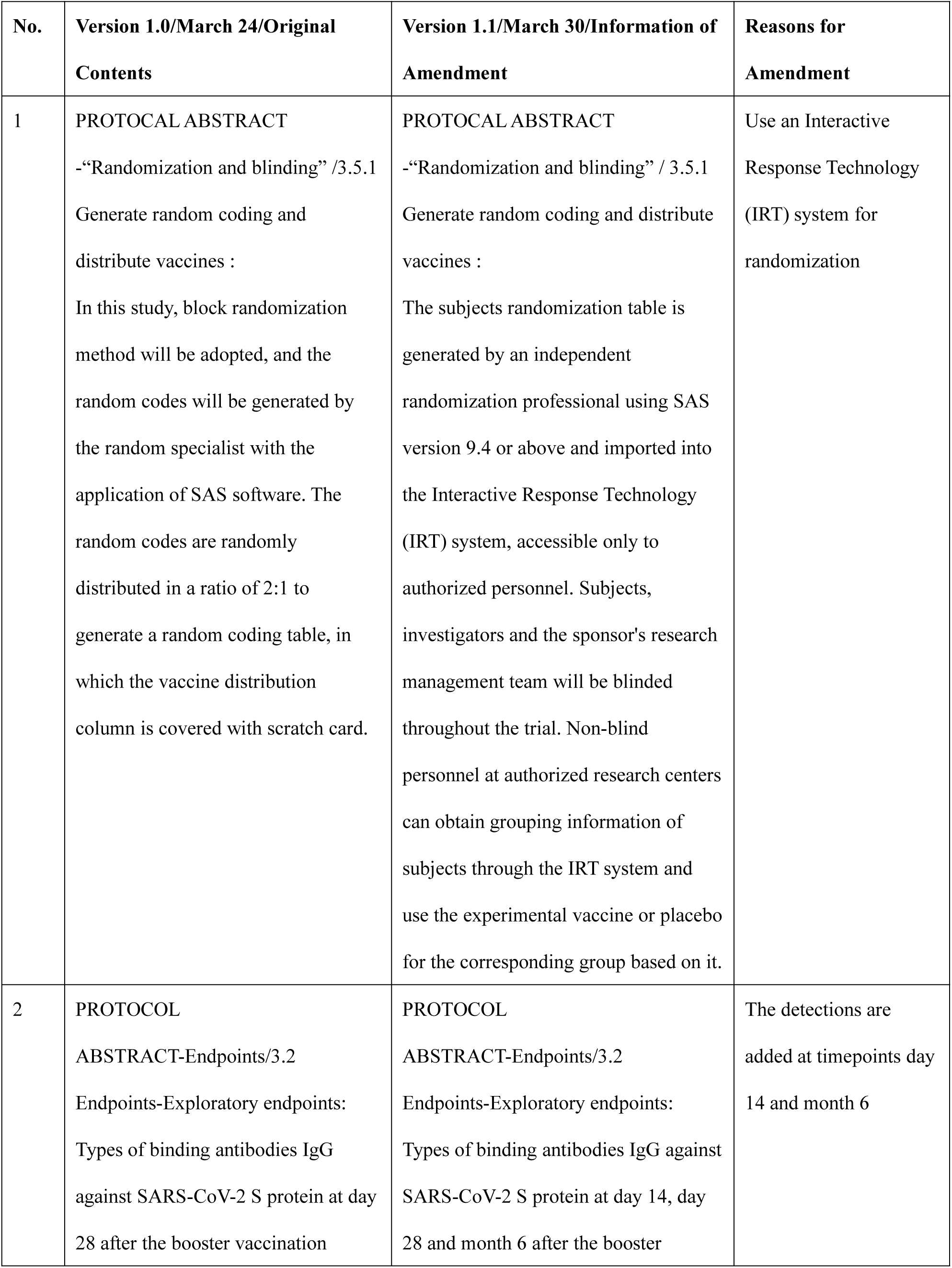
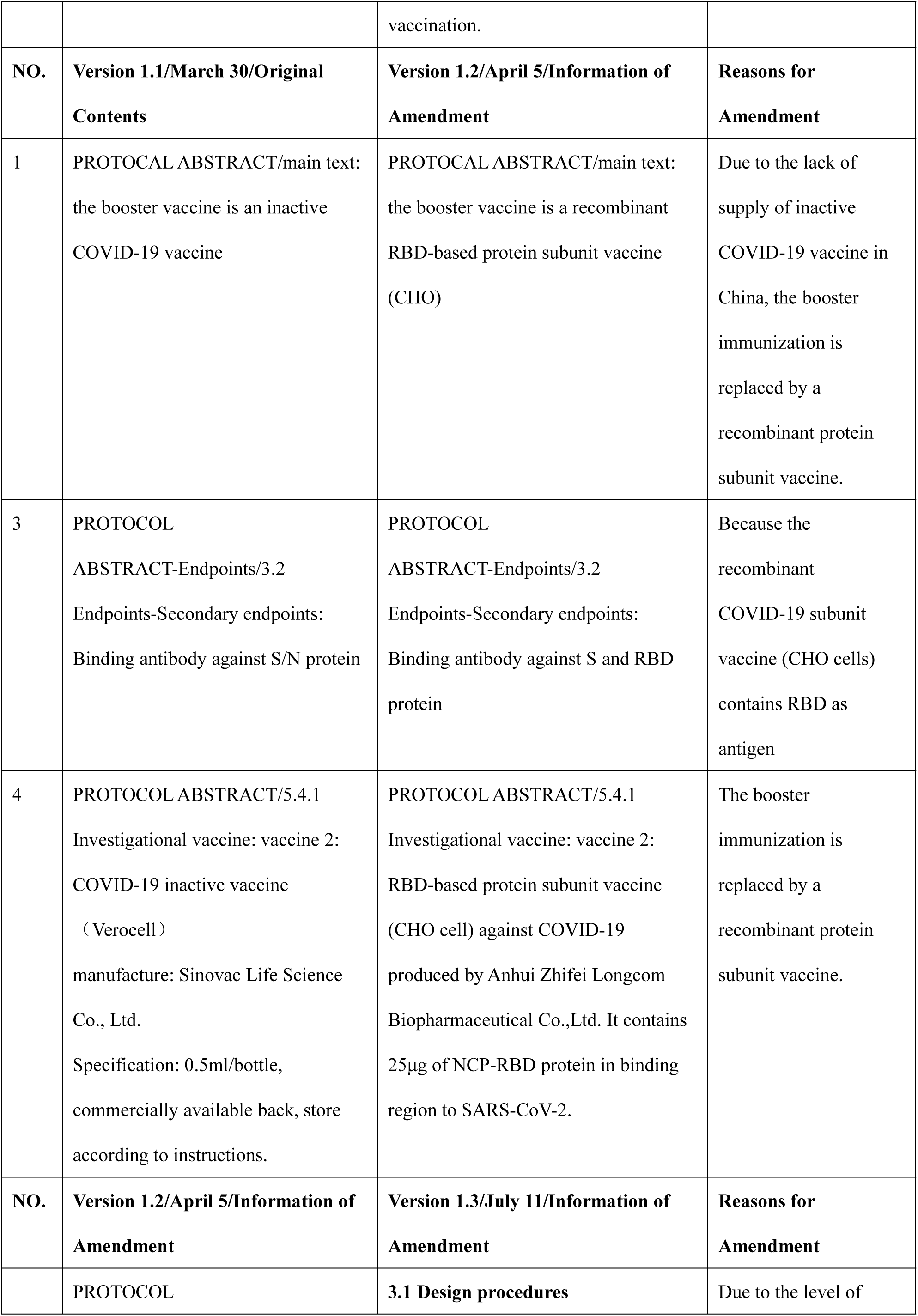
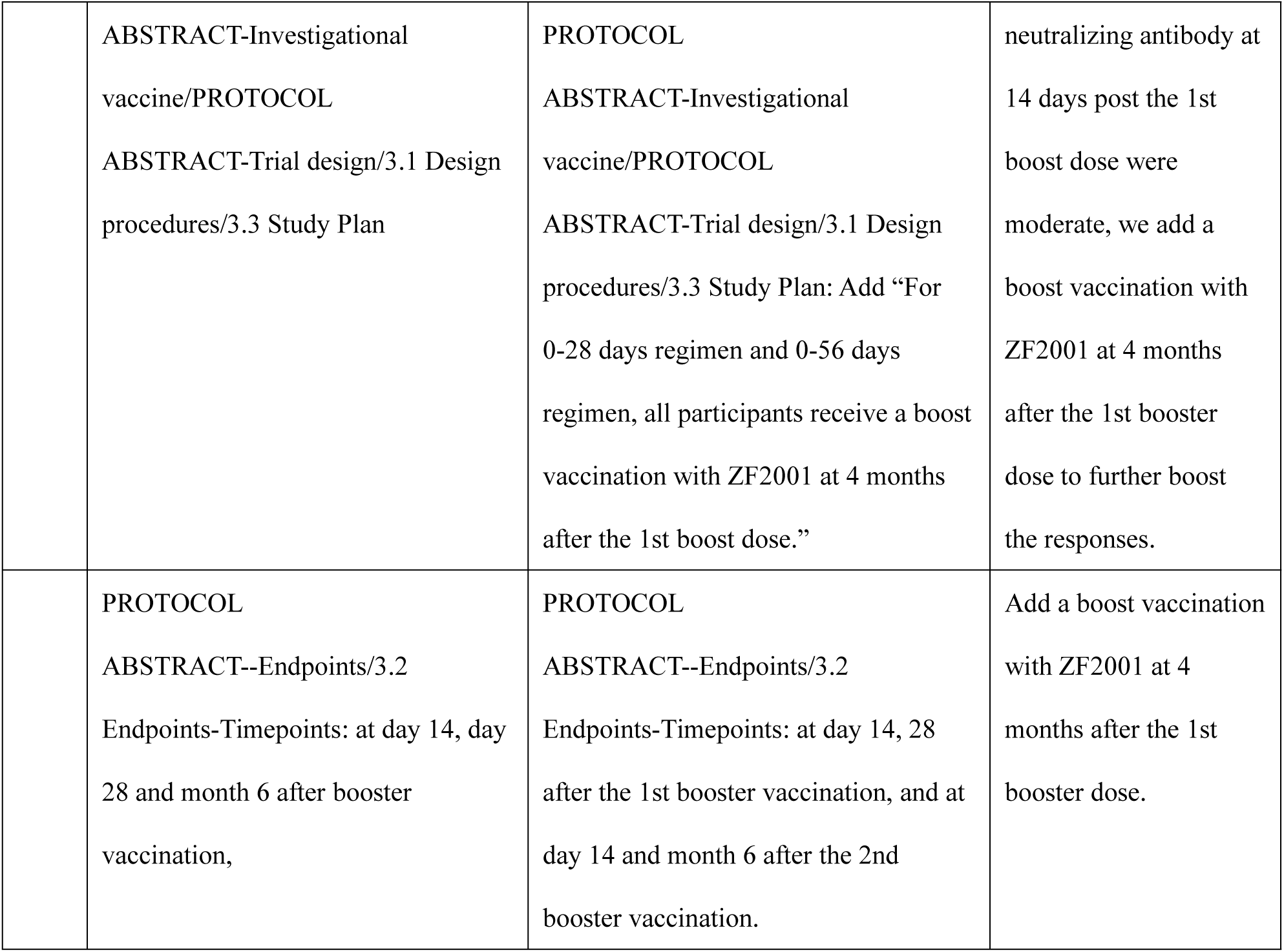

## ABBREVIATIONS

AE: Adverse Event
AR: Adverse Reaction
COVID-19: Corona Virus Disease 2019
eCRF: Electronic Case Report Form
ELISA: Enzyme-linked Immunosorbent Assay
FAS: Full Analysis Set
GCP: Good Clinical Practice
GMFI: Geometric Mean Fold Increase
GMP: Good Manufacturing Practice
GMT: Geometric Mean Titre
IEC: Independent Ethics Committee
ITT: Intent-to-treat
NIFDC: National Institute for Food and Drug Control
NMPA: National Medical Products Administration
PPS: Per Protocol Set
SAE: Serious Adverse Event
SOP: Standard Operation Procedure
SS: Safety Set

## PROTOCOL ABSTRACT

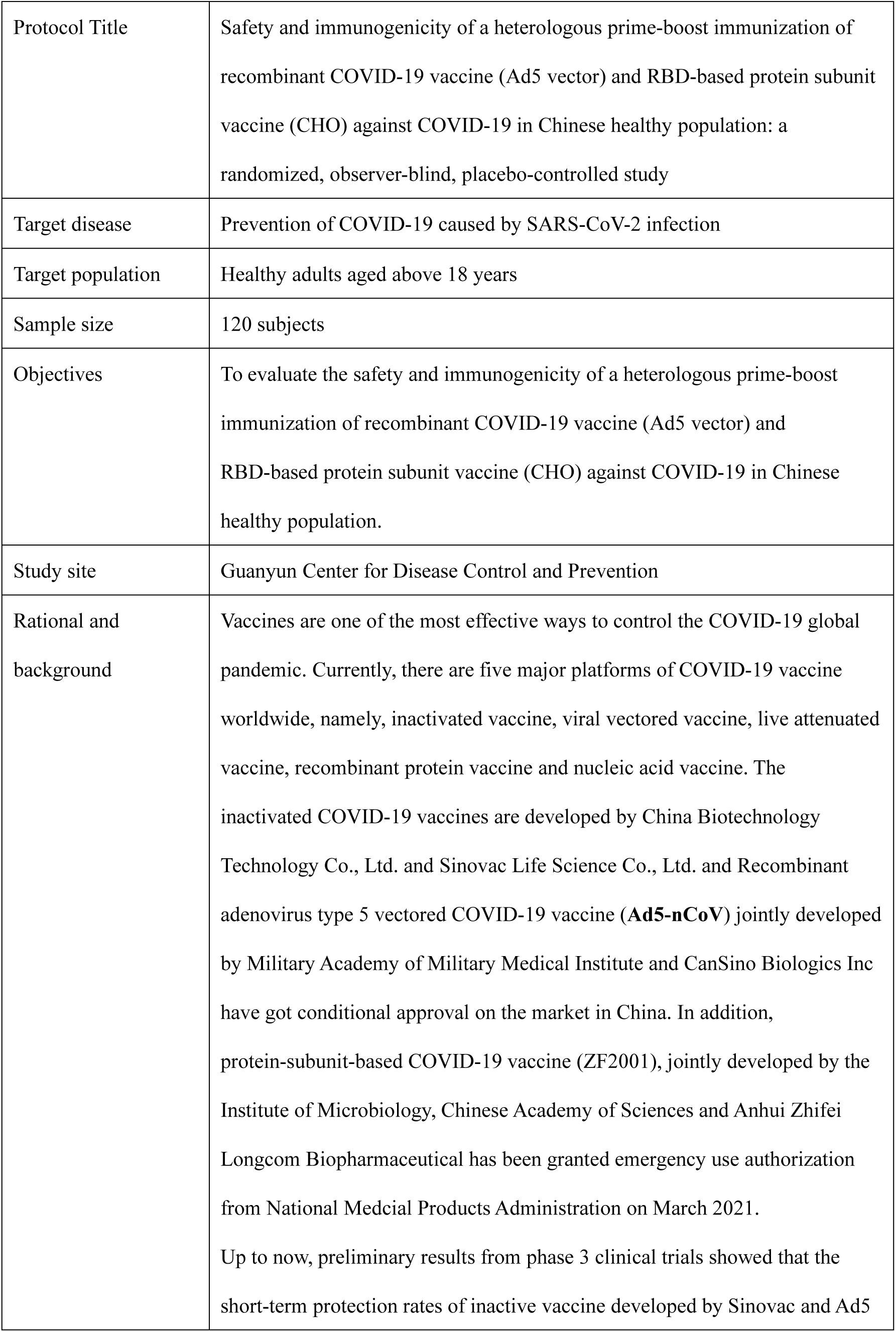
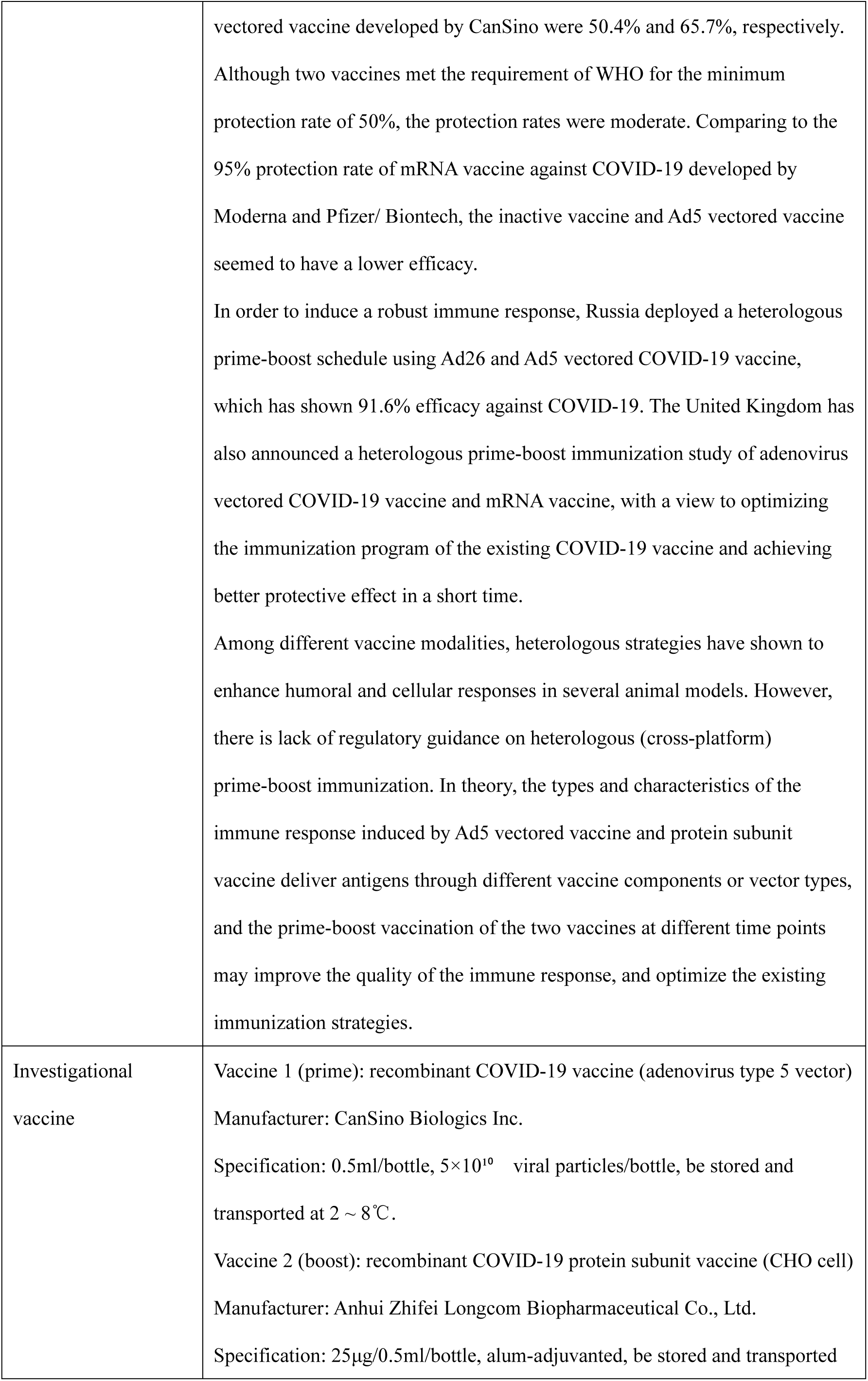
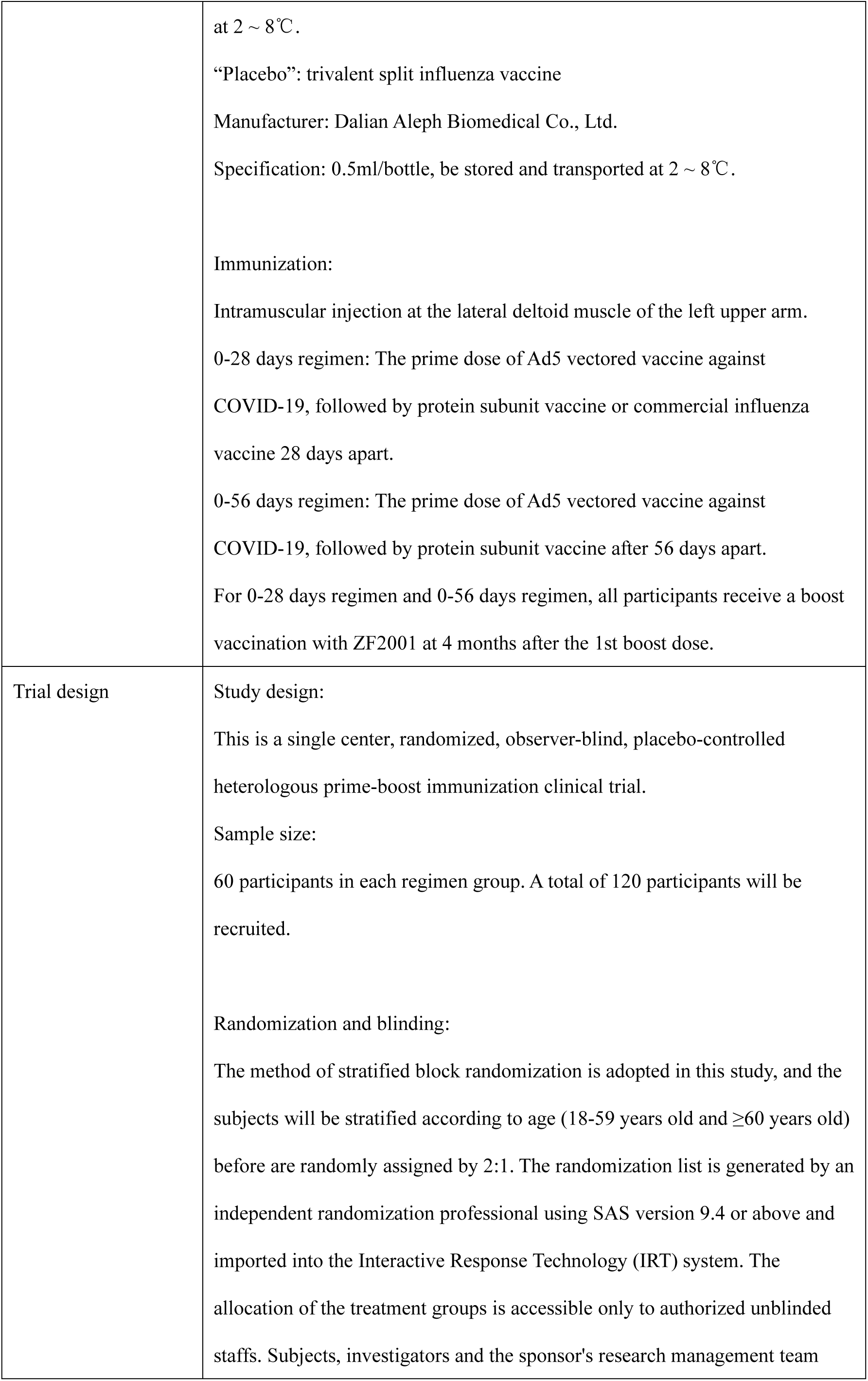
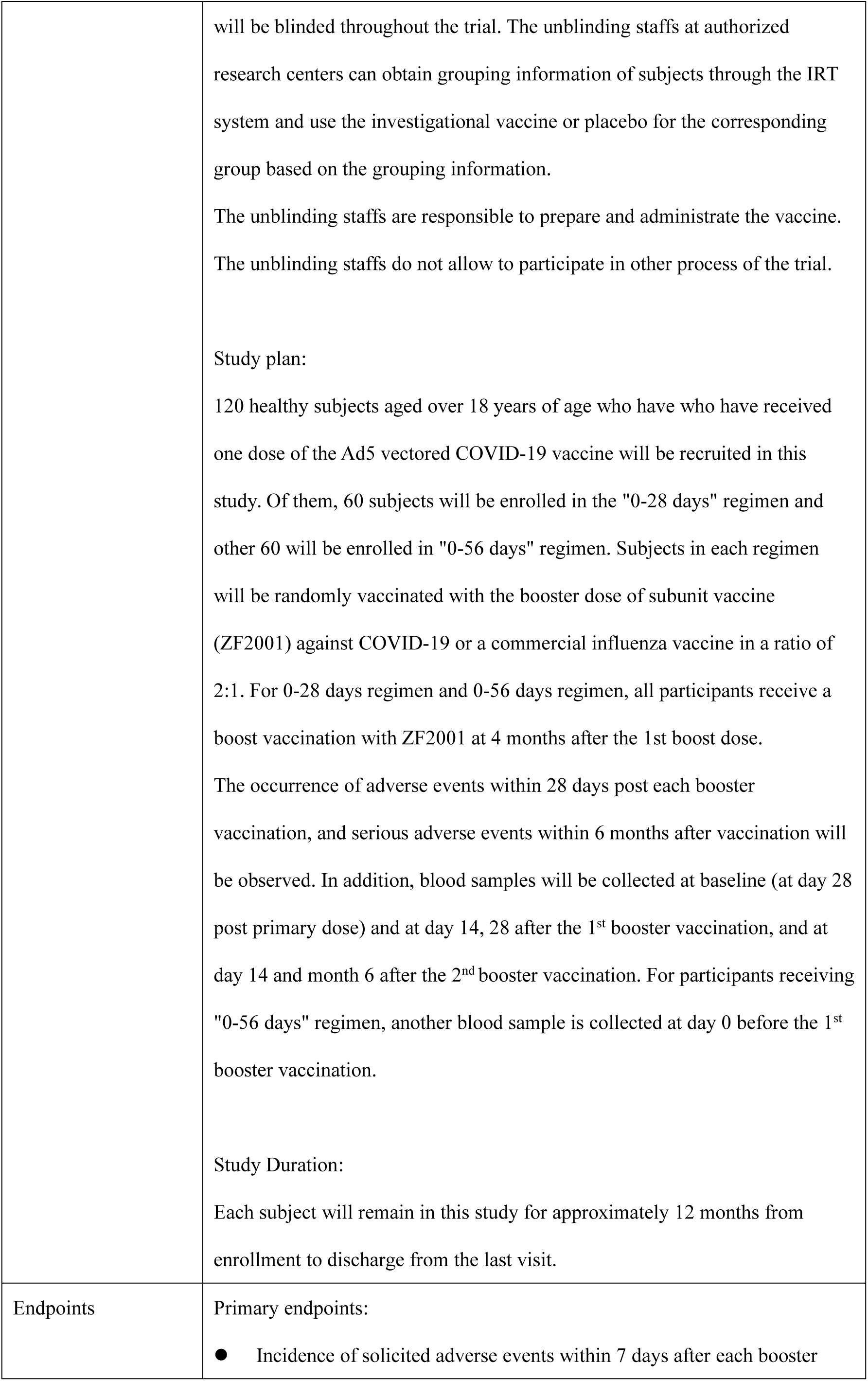
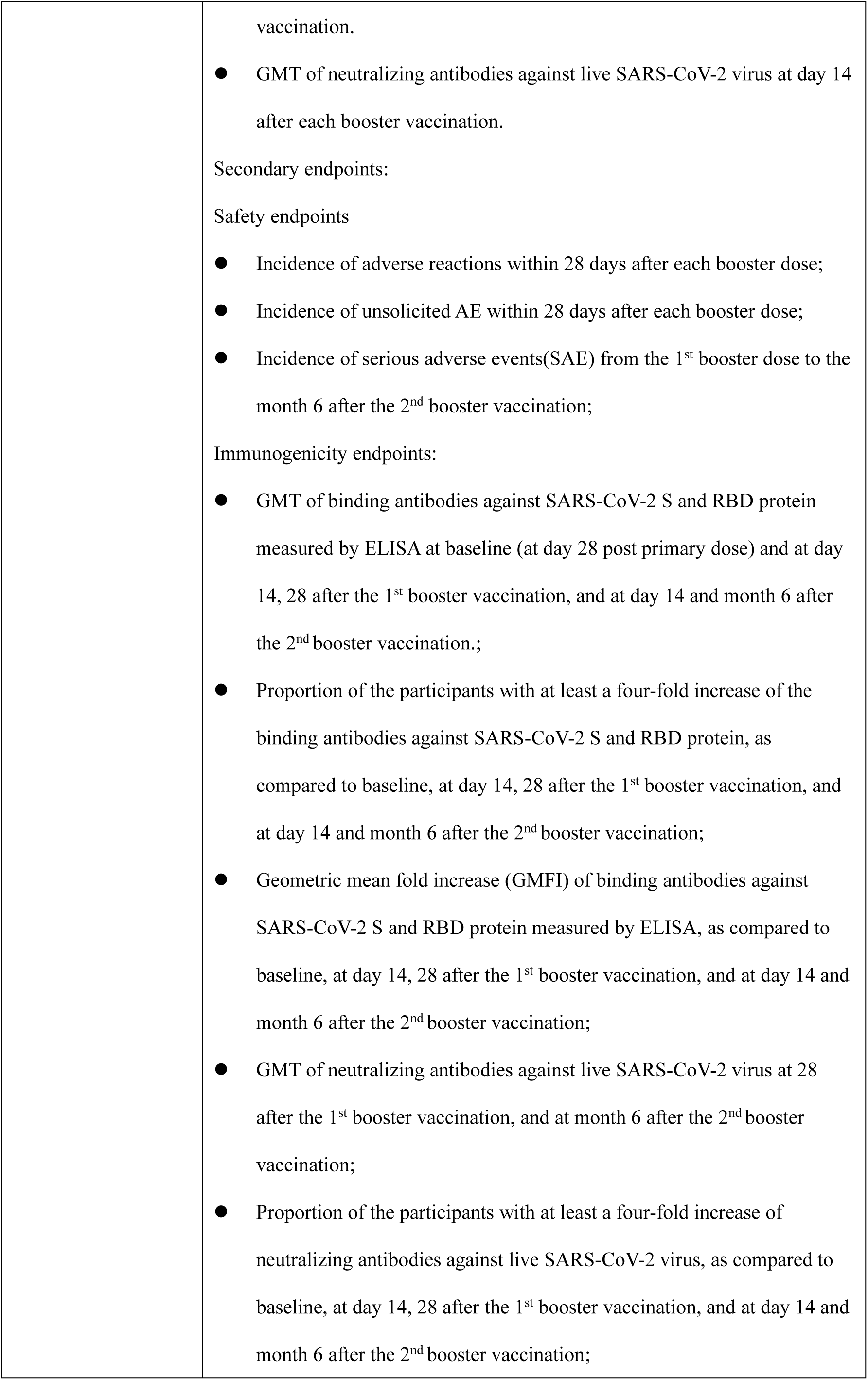
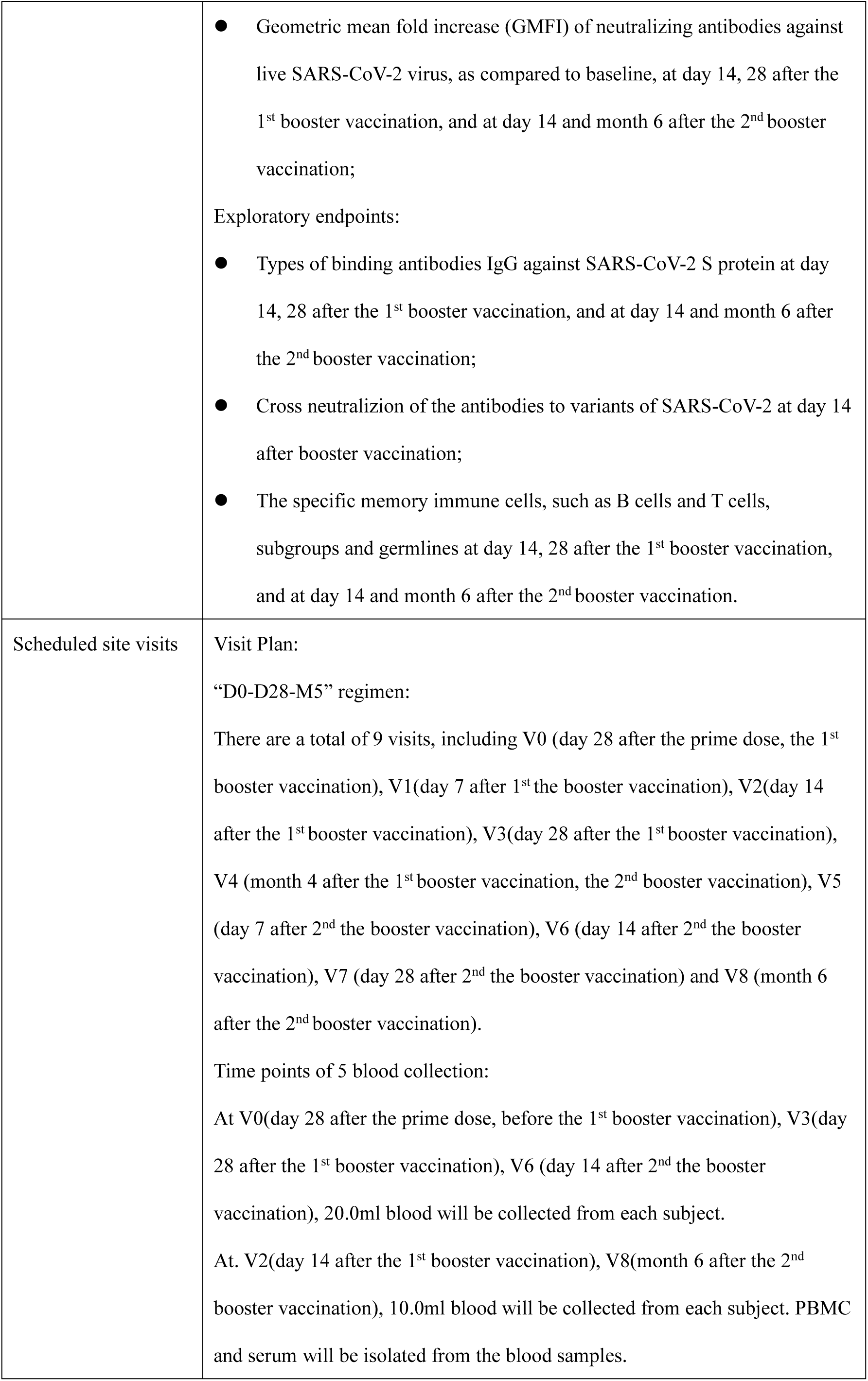
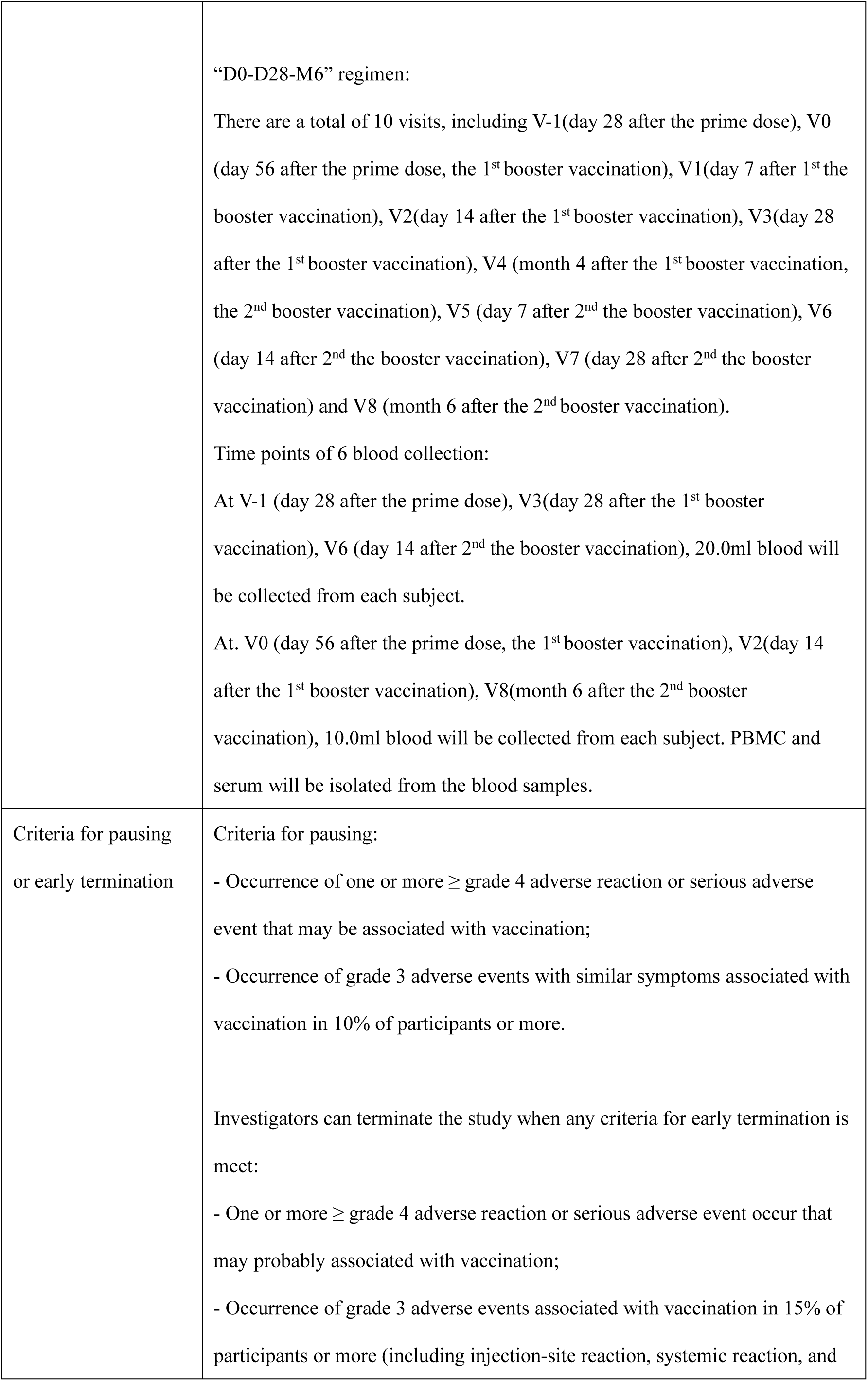
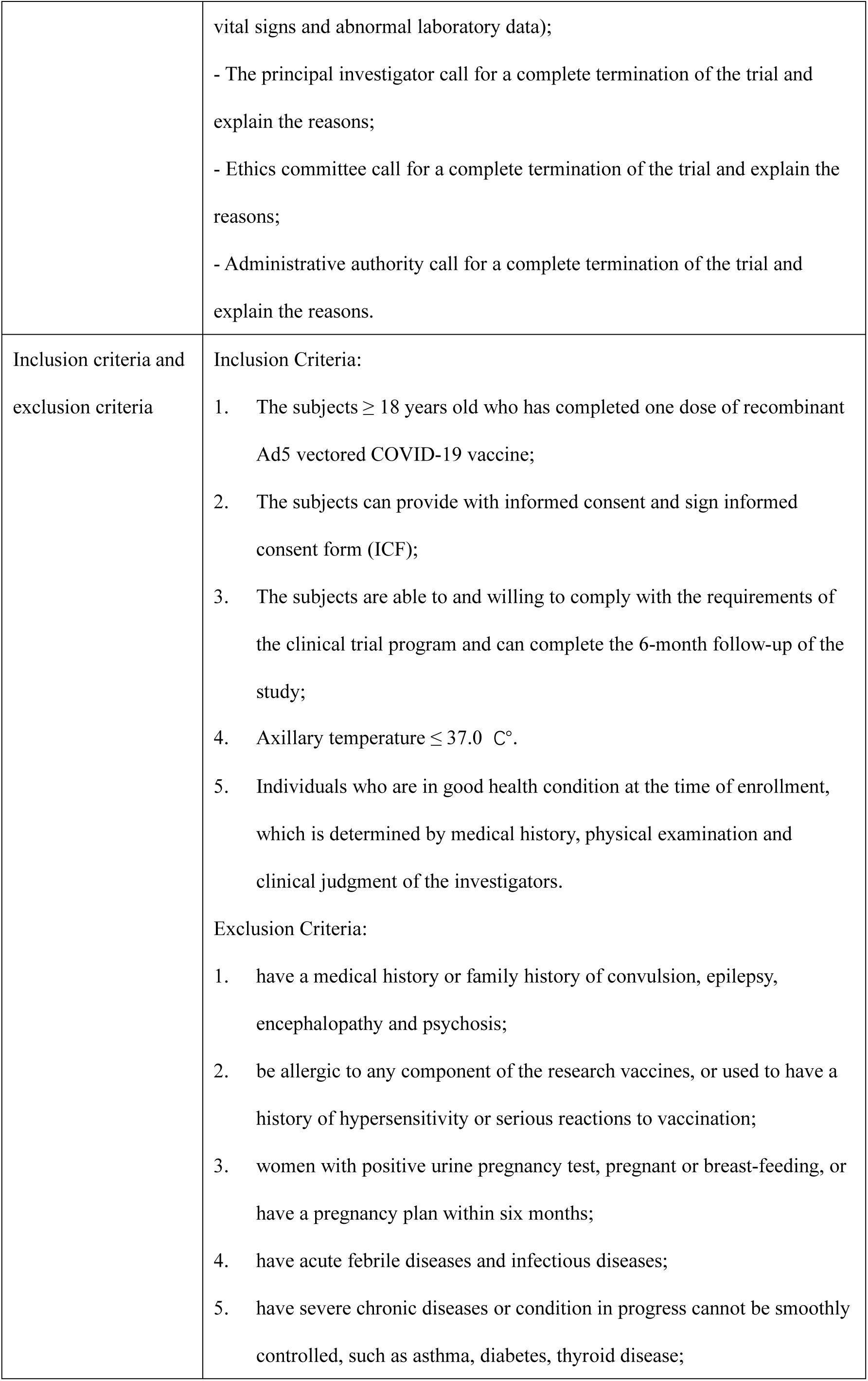
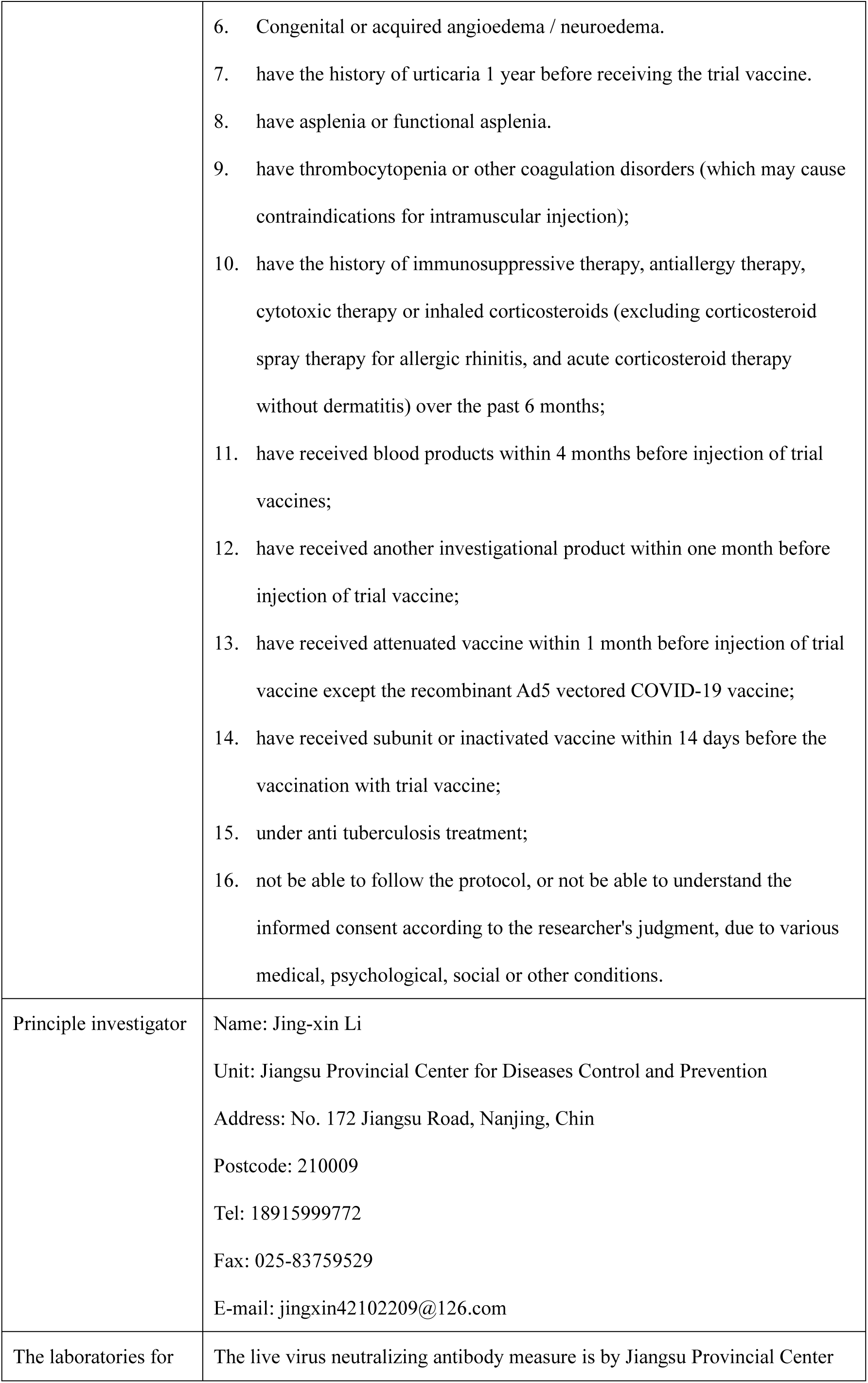
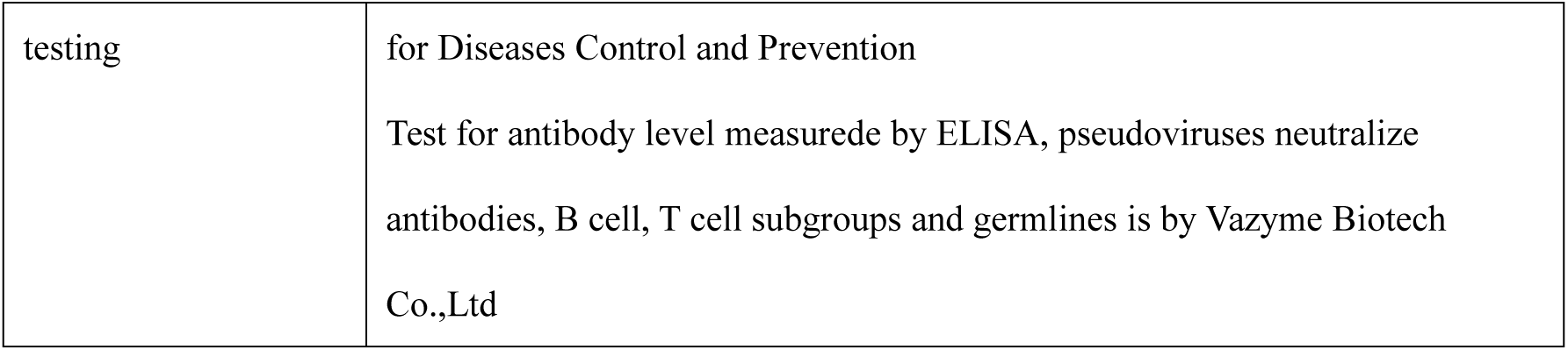

**Table 1.** “0-28 days” regimen subjects visit plan

| Visit No. | V0* | V1 | V2 | V3 | V4 | V5 | V6 | V7 | V8 |
| --- | --- | --- | --- | --- | --- | --- | --- | --- | --- |
| Day/Month | Day 28 | Day 35 | Day 42 | Day 56 | Day 150 | Day 157 | Day 164 | Day 178 | Day 330 |
| Visit interval | V0 | V0+7 days | V0+14 days | V0+28 days | V0+4 months | V4+7 days | V4+14 days | V0+28 days | V4+6 months |
| Time window | ±3 days | +3 days | +3 days | +3 days | +7 days | +3 days | +3 days | +3 days | ±15 days |
| Informed consent | ● |  |  |  | ● |  |  |  |  |
| Demographic information collection | ● |  |  |  |  |  |  |  |  |
| Physical examination and preliminary screening | ● |  |  |  | ● |  |  |  |  |
| Randomization | ● |  |  |  |  |  |  |  |  |
| Blood collection | ●(20ml) |  | ●(10ml) | ●(20ml) |  |  | ●(20ml) |  | ●(10ml) |
| Observation for 30 min post-vaccination | ● |  |  |  | ● |  |  |  |  |
| Safety visit(AR/AE) | ● | ● | ● | ● | ● | ● | ● | ● | ● |
| Report serious adverse event(SAE)※ | ● | ● | ● | ● | ● | ● | ● | ● | ● |
| Distribution of diary card | ● |  |  |  | ● |  |  |  |  |
| Return of diary card and distribute a contact card |  | ● |  |  |  | ● |  |  |  |
| Return of contact card |  |  |  | ● |  |  |  | ● |  |
| Record on the Vaccination | ● | ● | ● | ● | ● | ● | ● | ● | ● |
| and Visit Record Form |  |  |  |  |  |  |  |  |  |
| Record the combination drug/vaccine | • | • | • | • | • | • | • | • | • |

\* V0 is the time for the enrollment for a booster dose

**Table 2.** “0-56 day” regimen subjects visit plan

| Visit No. | V-1 | V0* | V1 | V2 | V3 | V4 | V5 | V6 | V7 | V8 |
| --- | --- | --- | --- | --- | --- | --- | --- | --- | --- | --- |
| Day/Month | Day 28 | Day 56 | Day 63 | Day 70 | Day 84 | Day 180 | Day 187 | Day 194 | Day 208 | Day 360 |
| Visit interval | V-1 | V0 | V0+<br>7 days | V0+<br>14 days | V0+<br>28 days | V0+<br>4 months | V4+7 days | V4+<br>14 days | V0+<br>28 days | V4+<br>6 months |
| Time window | 0 | ±3 days | +3 days | +3 days | +3 days | +7 days | +3 days | +3 days | +3 days | ±15 days |
| Informed consent | • |  |  |  |  | • |  |  |  |  |
| Demographic information collection | • |  |  |  |  |  |  |  |  |  |
| Physical examination and preliminary screening | • | • |  |  |  | • |  |  |  |  |
| Randomization |  | • |  |  |  |  |  |  |  |  |
| Blood collection | •(20ml) | •(10ml) |  | •(10ml) | •(20ml) |  |  | •(20ml) |  | •(10ml) |
| Observation for 30 min post-vaccination |  | • |  |  |  | • |  |  |  |  |
| Safety visit(AR/AE) |  | • | • | • | • | • | • | • | • | • |
| Report serious adverse event(SAE)※ |  | • | • | • | • | • | • | • | • | • |
| Distribution of diary card |  | • |  |  |  | • |  |  |  |  |
| Return of diary card and distribute a contact card |  |  | • |  |  |  | • |  |  |  |
| Return of contact card |  |  |  |  | • |  |  |  | • |  |
| Record on the Vaccination<br>and Visit Record Form | • | • | • | • | • | • | • | • | • | • |
| Record the combination<br>drug/vaccine |  | • | • | • | • | • | • | • | • | • |

\* V0 is the time for the enrollment for a booster dose.

## 1. Background and Principle

### 1.1 Pathogen

2019 Novel Coronavirus 2019(SARS-CoV-2) belongs to the genus β of coronavirus, with enveloped granules that are round or elliptic, often pleomorphic, with diameters ranging from 60 nm to 140nm. Its genetic characteristics are significantly different from those of SARS-CoV and MERS-CoV.

SARS-CoV-2 Coronaviruses belong to the genus Coronavirus in the family Coronaviridae. Coronaviruses are single-stranded RNA viruses with an envelope. They are a large group of viruses that exist widely in nature. Globally, 10% to 30% of upper respiratory tract infections are caused by HCoV-229E, HCoV-OC43, HCoV-NL63 and HCoV-HKU1, which are the second leading cause of the common cold, after rhinoviruses. Middle East Respiratory Syndrome (MERS) and Severe Acute Respiratory Syndrome (SARS), caused by coronavirus, are known to be serious infectious diseases.

The coronavirus genome encodes spike protein (S), envelope protein (E), membrane protein (M) and nucleoprotein (N) in sequence. Among them, S protein is the most important surface protein of coronavirus, which is related to the transmission ability of the virus. S protein contains two subunits: S1 and S2. S1 mainly contains receptor binding region, which is responsible for the recognition of cellular receptors. S2 contains the basic elements for the membrane fusion process. In the previous development of SARS and MERS vaccines, S protein was used as the most important candidate antigen.

### 1.2 Disease and epidemiological background

The COVID-19 is mainly characterized by fever, dry cough and fatigue. A small number of patients have symptoms such as nasal congestion, runny nose, sore throat, myalgia and diarrhea. Severe patients usually develop dyspnea and/or hypoxemia one week after onset, and in severe cases, rapid progression to acute respiratory distress syndrome, septic shock, refractory metabolic acidosis, haemorrhagic dysfunction and multiple organ failure, etc. It is worth noting that the course of the disease in the severe and critical patients may be moderate to low fever, or even no obvious fever. Some children and newborns showed atypical symptoms, such as diarrhea, vomiting and other digestive tract symptoms, or only mental weakness and shortness of breath.

At present, the source of infection is mainly patients infected by SARS-CoV-2. An asymptomatic infected person may also be a source of infection. The main route of transmission is by respiratory droplets and close contact is the main route of transmission. Exposure to high concentrations of aerosols in a relatively closed environment for a long period of time has the potential for aerosol transmission. SARS-CoV-2 can be isolated from feces and urine, and attention should be paid to the aerosol or contact transmission caused by feces and urine to environmental pollution. The population is generally susceptible.

### 1.3 Basis of the study

Currently, there are five major platforms of COVID-19 vaccine worldwide, namely, inactivated vaccine, viral vectored vaccine, live attenuated vaccine, recombinant protein vaccine and nucleic acid vaccine. The inactivated COVID-19 vaccines are developed by China Biotechnology Technology Co., Ltd. and Sinovac Life Science Co., Ltd. and Recombinant adenovirus type 5 vectored (**Ad5-nCoV**) jointly developed by Military Academy of Military Medical Institute and CanSino Biologics Inc have got conditional approval on the market in China. In addition, protein-subunit-based COVID-19 vaccine (ZF2001), jointly developed by the Institute of Microbiology, Chinese Academy of Sciences and Anhui Zhifei Longcom Biopharmaceutical has been granted emergency use authorization from National Medcial Products Administration on March 2021.

Up to now, preliminary results from phase 3 clinical trials showed that the short-term protection rates of inactive vaccine developed by Sinovac and Ad5 vectored vaccine developed by CanSino were 50.4% and 65.7%, respectively. Although two vaccines met the requirement of WHO for the minimum protection rate of 50%, the protection rates were moderate. Comparing to the 95% protection rate of mRNA vaccine against COVID-19 developed by Moderna and Pfizer/ Biontech, the inactive vaccine and Ad5 vectored vaccine seemed to have a lower efficacy.

In order to induce a robust immune response, Russia deployed a heterologous prime-boost schedule using Ad26 and Ad5 vectored COVID-19 vaccine, which has shown 91.6% efficacy against COVID-19. The United Kingdom has also announced a heterologous prime-boost immunization study of adenovirus vectored COVID-19 vaccine and mRNA vaccine, with a view to optimizing the immunization program of the existing COVID-19 vaccine and achieving better protective effect in a short time.

Among different vaccine modalities, heterologous strategies have shown to enhance humoral and cellular responses in several animal models. However, there is lack of regulatory guidance on heterologous (cross-platform) prime-boost immunization. In theory, the types and characteristics of the immune response induced by Ad5 vectored vaccine and protein subunit vaccine deliver antigens through different vaccine components or vector types, and the prime-boost vaccination of the two vaccines at different time points may improve the quality of the immune response, and optimize the existing immunization strategies.

In this study, subjects who have completed a dose of Ad5 vectored COVID-19 vaccine will be enrolled again to receive a booster dose of COVID-19 recombinant protein subunit vaccine at 28 or 56 days after the prime dose, which forms a heterologous prime-boost immunization program. To evaluate the effect of immunogenicity after immunization, the subjects in each regimen will be randomly vaccinated with the booster dose of subunit vaccine(ZF2001) against COVID-19 or a commercial influenza vaccine in a ratio of 2:1. In addition, due to clinical trial (JSVCT093) with two doses of Ad5 vectored COVID-19 vaccine and the phaseⅠ/ Ⅱ clinical trial with three dose of recombinant protein subunit vaccine (CHO cells) in healthy adults prior to this study, their serum samples after last vaccination can be provided for parallel detection of immunogenicity data, to provide complete control data.

The clinical study protocol is formulated in accordance with the requirements of the Vaccine Administration Law, the Good Practice for Quality Management of Drug Clinical Trials (GCP), the Technical Guiding Principles for Quality Management of Vaccine Clinical Trials and the Guiding Principles for Quality Management of Vaccine Clinical Trials (Trial).

## 2. Research Purposes

To evaluate safety and immunogenicity of a heterologous prime-boost immunization of recombinant COVID-19 Vaccine (Ad5 Vector) and RBD-based protein subunit vaccine (CHO) against COVID-19 in Chinese healthy adults.

## 3. Trial Design

### 3.1 Design procedures

This study is a single center, randomized, observer blind, placebo-controlled heterologous prime-boost immunization clinical trial, with “0-28 days” and “0-56 days” immunization regimens. The subjects are divided into two age groups, i.e. 18-59 years and 60 years and above.

Stage 1: According to the “0-28 days” regimen, sixty eligible subjects (30 in each age group) meeting the protocol requirements will be randomly assigned in a 2:1 ratio to receive a RBD-based protein subunit vaccine (CHO) or placebo (influenza vaccine).

Stage 2: According to the “0-56 days” regimen, sixty eligible subjects (30 in each age group) meeting the protocol requirements will be randomly assigned in a 2:1 ratio to receive a RBD-based protein subunit vaccine (CHO) or placebo (influenza vaccine).

Stage 3: For 0-28 days regimen and 0-56 days regimen, all participants receive a boost vaccination with ZF2001 at 4 months after the 1st boost dose.

### 3.2 Study Endpoints

#### 3.2.1 Primary endpoints

− Incidence of solicited adverse events within 7 days after each booster vaccination.
− GMT of neutralizing antibodies against live SARS-CoV-2 virus at day 14 after each booster vaccination.

#### 3.2.2 Secondary endpoints

**1. Safety endpoints**

− Incidence of adverse reactions within 28 days after each booster dose;
− Incidence of unsolicited AE within 28 days after each booster dose;
− Incidence of serious adverse events(SAE) from the 1^st^ booster dose to the month 6 after the 2^nd^ booster vaccination

**2. Immunogenicity endpoints**

− GMT of binding antibodies against SARS-CoV-2 S and RBD protein measured by ELISA at baseline (at day 28 post primary dose) and at day 14, 28 after the 1st booster vaccination, and at day 14 and month 6 after the 2nd booster vaccination.;
− Proportion of the participants with at least a four-fold increase of the binding antibodies against SARS-CoV-2 S and RBD protein, as compared to baseline, at day 14, 28 after the 1st booster vaccination, and at day 14 and month 6 after the 2nd booster vaccination;
− Geometric mean fold increase (GMFI) of binding antibodies against SARS-CoV-2 S and RBD protein measured by ELISA, as compared to baseline, at day 14, 28 after the 1st booster vaccination, and at day 14 and month 6 after the 2nd booster vaccination;
− GMT of neutralizing antibodies against live SARS-CoV-2 virus at 28 after the 1st booster vaccination, and at month 6 after the 2nd booster vaccination;
− Proportion of the participants with at least a four-fold increase of neutralizing antibodies against live SARS-CoV-2 virus, as compared to baseline, at day 14, 28 after the 1st booster vaccination, and at day 14 and month 6 after the 2nd booster vaccination;
− Geometric mean fold increase (GMFI) of neutralizing antibodies against live SARS-CoV-2 virus, as compared to baseline, at day 14, 28 after the 1st booster vaccination, and at day 14 and month 6 after the 2nd booster vaccination;

#### 3.2.3 Exploratory endpoints

− Types of binding antibodies IgG against SARS-CoV-2 S protein at day 14, 28 after the 1st booster vaccination, and at day 14 and month 6 after the 2nd booster vaccination;
− Cross neutralizion of the antibodies to variants of SARS-CoV-2 at day 14 after booster vaccination;
− The specific memory immune cells, such as B cells and T cells, subgroups and germlines at day 14, 28 after the 1st booster vaccination, and at day 14 and month 6 after the 2nd booster vaccination.

### 3.3 Study Plan

120 healthy subjects aged over 18 years of age who have who have received one dose of the Ad5 vectored COVID-19 vaccine in three batches consistence clinical trials will be recruited in this study. Of them, 60 subjects will be enrolled in the “0-28 days” regimen and other 60 will be enrolled in “0-56 days” regimen. Subjects in each regimen will be randomly assigned to receive the booster dose of subunit vaccine (ZF2001) against COVID-19 or a commercial influenza vaccine in a ratio of 2:1. For 0-28 days regimen and 0-56 days regimen, all participants receive a boost vaccination with ZF2001 at 4 months after the 1st boost dose.

The occurrence of adverse events within 28 days and serious adverse events within 6 months after vaccination will be observed. In addition, blood samples will be collected at baseline (at day 28 post primary dose), and at day 14, day 28 post 1^st^ booster dose, and at day 14 and month 6 after the 2^nd^ booster vaccination to test serum antibody levels and to profile the specific memory immune cells and antibody repertoire. Each subject will remain in this study for approximately 12 months.

In “0-28 days” regimen, participants will attend 9 visits in total, including V0 (day 28 after the prime dose, the 1^st^ booster vaccination), V1(day 7 after 1^st^ the booster vaccination), V2(day 14 after the 1^st^ booster vaccination), V3(day 28 after the 1^st^ booster vaccination), V4 (month 4 after the 1^st^ booster vaccination, the 2^nd^ booster vaccination), V5 (day 7 after 2^nd^ the booster vaccination), V6 (day 14 after 2^nd^ the booster vaccination), V7 (day 28 after 2^nd^ the booster vaccination) and V8 (month 6 after the 2^nd^ booster vaccination).

In “0-56 days” regimen, participants will attend 10 visits in total, including V-1(day 28 after the prime dose), V0 (day 56 after the prime dose, the 1^st^ booster vaccination), V1(day 7 after 1^st^ the booster vaccination), V2(day 14 after the 1^st^ booster vaccination), V3(day 28 after the 1^st^ booster vaccination), V4 (month 4 after the 1^st^ booster vaccination, the 2^nd^ booster vaccination), V5 (day 7 after 2^nd^ the booster vaccination), V6 (day 14 after 2^nd^ the booster vaccination), V7 (day 28 after 2^nd^ the booster vaccination) and V8 (month 6 after the 2^nd^ booster vaccination).

**Table 1.** “0-28 days” regimen subjects visit plan

| Visit No. | V0* | V1 | V2 | V3 | V4 | V5 | V6 | V7 | V8 |
| --- | --- | --- | --- | --- | --- | --- | --- | --- | --- |
| Day/Month | Day 28 | Day 35 | Day 42 | Day 56 | Day 150 | Day 157 | Day 164 | Day 178 | Day 330 |
| Visit interval | V0 | V0+<br>7 days | V0+<br>14 days | V0+<br>28 days | V0+<br>4 months | V4+<br>7 days | V4+<br>14 days | V0+<br>28 days | V4+<br>6 months |
| Time window | ±3 days | +3 days | +3 days | +3 days | +7 days | +3 days | +3 days | +3 days | ±15 days |
| Informed consent | ● |  |  |  | ● |  |  |  |  |
| Demographic information collection | ● |  |  |  |  |  |  |  |  |
| Physical examination and preliminary screening | ● |  |  |  | ● |  |  |  |  |
| Randomization | ● |  |  |  |  |  |  |  |  |
| Blood collection | ●(20ml) |  | ●(10ml) | ●(20ml) |  |  | ●(20ml) |  | ●(10ml) |
| Observation for 30 min post-vaccination | ● |  |  |  | ● |  |  |  |  |
| Safety visit(AR/AE) | ● | ● | ● | ● | ● | ● | ● | ● | ● |
| Report serious adverse event(SAE)※ | ● | ● | ● | ● | ● | ● | ● | ● | ● |
| Distribution of diary card | ● |  |  |  | ● |  |  |  |  |
| Return of diary card and distribute a contact card |  | ● |  |  |  | ● |  |  |  |
| Return of contact card |  |  |  | ● |  |  |  | ● |  |
| Record on the Vaccination and Visit Record Form | • | • | • | • | • | • | • | • | • |
| Record the combination drug/vaccine | • | • | • | • | • | • | • | • | • |

\* V0 is the time for the enrollment for a booster dose.

**Table 2.** “0-56 days” regimen subjects visit plan

| Visit No. | V-1 | V0* | V1 | V2 | V3 | V4 | V5 | V6 | V7 | V8 |
| --- | --- | --- | --- | --- | --- | --- | --- | --- | --- | --- |
| Day/Month | Day 28 | Day 56 | Day 63 | Day 70 | Day 84 | Day 180 | Day 187 | Day 194 | Day 208 | Day 360 |
| Visit interval | V-1 | V0 | V0+<br>7 days | V0+<br>14 days | V0+<br>28 days | V0+<br>4 months | V4+7 days | V4+<br>14 days | V0+<br>28 days | V4+<br>6 months |
| Time window | 0 | ±3 days | +3 days | +3 days | +3 days | +7 days | +3 days | +3 days | +3 days | ±15 days |
| Informed consent | • |  |  |  |  | • |  |  |  |  |
| Demographic information collection | • |  |  |  |  |  |  |  |  |  |
| Physical examination and preliminary screening | • | • |  |  |  | • |  |  |  |  |
| Randomization |  | • |  |  |  |  |  |  |  |  |
| Blood collection | •(20ml) | •(10ml) |  | •(10ml) | •(20ml) |  |  | •(20ml) |  | •(10ml) |
| Observation for 30 min post-vaccination |  | • |  |  |  | • |  |  |  |  |
| Safety visit(AR/AE) |  | • | • | • | • | • | • | • | • | • |
| Report serious adverse event(SAE)※ |  | • | • | • | • | • | • | • | • | • |
| Distribution of diary card |  | • |  |  |  | • |  |  |  |  |
| Return of diary card and<br>distribute a contact card |  |  | • |  |  |  | • |  |  |  |
| Return of contact card |  |  |  |  | • |  |  |  | • |  |
| Record on the Vaccination<br>and Visit Record Form | • | • | • | • | • | • | • | • | • | • |
| Record the combination<br>drug/vaccine |  | • | • | • | • | • | • | • | • | • |

\* V0 is the time for the enrollment for a booster dose.

### 3.4 Sample size calculation

Sample size:

Hypothesis: GMT of vaccine group is superior to that in the control group at day 28 after the booster vaccination.

The baseline GMT level before the booster immunization is expected to be about 1:20 (log_10_X=1.3) after one dose of prime vaccination with Ad5 vectored COVID-19 vaccine. After the booster dose, GMT level in the vaccine group is estimated to reach 1:60 (log_10_X =1.78), while the control group remains 1:20. Assume that the Standard Deviation is about 4 (log_10_X=0.6), and the allocation ratio for the vaccine group and placebo group is 2:1, then the minimal sample size to provide 80% power calculated is 40 and 20, respectively.

In each regimen group, subjects are stratified to two sub-age groups: 18-59 years and 60 years or above.

**Table 3.** Sample size of each immunization regimen

| regimen | Sub-age group | sample size |  |
| --- | --- | --- | --- |
|  |  | Experimental vaccine group | Placebo control group |
| 0-28 days | 18-59 years | 20 | 10 |
|  | 60 years or above | 20 | 10 |
| 0-56 days | 18-59 ages | 20 | 10 |
|  | 60 years or above | 20 | 10 |
| Total | — | 80 | 40 |

### 3.5 Randomization and blinding

#### 3.5.1 Generate random coding and distribute vaccines

The study adopts the method of stratified block randomization, and subjects will be randomly assigned by 2:1 according to age group (18-59 years old and ≥60 years old). The subjects randomization table is generated by an independent randomization professional using SAS version 9.4 or above and imported into the Interactive Response Technology (IRT) system, accessible only to authorized personnel. Subjects, investigators and the sponsor’s research management team will be blinded throughout the trial. Non-blind personnel at authorized research centers can obtain grouping information of subjects through the IRT system and use the experimental vaccine or placebo for the corresponding group based on it.

#### 3.5.2 Maintenance of blinding

Subjects, safety observers, laboratory testers will be blinded.

Those who administer, prepare and administer vaccines are unblinded staff and must sign a blinding maintenance agreements to ensure that any documents of the unblinding information are only accessible for the authorized non-blinded staff. The labels on vaccine syringe will be covered with a study number label after the preparation of the vaccine and put it ready to use.

#### 3.5.3 Unblinding

The investigators must not disrupt the blind study of the vaccine/placebo unless the study information is medically necessary for the subjects. In the event of a medical emergency, the principal investigator should be contacted as far as possible before disrupting the study vaccine/placebo blinding to discuss the need for an urgent unblinding.

Blinding will be uncover when completing the initial analysis of safety and immunogenicity 28 days after the booster dose, but the subjects and safety observers will remain blinded.

### 3.6 Criteria for pausing or early termination

Criteria for pausing:

- Occurrence of one or more ≥ grade 4 adverse reaction or serious adverse event that may be associated with vaccination;
- Occurrence of grade 3 adverse events associated with vaccination in 10% of participants or more.
- Administrative authority call for a complete termination of the trial and explain the reasons. investigators can terminate the study when any criteria for early termination is meet:
- One or more ≥ grade 4 adverse reaction or serious adverse event occur that may probably associated with vaccination;
- Occurrence of grade 3 adverse events associated with vaccination in 15% of participants or more (including injection-site reaction, systemic reaction, and vital signs and abnormal laboratory data;
- The principal investigator call for a complete termination of the trial and explain the reasons;
- Ethics committee call for a complete termination of the trial and explain the reasons;

### 3.7 Duration of study

It will take about 12 months for each participant from recruiting to completing the last visit.

## 4. Participants

### 4.1 Participants selection

Healthy people aged 18 and above who have been vaccinated of a dose recombinant COVID-19 Vaccine (Ad5 Vector), are selected as the target population, and informed in writing by informed consent approved by the ethics committee. On the premise that the volunteers themselves will sign the informed consent, they can only participate in the study after passing the physical examination and the following inclusion and exclusion criteria.

### 4.2 Inclusion criteria

− The subjects over 18 years old who has completed one dose of recombinant Ad5 vectored COVID-19 vaccine;
− The subjects can provide with informed consent and sign informed consent form (ICF);
− The subjects are able to and willing to comply with the requirements of the clinical trial program and can complete the 6-month follow-up of the study;
− Axillary temperature ≤ 37.0 ℃.
− Individuals who are in good health condition at the time of entry into the trial as determined by medical history, physical examination and clinical judgment of the investigators and meet the requirements of these products immunization.

### 4.3 Exclusion Criteria

− Have the medical history or family history of convulsion, epilepsy, encephalopathy and psychosis;
− Be allergic to any component of the research vaccines, or used to have a history of hypersensitivity or serious reactions to vaccination;
− Women with positive urine pregnancy test, pregnant or breast-feeding, or have a pregnancy plan within six months;
− Have acute febrile diseases and infectious diseases;
− Have severe chronic diseases or condition in progress cannot be smoothly controlled, such as asthma, diabetes, thyroid disease;
− Congenital or acquired angioedema / neuroedema.
− Have the history of urticaria 1 year before receiving the trial vaccine.
− Have asplenia or functional asplenia.
− Have thrombocytopenia or other coagulation disorders (which may cause contraindications for intramuscular injection);
− Have the history of immunosuppressive therapy, antiallergy therapy, cytotoxic therapy or inhaled corticosteroids (excluding corticosteroid spray therapy for allergic rhinitis, and acute corticosteroid therapy without dermatitis) over the past 6 months;
− Have received blood products within 4 months before injection of trial vaccines;
− Have received another investigational product within 1 month before injection of trial vaccine;
− Have received attenuated vaccine within 1 month before injection of trial vaccine except the recombinant Ad5 vectored COVID-19 vaccine;
− Have received subunit or inactivated vaccine within 14 days before the vaccination with trial vaccine;
− Under anti tuberculosis treatment;
− Not be able to follow the protocol, or not be able to understand the informed consent according to the researcher’s judgment, due to various medical, psychological, social or other conditions.

### 4.4 Withdraw from the study

Participants have the right to withdraw from the study at any time during the study period, and the investigators should record the reason of withdraw:

− Loss of contact, early withdraw of the study;
− Request to withdraw without any reason;
− Withdraw for reasons unrelated to the study, such as long-term departure, relocation, etc., and the specific reason for withdrawal should be recorded;
− Withdrawal for reasons related to the study, such as intolerance of adverse reactions, intolerance of biological specimen collection, etc., and the specific reason for withdrawal should be recorded. If a participant withdraw because of AE or SAE, investigators should follow up the participant until the resolve of AE or SAE.
− Participants can require a complete withdraw from the study, all study behaviors can be stopped, including vaccination, biological specimen collection and safety observation. The data before withdrawal will not be used for analysis if he or she require so. If the participants allow the investigators use the data collected before the withdrawal, the data can be included in analysis;
− Participants can require a partially withdraw from the study, such as refuse to vaccination or blood drawn only, but still participate in other procedures during the follow-up.

### 4.5 Complete of the study

#### 4.5.1 Complete of the safety data collection

The participants who receive experimental vaccine, and complete safety observation within 28 days post each booster dose, and reported SAEs through the study will be considered as complete of the safety data collection.

#### 4.5.2 Complete of immunogenicity data collection

The participants who meet the inclusion and do not meet any exclusion criteria, take the vaccination, and complete the visits and blood collection required by the study protocol will be considered as complete of the immunogenicity data collection.

### 4.6 Protocol violation and protocol deviation

#### 4.6.1 Protocol violation (including but not limited to)

− No informed consent singed by the participant;
− The enrolled participant does not meet the all the inclusion criteria or meet one or more exclusion criteria;
− The participant received incorrect intervention;
− The participant received a vaccine fail to meet the requirements;
− Any other reasons identified by the investigators and confirmed by the principal investigator.

#### 4.6.2 Protocol deviation (including but not limited to)

− Beyond the visiting time window;
− Low compliance of participants, and the participants do not complete the blood sample collection;
− Serious adverse events do not report in time (SAE);
− Participants are treated with unallowed drugs (intramuscular, oral or intravenous corticosteroids for ≥ 2mg/kg/days, continuous use for ≥ 14 days, or other immunosuppressants);
− The interval between vaccination with other vaccines is insufficient;
− Other reasons considered as protocol deviation by the principal investigator. Investigators or monitors should report any protocol violation or deviation to principal investigator or coordinators as soon as possible after knowing it by fax or e-mail. Protocol violation should also be reported to the ethics committees.

**5.0 Methods and procedures**

## 5.1 Participants selection

Healthy people aged 18 and above who have been vaccinated of a dose recombinant COVID-19 Vaccine (Ad5 Vector), are selected as the target population.

## 5.2 Informed Consent

When obtaining and recording informed consent, researchers should abide by relevant regulations, GCP and the ethical principles stipulated in the Declaration of Helsinki. Before the start of the study, the investigators should obtain written approval/consent from the ethics review committee for the informed consent form and other documents provided to the subjects.

Before participating in this clinical study, researchers should explain the contents of the informed consent form to the subjects and/or their witnesses, and the subjects and/or their witnesses should be given sufficient time to consult the details of the study before signing the informed consent form. When explaining the information of informed consent to multiple persons, each subject and/or witness should be given the opportunity to ask the investigators individually before signing the informed consent form. Researchers should keep the informed consent form signed by each subject, and provide the subject with a copy of the signed name and date of the informed consent form.

## 5.3 Physical examination and screening

The subjects’ body temperature will be measured before enrollment, and HCG detection will be performed on pre-menopausal women.

According to the “inclusion and exclusion criteria”, the interviewers conduct medical history inquiry and screening. Only those who passed the screening can be enrolled and participate in the randomization.

## 5.4 Vaccine distribution and inoculation

The unblind staff, who are responsible for vaccine preparation will assign the allocated treatment to the subjects according to the random number generated by an independent statistical party. After the preparation of the vaccine, they hand the ready-to-use syringes to the vaccination nurse, who will administrate the vaccination.

First aid drugs such as epinephrine hydrochloride and first aid equipment such as simple ventilator and ECG monitor should be provided at the vaccination site.

### 5.4.1 Investigational vaccine

Investigational vaccine 1 (prime dose): the recombinant New Coronavirus vaccine (adenovirus vector) produced by CanSino Biologics Inc, liquid dosage form, 0.5 ml/ bottle, contains recombinant replication defective human 5 adenovirus 5×10^10^ virus particles expressing New Coronavirus S protein. Investigational vaccine 2 (booster dose): the New Coronavirus recombinant subunit vaccine (CHO cell) produced by Anhui ZhiFei Longcom Biopharmaceutical Co., Ltd., “Zhi Ke Wei De”, 0.5ml/ bottle, contains 25 μg NCP-RBD protein of New Coronavirus spike protein binding region NCP-RBD . “Placebo”: a Trivalent split influenza vaccine produced by Dalian Aleph Biomedical Co., Ltd., liquid dosage form, 0.5 ml/ bottle, contains 15 μg of H1N1, 15 μg of H3N2 and 15 μg of B-line hemagglutinin.

### 5.4.2 Administration

Subjects are vaccinated according to the immunization procedure. The vaccine should be fully shaken before use, and should be used immediately after opening. In case of cracks, unclear or invalid labels, or abnormal appearance of the vaccine, it should not be used.

The vaccines inject intramuscularly at the attachment of the lateral deltoid muscle of the upper arm, with priority given to the left arm.

### 5.4.3 Storage and transportation of investigational vaccines

1. Vaccine storage: investigational vaccines must be stored in a safe and locked place, and must not be contacted by unauthorized persons. The temperature of vaccine storage place should be controlled in the range of 2-8℃ to prevent freezing; the storage temperature of vaccine should be recorded once in the morning and afternoon of each working day.
2. Vaccine transportation: vaccines are transported from the research site to the vaccination site, from the vaccination site to the research site, and stored in the refrigerator or freezer. Each cold chain equipment is equipped with a thermometer. The vaccine administrator records the temperature every 30 minutes, and fills in the transportation and storage temperature records in detail. The storage temperature (2-8℃) must be kept during transportation. Any over temperature must be reported to the site responsible researcher or project coordinator for instructions. All vaccine transportation processes must be recorded.

### 5.4.4 Combined medication/vaccine

When the medical events happen during the study period, the participant are allowed to carry out the appropriate medical treatment, but the medical treatment should be recorded in time.

Other vaccination is not recommended except for emergency during the research period, such as rabies vaccine, tetanus vaccine, or other emergent vaccination need. Any vaccine used is required to be recorded during the study period.

## 5.5 Safety follow up and evaluation

### 5.5.1 Safety observation

After vaccination, the participants will stay at the clinic for 30-minute safety observation. The trained researchers should systematically observe each subject, record the local and systemic reactions within 30 minutes, and record the severity.

The participants are followed for the next a few days, and asked to record the safety observation by themselves on the diary card till 7 days after the vaccination. From the day 8 to the day 28 after vaccination, the adverse events are recorded passively. At the clinic visit, the researcher will retrospectively check and verified the adverse events recorded during the safety observation. From day 28 to month 6 after vaccination, the subjects are asked to report only serious adverse events during this period.

### 5.5.2 Safety observation contents and indicators

. Adverse events from the clinical trial are graded according to the guiding principles for the classification of adverse events in clinical trials of preventive vaccines (NMPA [2019] No. 102), as follows: (table 4-5)

**Table 4.** Grading of (local) AEs at injection site

| Symptoms | Grade 1 | Grade 2 | Grade 3 | Grade 4 |
| --- | --- | --- | --- | --- |
| **Pain** | Do not affect or slightly affect physical activity | affect physical activity | Affect daily life | Loss of basic self-care ability or hospitalization |
| **Induration\*, swelling (optional)\*\* #** | Diameter 2.5~<5 cm or area 6.25~<25 cm<sup>2</sup> and does not affect or slightly affect daily life | Diameter 5~<10 cm or area 25~<100 cm<sup>2</sup> or affect daily life | Diameter ≥ 10 cm or area ≥ 100 cm<sup>2</sup> or ulceration or secondary infection or phlebitis or aseptic abscess or wound drainage or seriously affect daily life | Abscess, exfoliative dermatitis, dermal or deep tissue necrosis |
| **Rash\*, Redness (optional)\*\* #** | Diameter 2.5~<5 cm or area 6.25~25 cm<sup>2</sup> and does not affect or slightly affect daily life | Diameter 5~<10 cm or area 25~<100 cm<sup>2</sup> or affect daily life | Diameter ≥ 10 cm or area ≥ 100 cm<sup>2</sup> or ulceration or secondary infection or phlebitis or aseptic abscess or wound drainage or seriously affect daily life | Abscess, exfoliative dermatitis, dermal or deep tissue necrosis |
| **Itch** | Itching at the vaccination site, relieved by itself or within 48 hours after treatment | Itching at the vaccination site, which does not resolve within 48 hours after treatment | Affect daily life | NA |
| **Cellulitis** | NA | Non-injectable treatment is required (e.g. oral antibacterial, antifungal, antiviral therapy) | Intravenous treatment is required (e.g. intravenous antibacterial, antifungal, antiviral therapy) | Sepsis, or tissue necrosis, etc. |

Note: \*: in addition to directly measuring the diameter for grading and evaluation, the progress of the measurement results should also be recorded.
\*\* the maximum measuring diameter or area should be used.

**Table 5.** Grading for systemic adverse events.

| Sign | Grade 1 | Grade 2 | Grade 3 | Grade 4 |
| --- | --- | --- | --- | --- |
| **Fever [Axillary temperature (°C) ]**<br>**>14 years old** | 37.3~<38.0 | 38.0~<38.5 | 38.5~<39.5 | ≥ 39.5, last more than 3 days |
| **Gastrointestinal system** |  |  |  |  |
| Diarrhea | Mild or transient, 3 to 4 times a day, | Moderate or persistent, 5-7 times a day, | >7 times/day, abnormal stool, or | Hypotension shock, hospitalization |
|  | abnormal stool, or mild diarrhea last less than 1 week | abnormal stool characteristics, or diarrhea >1 week | hemorrhagic diarrhea, orthostatic hypotension, electrolyte imbalance, need intravenous infusion >2L | required |
| Dysphagia | Mild discomfort when swallowing | Diet is restricted | Diet and conversation are very limited; you can't eat solid food. | Can't eat liquid food; need parenteral nutrition. |
| Anorexia | Loss of appetite, but no reduction in food intake | Loss of appetite, reduced food intake, but no significant weight loss. | Loss of appetite and weight loss | Need for intervention (e.g. gastric tube feeding, parenteral nutrition) |
| Vomiting | 1- 2 times/24 hours and does not affect the activity | 3- 5 times/24 hours or activity is restricted | >6 times/24 hours or need intravenous rehydration | Hypotension shock requires hospitalization or other means of nutrition |
| Nausea | Transient (<24 hours) or intermittent and food intake is normal | Continued nausea leads to reduced food intake (24-48 hours) | Persistent nausea results in almost no food intake (> 48 hours) or requires intravenous fluid replacement | Life-threatening (eg hypotension shock) |
| **Musculoskeletal and connective tissue** |  |  |  |  |
| Non-injection-site muscle pain | Does not affect daily activities | Slightly affect daily activities | Severe muscle pain that seriously affects daily activities | Emergency or hospitalization |
| Arthritis | Mild pain with | Moderate pain with | Severe pain with | Permanent and/or |
|  | inflammation, erythema, or swelling of joints; but does not interfere with function | inflammation, erythema, or swelling of joints; impairs function but does not affect daily activities | inflammation, erythema, or joint swelling; affecting daily activities | disabling joint injury |
| Arthralgia | Mild pain without hindering function | Moderate pain; need analgesics and/or pain that impedes function but does not affect daily activities | Severe pain; need analgesics and/or pain affecting daily activities | Disability pain |
| **nervous system** |  |  |  |  |
| Headache | Does not affect daily activities and requires no treatment | Transient, slightly affects daily activities and may require treatment or intervention | Seriously affects daily activities and requires treatment or intervention | Intractable and requires emergency or hospitalization |
| Syncope | Close to syncope without losing consciousness (pre-syncope) | Loss of consciousness without treatment | Loss of consciousness and needs treatment or hospitalization | NA |
| **The spirit system** |  |  |  |  |
| Insomnia | Mild difficulty in falling asleep, not affecting or slightly affecting daily life | Moderate difficulty in falling asleep, affecting daily life | Serious difficulty in falling asleep, seriously affecting daily life, requiring treatment or hospitalization | NA |
| **Skin and subcutaneous tissue** |  |  |  |  |
| Non-injection-site | Slightly itchy without | Itching affects daily | Itching makes it | NA |
| itching (no skin lesions) | affecting or slightly affecting daily life | life | impossible to carry on daily life. |  |
| Abnormal skin and mucosa | Erythema/itching/col or change | Diffuse rash/macular papule/dryness/desquamation | Blister/exudation/desquamation/ulcer | Exfoliative dermatitis involving mucous membrane, or erythema multiforme, or suspected Stevens-Johnson syndrome |
| **The respiratory system** |  |  |  |  |
| Cough | Transient, without treatment | Persistent cough, effective treatment | Paroxysmal cough, uncontrollable treatment | Emergency or hospitalization |
| **The immune system** |  |  |  |  |
| Acute allergic reaction ** | Local urticaria (blister) without treatment | Local urticaria requiring treatment or mild angioedema without treatment | Extensive urticaria or angioedema requiring treatment or mild bronchospasm | Anaphylactic shock or life-threatening bronchospasm or throat edema |
| **Others** |  |  |  |  |
| Fatigue | Does not affect daily activities | Affects normal daily activities | Seriously affects daily activities and cannot work | Emergency or hospitalization |
| Non-injection-site pain# (Specify the location when reporting) | Minor pain that does not affect or slightly affect daily life | Pain affects daily life | Pain can't carry on daily life | Disability pain, loss of basic self-care ability |

Note: \* refers to type I hypersensitivity.

#### General principles for the grading for other adverse events

The intensity of adverse events not mentioned in the rating table shall be evaluated according to the following criteria:

| Grade 1 | Grade 2 | Grade 3 | Grade 4 | Grade 5 |
| --- | --- | --- | --- | --- |
| Mild: Short-term (< 48 hours) or mild discomfort, no influence on activities, treatment not indicated | Moderate: Mild or moderate restricted activities, presentation indicated possibly, treatment not indicated or mild treatment indicated | Severe: Significant restricted activities, presentation and treatment indicated, hospitalization indicated possibly | Critical: Life-threatening possibly, severely restricted activities, intensive care indicated | Death |

### 5.5.3 Outcome of AEs

The outcomes of ARs/AEs include: (1) Recovery; (2) Not yet recovered; (3) Recovered but sequelae; (4) Death; (5) Loss of visit.

### 5.5.4 Relationship between AE and vaccination

Investigators should make the best interpretation of AE, and assess the possible causal relationship between vaccination and reactions (such as history of underlying diseases, combined treatment of causation). This applies to all AEs, including severe ones and non-severe ones. The assessment of causality will be reasonably explained in the following or more aspects of the event: The similar reaction to the solution was observed in the past; identical events of similar types solution have been reported in the literature; the incident occurred along with the time of the vaccination, and again after the secondary vaccination According to definitions, all the solicited AE (that is, the local adverse event of the collection of the report) will be considered to be related to vaccination. The causal relationship of AE should be evaluated according to the following questions, and according to your judgment, the reasonable possibility of relationship between AE and vaccination is caused by the vaccination:

1. Related: there is a suspicion that a link between vaccine and the AE (do not need to be determined); the vaccine has a reasonable potential for promoting the AE.
2. Unrelated: there is no suspicion that a link exists between vaccine and the AE; there are other more likely causes, and vaccination has not been suspected to promote the AE.

### 5.5.5 Treatment of AEs/ARs

An adverse event (AE) is any untoward medical occurrence in a patient or clinical trial participant administered with a pharmaceutical product and which does not necessarily have a causal relationship with this treatment.

Adverse reactions (AR): unexpected or harmful reactions in the process of vaccination according to the prescribed dose and procedure, usually related to vaccination.

Serious adverse event (SAE): refers to the following important medical events, whether or not related to the vaccine clinical trial, including: 1) death; 2) life threatening; 3) hospitalization or prolonged hospitalization; 4) permanent or significant disability / loss of function; 5) congenital abnormality or birth defect; 6) severe adverse event It may lead to other important medical events, such as those listed above without treatment.

Suspected Unexpected Serious Adverse Reaction (SUSAR): Suspected adverse reactions refer to the adverse reactions of subjects at any dose that have nothing to do with the purpose of the medication. After analysis, it is considered that the relationship with the drug is at least likely to be related; Unexpected refers to adverse reactions. The nature, extent, consequences, or frequency are different from the expected risks described in the previous plan or other related materials (such as the investigator’s manual and instructions).

If subjects have any clinically significant disease/event after vaccination, it should be reported to the investigators as soon as possible. The investigators should follow up the adverse reaction/event until the symptoms disappear or the symptoms stabilize. When the investigators deem it necessary, treatment will be provided unconditionally to relieve the pain caused by the adverse reaction/event for the subjects. All medical treatments will be recorded at each follow-up.

In the event of a serious adverse event/reaction, the investigators should take necessary measures quickly, fill in the “Serious Adverse Event Report Form” within 24 hours, and report it to the principal investigator in the form of fax or E-mail.

### 5.5.6 Treatment of pregnancy events

Vaccination of the trial is not allowed during pregnancy. Before vaccination, the subjects will be given urine pregnancy test, and those who are positive in the urine pregnancy test should not be included in the group. If a pregnancy event occurs within 6 months of the visit, the Pregnancy Case Survey Form needs to be filled out.

## 5.6 Immunogenicity evaluation

### 5.6.1 Samples collection

“0-28 days” immunization regimen:

At V0 (day 28 after the prime dose and before the booster dose) and V3 (day 28 after the booster dose), 20 ml of venous blood will be collected by vacuum anticoagulant blood collection vessel. PBMC and serum will be separated to detect the antibody level, immune cell differentiation and antibody spectrum induced by the vaccine.

In V2 (day 14 after the booster dose) and V4 (month 6 after the booster dose), 10ml of venous blood will be collected by vacuum anticoagulant blood collection vessel, and the serum will be separated to detect the antibody level, immune cell differentiation and antibody spectrum induced by the vaccine.

“0-56 days” immunization regimen:

At V-1 (day 28 after the prime dose) and V3 (day 28 after the booster dose), 20 ml of venous blood will be collected by vacuum anticoagulant blood collection vessel. PBMC and serum will be separated to detect the antibody level, immune cell differentiation and antibody spectrum induced by the vaccine.

At V0 (day 56 after the prime dose and the booster dose), V2 (day 14 after the booster dose) and V4 (month 6 after the booster dose), 10ml of venous blood will be collected by vacuum anticoagulant blood collection vessel, and the serum will be separated to detect the antibody level, immune cell differentiation and antibody spectrum induced by the vaccine.

The serum samples of this clinical trial will be used to evaluate the immune response level of the research vaccine. For other studies, the approval of the ethics committee and the consent of the subjects are required.

### 5.6.2 Preservation and transportation of samples

Unified operation standards are adopted in the process of preservation and transportation. The storage temperature of serum should be - 20℃ and below, and it should be transported to the testing laboratory in time. The separation, transportation and preservation of BPMC used for the differentiation of immune cell population and the detection of antibody spectrum are operated by the third party laboratory according to the standard operating procedures.

## 5.7 Data management

### 5.7.1 Data collection, entry and reporting

Vaccination and visit records and other original data should be clearly recorded, which should be filled in with a black signature pen. The errors of the original records should not be wiped or covered, but should be crossed off, put the corrected data aside, and signed and dated by the investigators.

Fill in all the case report forms according to the protocol. Case report form is used to record the data of clinical trials. It is an important part of clinical trials and research reports. It should be filled in clearly and completely. It is required to fill in with a black signature pen. The mistakes should not be erased or covered by the original record. Instead, a horizontal line should be drawn on it, and the corrected data should be indicated in the blank beside it. The revised researcher should sign his name and indicate the date.

According to the requirements of the scheme, data collection, biological sample collection and examination should be carried out in each visit time window, and the original documents and records should be complete, and the examination conclusions should be entered into the case report form (CRF) in time.

### 5.7.2 Verification of data records

The quality controller should check the data record regularly and irregularly until the CRF is completed. Before withdrawing the CRF, the quality controller should carefully check the CRF number of the subjects, the number of pages of each CRF and the necessary signature of the researcher. The main content of quality control should focus on the following links: signing of informed consent, volunteer screening, immunization, management of experimental vaccine, safety observation, collection and preservation of immunogenic samples, etc. the consistency of research data and original data should be focused on, and manual verification should be carried out. The verification results shall be recorded. The transfer of CRF and other research materials should be documented.

### 5.7.3 Database establishment and data entry

Establishment of database:

The personnel in charge of data management shall establish the database structure and check procedures according to CRF, ensure that the database can be correctly converted to SAS file format, and modify and confirm the database structure through trial input.

Further verification of CRF:

Before entering CRF, data management personnel should check CRF again, mainly to see if there are omissions and obvious errors.

Data entry:

After the training of data entry personnel, data entry is carried out by two persons and two computers.

Data comparison and examination:

Check the consistency of the database data independently completed by the two people, report the inconsistent values and information, and then check the original data item by item to correct until the two databases are consistent. Use the computer program that has been written and confirmed to logically check the data, issue the query form and ask the researcher to confirm and then modify the database until there is no doubt. It is necessary to select a certain number of CRF randomly according to a certain proportion to control the quality of the database, and compare with the data in the database manually, so as to ensure that the data in the database is consistent with the CRF content.

### 5.7.4 Database locking

Before statistical analysis, it is necessary to conduct a check and clean of the database. The analyzed population will be set according to the definition of the data sets, including the FAS set, PPS set and safety analysis data set, and to determine the deviation from the regimen and its impact on the analysis data set. After the database is cleaned, the database will be locked, and the statistical analysis plan is locked at the same time.

## 5.8 Statistics Plan and Statistical Analysis

### 5.8.1 Statistical plan

The statistical analysis of this study will be completed in two stages. After the subjects complete the visit 28 days after the booster dose immunization, the study database will be recorded, reviewed and locked for the first stage statistical analysis.

The safety and immunogenicity data after 28 days till 6 months after immunization will be collected, reviewed and locked for the second stage statistical analysis and summary.

### 5.8.2 Selection of analysis data sets

Safety data set (SS):

The safety evaluation should be conducted for all participants who receive vaccines after randomization. Data violating the protocol should not be eliminated.

Immunogenicity data set:

Full Analysis Set (FAS): It is defined as ideal participant population determined according to the ITT (Intention-to-treat analysis) principle, all participants who meet the inclusion / exclusion criteria, and are randomized and given vaccine and have at least one post-immunization blood test results are included in the FAS.

Per-Protocol Set (PPS): It is a subset of FAS. Participants in this set are more compliant with the protocol, experience no major protocol violation, comply with all inclusion criteria / exclusion criteria, and complete the vaccination within the time window as required in the protocol and all blood samplings are included in the PPS set. Participants who violate the trial protocol, such as poor compliance or lost to follow-up, and those who suffer intercurrent SARS-CoV-2 infection will not be included in this analysis set.

In this trial, the FAS will be used as the primary analysis set. However, PPS should be analyzed simultaneously. Any inconsistency between PPS and FAS analysis results should be discussed in the report.

### 5.8.3 Data statistics method

During statistical analysis, first, the number of completed cases and drop-out cases should be checked. Then demographic and baseline characteristics of each group at enrollment should be analyzed to investigate intergroup comparability. Efficacy evaluation of vaccine includes the determination of evaluation indicators and intergroup comparison of efficacy. Safety evaluation includes the statistics of clinical ARs/AEs.

Participant elimination criteria: participants don’t meet the inclusion criteria; data and information after vaccination are not followed up; information and data after randomization are seriously missing; participants meet exclusion criteria but are not withdrawn; participants receive wrong vaccination or incorrect dose.

Safety analysis in this trial mainly includes descriptive analysis of the incidence of ARs/AEs. *χ*^2^ test may be carried out for intergroup comparison, and Fisher’s exact test may be performed if necessary. After immunization, the number of case-times and person-times of local AEs in the high-dose group will be calculated (with conventional calculation method). The number of person-times will be calculated based on the highest severity in both arms, and the number of case-times will be calculated based on the cumulative local AEs actually occurring at the vaccination site. Logarithmic transformation is required for analysis of immunogenicity indicator of antibody level which should be expressed as GMT, standard deviation, median, maximum and minimum and 95% confidence interval. Classification indicators will be compared between groups. Antibody seroconversion rate will be analyzed by *χ*^2^ test and Fisher’s exact test may be used if necessary. Study data at different time points will be analyzed with statistical analysis for repeated measurement data.

SAS 9.4 is adopted for all statistical analyses with two-sided test. The P value is directly calculated while carrying out Fisher’s exact test when test statistics and corresponding P values are given, and in case of P ≤ 0.05, the difference is statistically significant.

## 6. Monitoring of Clinical Trial

### 6.1 Quality assurance and quality control

Carry out on-site quality control in strict accordance with the relevant requirements of Good Clinical Practice (GCP).

Investigators in some positions are qualified as physicians or above. Prior to the clinical trial, they will be trained in the clinical protocol and all trial procedures, including information about the trial vaccine, procedures for obtaining informed consent, operating procedures for each position, and procedures for reporting adverse reactions/events.

The data of each subject is reviewed at each stage of the clinical trial to ensure that the content of the clinical trial meets the requirements of the protocol and that the obtained data are complete and reliable. The quality controller controls the whole process of the clinical trial.

All the work on site are carried out strictly in accordance with the clinical trial field operation manual. Each subject records the “*Diary Card*” by themselves, follows up and retrospectively investigats by the researcher, and reviews and guides the filling in of the “*Diary Card*”.

The quality controller shall conduct a comprehensive check on the original data, and after training, a special person shall enter the data of eCRF. The double entry method shall be adopted and completed by two people independently.

Calibration or standardization of the instruments used in this clinical trial.

### 6.2 Modification of clinical protocol

After this plan is approved by the Ethics Committee, if there is any major modification in the implementation process, it shall be reported to the Ethics Committee for approval before it can be implemented. The investigators shall not execute any deviation or change without the consent of the Sponsor and prior review and written approval of the Ethics Committee (EC).

Any changes to the scheme, whether material or non-material, are required to be in writing. EC approval is required to identify substantive protocol changes that would affect the safety of subjects, the scope of the study, or the scientific quality of the study.

### 6.3 Scheme deviation

The investigators shall carry out the clinical trial according to protocol approved by the ethics committee and the provisions of GCP. During the trial, the researcher shall not deviate from the protocol unless the harm to the subjects is eliminated.

The research center shall record all protocol deviations in the original data of subjects, including but not limited to the occurrence time of protocol deviation, discovery time, event description and measures, etc. In case of serious protocol deviations, the main researchers should be informed in time and report to the IEC.

### 6.4 Confidentiality

The sponsor, investigators, IEC, or a fully authorized representative of regulatory authority should have the right to obtain data related to the clinical trial, but relevant content cannot be used for any other clinical trials, nor can it be disclosed to any other individuals or entities.

Investigators must sign a confidentiality agreement to confirm that he/she knows and agrees to hold the information of this study confidential.

Investigators and other study personnel should keep all information provided by the sponsor and all data/information generated at the study site (except for medical records of participants) confidential. Such information and data should not be used for any purposes other than the study. This restriction does not apply to: (1) study information is not disclosed because of violations by investigators and researchers; (2) study information is disclosed only to the IRB/IEC for the purpose of study evaluation; (3) study information is disclosed to provide appropriate medical assistance to participants.

## 7. Schedules

In this study, it will take about 10 months from the preparation before initiating this study to the completion of the summary report. The schedule of clinical trial is shown as follows (for reference only):

| Implementation process of clinical trial | Proposed duration |
| --- | --- |
| 1. Preparation before clinical trial | 18 days |
| 2. IEC Review Approval | 3 days |
| 3. Recruitment and enrollment of the first participant | 1 month |
| 4. 28-day visit after the last participant completes the last dose |  |
| 5. First analysis | 1 month |
| 6. First analysis report |  |
| 7. 6-month visit after the last participant completes the last immunization | 6 months |
| 8. Final analysis | 1 month |
| 9. Summary Report |  |

## 8. Ethical Approval

### 8.1 Ethical Review and Approval

The PI should submit the clinical trial protocol and all necessary additional documents to the IEC for initial review:

- Clinical study protocol (indicated with version No. / date)
- Informed Consent Form (indicated with version No./date)
- Participant recruitment materials (indicated with version No. / date)
- Diary card (indicated with version No. / date)
- Contact card (indicated with version No. / date)
- Vaccination Visit Record (indicated with version No./date)
- PI’s CV
- Drug clinical trial approval from the NMPA

After the above study documents are reviewed and approved by the IEC, a written approval certificate will be issued to the investigators.

### 8.2 Follow-up Review

Whether the methods for inclusion of participants and the information provided to participants are complete and understandable; whether the methods for obtaining informed consent are appropriate; whether SAEs are reported timely; and whether timely medical treatment can be provided for the SAE related to inoculation with the candidate vaccine.

During the whole trial, the IEC should supervise whether the risk-benefit ratio of the study is increased and whether the rights and interests of participants are effectively protected.

### 8.3 Potential risks and minimization of risks

#### 8.3.1 Benefits and risks

It is expected that participants in this program will likely gain an improved level of immune response against SARS-CoV-2 from a booster dose of COVID-inactived vaccine or immune protection against influenza viruses from a booster dose of influenza vaccine. Mass vaccinations of both inactivated and influenza vaccines have demonstrated good safety, so no significant increase in safety risk is expected with an additional dose of vaccine. Participants in this clinical trial will not need to pay for the inoculation with investigational vaccines. Participants in this trial are provided with reasonable transportation fee, charge for loss of working time, blood collection compensation and nutrition fee. During participating in this clinical trial, participants will receive one dose of inactived COVID-19 vaccine or influenza vaccine. At the same time, injection of vaccines may cause some ARs. Common ARs of vaccination include: pyrexia, injection site tenderness, redness and swelling. Generally, the AEs will be alleviated within 3 - 5 days after occurrence.

#### 8.3.2 Vaccination

Qualified inoculation consumables will be purchased, and aseptic inoculation will be performed in strict accordance with standard method to avoid AEs caused by improper inoculation or inoculation error.

If a participant experiences a grade 3 or above AR during the safety observation period, or experiences a SAE related or possibly related to the candidate vaccine, he/she should be able to receive timely medical treatment, and if necessary, the “green channel for medical treatment” should be immediately initiated for emergency treatment.

#### 8.3.3 Blood specimen collection

After qualification review by the PI, the experienced nursing staff will be employed to collect venous blood samples after training as per the specified procedures to minimize pains or risks (including pain and less chance of venipuncture site infection) from which the participants suffered.

## Notes

### Competing Interest Statement

I have read the journal's policy and the authors of this manuscript have the following competing interests: Jingxin Li reports grants from National Natural Science Foundation of China (grant 82173584). Fengcai Zhu reports grants from Jiangsu Provincial Key Research and Development Program grant (BE2021738). Wei Chen, Yanan Zhang and Cancan Chen are the employees of Anhui Zhifei Longcom Biopharmaceutical. All other authors declare no competing interest.

### Clinical Trial

ClinicalTrial.gov NCT04833101

### Author Declarations

Ethics Committee of Jiangsu Provincial Center for Disease Prevention and Control (approval number: JSJK2021-A005-02)

## References

1. World Health Organization, Impact of COVID-19 on people’s livelihoods, their health and our food systems. https://www.who.int/news/item/13-10-2020-impact-of-covid-19-on-people’s-livelihoods-their-health-and-our-food-systems. Updated October 13, 2020. Accessed February 7, 2022.

2. Our World in Data, Coronavirus (COVID-19) Vaccinations. https://ourworldindata.org/covid-vaccinations. Updated February 16, 2022. Accessed February 17, 2022.

3. World Health Organization, COVID-19 vaccination dashboard. https://www.who.int/emergencies/diseases/novel-coronavirus-2019/covid-19-vaccines. Accessed 2 January, 2022.

4. Khoury DS, and Cromer D. Neutralizing antibody levels are highly predictive of immune protection from symptomatic SARS-CoV-2 infection. Nat Med. 2021;27(7):1205–11.

5. Krause PR, Fleming TR, Peto R, Longini IM, Figueroa JP, Sterne JAC, et al. Considerations in boosting COVID-19 vaccine immune responses. Lancet (London, England). 2021;398(10308):1377–80.

6. Tartof SY, Slezak JM, Fischer H, Hong V, Ackerson BK, Ranasinghe ON, et al. Effectiveness of mRNA BNT162b2 COVID-19 vaccine up to 6 months in a large integrated health system in the USA: a retrospective cohort study. Lancet. 2021;398(10309):1407–16.

7. Cohn BA, and Cirillo PM. SARS-CoV-2 vaccine protection and deaths among US veterans during 2021. Science. 2022;375(6578);331–336.

8. Logunov DY, Dolzhikova IV, Zubkova OV, Tukhvatullin AI, Shcheblyakov DV, Dzharullaeva AS, et al. Safety and immunogenicity of an rAd26 and rAd5 vector-based heterologous prime-boost COVID-19 vaccine in two formulations: two open, non-randomised phase 1/2 studies from Russia. Lancet. 2020;396(10255):887–97.

9. Logunov DY, Dolzhikova IV, Shcheblyakov DV, Tukhvatulin AI, Zubkova OV, Dzharullaeva AS, et al. Safety and efficacy of an rAd26 and rAd5 vector-based heterologous prime-boost COVID-19 vaccine: an interim analysis of a randomised controlled phase 3 trial in Russia. Lancet. 2021;397(10275):671–81.

10. Greinacher A, Thiele T, and Warkentin TE. Thrombotic Thrombocytopenia after ChAdOx1 nCov-19 Vaccination. N Engl J Med. 2021;384(22):2092–101.

11. Scully M, Singh D, Lown R, Poles A, and Solomon T. Pathologic Antibodies to Platelet Factor 4 after ChAdOx1 nCoV-19 Vaccination. N Engl J Med. 2021;384(23):2202–11.

12. MacNeil JR, Su JR, Broder KR, Guh AY, Gargano JW, Wallace M, et al. Updated Recommendations from the Advisory Committee on Immunization Practices for Use of the Janssen (Johnson & Johnson) COVID-19 Vaccine After Reports of Thrombosis with Thrombocytopenia Syndrome Among Vaccine Recipients - United States, April 2021. MMWR Morb Mortal Wkly Rep. 2021;70(17):651–6.

13. Centers for Disease Control and Prevention, COVID-19 Vaccine Booster Shots. https://www.cdc.gov/coronavirus/2019-ncov/vaccines/booster-shot.html. Updated February 2, 2022. Accessed February 7, 2022.

14. World Health Organization, Interim recommendations for heterologous COVID-19 vaccine schedules. https://www.who.int/publications/i/item/WHO-2019-nCoV-vaccines-SAGE-recommendation-heterologous-schedules. Updated December 16, 2022. Accessed February 7, 2022.

15. Sablerolles RSG, Rietdijk WJR, Goorhuis A, Postma DF, Visser LG, Geers D, et al. Immunogenicity and Reactogenicity of Vaccine Boosters after Ad26.COV2.S Priming. N Engl J Med. [published online Jan 19, 2021]. https: doi: 10.1056/NEJMoa2116747.

16. Stuart ASV, Shaw RH, Liu X, Greenland M, Aley PK, Andrews NJ, et al. Immunogenicity, safety, and reactogenicity of heterologous COVID-19 primary vaccination incorporating mRNA, viral-vector, and protein-adjuvant vaccines in the UK (Com-COV2): a single-blind, randomised, phase 2, non-inferiority trial. Lancet. 2021;399(10319):36–49.

17. Munro APS, Janani L, Cornelius V, Aley PK, Babbage G, Baxter D, et al. Safety and immunogenicity of seven COVID-19 vaccines as a third dose (booster) following two doses of ChAdOx1 nCov-19 or BNT162b2 in the UK (COV-BOOST): a blinded, multicentre, randomised, controlled, phase 2 trial. Lancet. 2021;398(10318):2258–76.

18. Yang S, Li Y, Dai L, Wang J, He P, Li C, et al. Safety and immunogenicity of a recombinant tandem-repeat dimeric RBD-based protein subunit vaccine (ZF2001) against COVID-19 in adults: two randomised, double-blind, placebo-controlled, phase 1 and 2 trials. The Lancet Infectious diseases. 2021;21(8):1107–19.

19. Zhu F, Jin P, Zhu T, Wang W, Ye H, Pan H, et al. Safety and immunogenicity of a recombinant adenovirus type-5-vectored COVID-19 vaccine with a homologous prime-boost regimen in healthy participants aged 6 years and above: a randomised, double-blind, placebo-controlled, phase 2b trial. Clin Infect Dis. [published online September 22, 2021]. https:doi: 10.1093/cid/ciab845.

20. Zhu FC, Li YH, Guan XH, Hou LH, Wang WJ, Li JX, et al. Safety, tolerability, and immunogenicity of a recombinant adenovirus type-5 vectored COVID-19 vaccine: a dose-escalation, open-label, non-randomised, first-in-human trial. Lancet. 2020;395(10240):1845–54.

21. Li J, and Hui A. Safety and immunogenicity of the SARS-CoV-2 BNT162b1 mRNA vaccine in younger and older Chinese adults: a randomized, placebo-controlled, double-blind phase 1 study. Nat Med. 2021;27(6):1062–70.

22. World Health Organization. WHO International Standard First WHO International Standard for anti-SARS-CoV-2 immunoglobulin (human). 2021.

23. Stanley A. Plotkin WAO, Paul A. Offit, Kathryn M. Edwards. Plotkin’s Vaccines. In: Siegrist CA, eds. Vaccine Immunology. Elsevier Press; 2018: 24–26.

24. Zhao X, Li D, Ruan W, Chen Z, Zhang R, Zheng A, et al. Effects of a Prolonged Booster Interval on Neutralization of Omicron Variant. [published online January 26, 2022]. N Engl J Med. https:doi: 10.1056/NEJMc2119426.

25. Heaton PM. Booster dose of Janssen COVID-19 vaccine (Ad26.COV2.S) following primary vaccination. Presentation sliders presented at the meeting of Advisory Committee on Immunization Practices (ACIP); October 21, 2021. https://www.cdc.gov/vaccines/acip/meetings/downloads/slides-2021-10-20-21/03-COVID-Heaton-Douoguih-508.pdf.

26. Voysey M, Costa Clemens SA, Madhi SA, Weckx LY, Folegatti PM, Aley PK, et al. Single-dose administration and the influence of the timing of the booster dose on immunogenicity and efficacy of ChAdOx1 nCoV-19 (AZD1222) vaccine: a pooled analysis of four randomised trials. Lancet. 2021;397(10277):881–91.

27. Ai J, Zhang H, and Zhang Q. Recombinant protein subunit vaccine booster following two-dose inactivated vaccines dramatically enhanced anti-RBD responses and neutralizing titers against SARS-CoV-2 and Variants of Concern. Cell Res. 2021;32(1):103–106.

28. Halperin SA, Ye L, MacKinnon-Cameron D, Smith B, Cahn PE, Ruiz-Palacios GM, et al. Final efficacy analysis, interim safety analysis, and immunogenicity of a single dose of recombinant novel coronavirus vaccine (adenovirus type 5 vector) in adults 18 years and older: an international, multicentre, randomised, double-blinded, placebo-controlled phase 3 trial. Lancet. 2022;399(10321):237–248.

29. Anhui Zhifei Longcom Biopharmaceutical Co. L. A study published in the New England Journal of Medicine showed that the recombinant subunit vaccine ZF2001 (Zecvide ®) effectively induced antibodies to neutralize the omicron mutant strain. http://www.zflongkema.com/news/qyyw/2022-01-28/332.html. Updated January 28, 2022. Accessed February 7, 2022.

30. Feng S, and Phillips DJ. Correlates of protection against symptomatic and asymptomatic SARS-CoV-2 infection. Nat Med. 2021;27(11):2032–40.

31. He X, and Hong W. SARS-CoV-2 Omicron variant: Characteristics and prevention. MedComm. 2021;2(4):838–845.

32. Cameroni E, Saliba C, Bowen JE, Rosen LE, Culap K, Pinto D, et al. Broadly neutralizing antibodies overcome SARS-CoV-2 Omicron antigenic shift. [published online December 23, 2021]. Nature. https:doi: 10.1038/s41586-021-04386-2.

33. Lu L, Mok BW, Chen LL, Chan JM, Tsang OT, Lam BH, et al. Neutralization of SARS-CoV-2 Omicron variant by sera from BNT162b2 or Coronavac vaccine recipients. Clin Infect Dis. [published online December 16, 2021]. Clin Infect Dis. https: doi: 10.1093/cid/ciab1041.

